# Time-Scale Target Parameters and Two-Step Estimation in Longitudinal Trials for Progressive Diseases

**DOI:** 10.1101/2024.05.31.24307898

**Authors:** Florian Stijven, Craig Mallinckrodt, Geert Molenberghs, Ariel Alonso, Sam Dickson, Suzanne Hendrix

**Affiliations:** I-BioStat, KU Leuven, Belgium; Pentara Corporation, Utah, United States; I-BioStat, Universiteit Hasselt, Belgium

**Author notes:** **Correspondence** Florian Stijven. Kapucijnenvoer 7, 3000 Leuven, Belgium.

**Keywords:** Two-Step Estimator, Longitudinal Data, Progressive Diseases, Alzheimer’s Disease

## Abstract

In progressive diseases such as Alzheimer’s, treatments that slow progression should start early to preserve higher levels of functioning for a longer period. In corresponding clinical trials, treatment effects are usually expressed as mean differences on a clinical scale at fixed time points. Early in the disease course, however, these mean differences may appear small but may nonetheless correspond to an important slowing of disease progression. This complicates the appreciation of the relevance of observed treatment effects. We introduce a class of target parameters that quantify treatment effects on the time scale in longitudinal studies; for instance, in terms of time saved or percentage slowing of progression. We focus on data from randomized trials where the target parameters are identified under regularity assumptions. These target parameters remain well defined if treatment was not randomized, but additional untestable assumptions are required for identification. We propose general two-step estimators. In the first step, the data can be analyzed with standard methods for longitudinal data and standard software can thus be used. In the second step, summary statistics from the first step are used for inferences about the target parameters. The second step has been implemented in the *TCT* R package. We study the asymptotic properties and efficiency of these two-step estimators, and evaluate them in an extensive simulation study. These estimators are used in a phase 2/3 clinical trial for Alzheimer’s disease, leading to important additional insights into the treatment effect.

## 1 INTRODUCTION

It is often recommended to start disease-modifying treatments (DMT) early for progressive diseases to maintain lower symptom severity and higher functioning levels for a longer time ^1^. However, disease progression is typically slow in the early disease stages, resulting in small effect sizes for DMTs. Nonetheless, small effect sizes early on can imply a considerable slowing of the disease progression. In contrast, symptomatic treatments may have a large effect early on, without slowing disease progression on the long term. This poses considerable challenges for the design, analysis, and interpretation of clinical trials for DMTs in progressive diseases.

A recent trial in Alzheimer’s disease (AD) highlights these challenges ^2^. In this study, patients with mild cognitive impairment (MCI)—a preclinical stage of AD—progressed more slowly than patients with mild AD. In the placebo group, MCI patients deteriorated on average 1.25 points on the Clinical Dementia Rating – Sum of Boxes (CDR-SB) scale ^3^, while mild AD patients deteriorated on average 2.30 points. A similar pattern emerged for the treatment effect: a mean drug–placebo difference of –0.62 in the mild AD group but only -0.35 in the MCI group. Although the absolute differences suggest that the drug is more effective in later AD stages, the mean changes under active treatment were about 28% less than those under placebo in both groups. Hence, the drug’s “relative” effects may be comparable across different stages of AD, even though the absolute effects (which emphasize symptom reduction) are larger in patients with mild AD.

Target parameters that quantify treatment effects on the time scale express treatment effects in terms of time saved (e.g., progression has been delayed with 3 months after 1 year of treatment) or percentage slowing of progression (e.g., treatment slows progression with 25%) ^4,5,6^. Raket ^4^ introduced non-linear parametric models for longitudinal data where such target parameters coincide with the parametric model’s parameters. We extend this work by introducing a general class of non- and semi-parametric target parameters which reduce to Raket’s parameters under certain parametric assumptions. These parameters are causal and, consequently, are only identifiable under typical untestable assumptions from the causal inference literature. In this paper, however, we focus on data from randomized trials where these untestable assumptions hold by design.

We propose two-step estimators for the target parameters. In the first step, practically any method for longitudinal data can be used; hence, standard software can be used here. For instance, a mixed model for repeated measures (MMRM), which is a standard method for longitudinal data from clinical trials, may be used here ^7^. Semi- or non-parametric methods may be used as well ^8^. In the second step, summary statistics obtained from the first step are used for estimation and inferences about the target parameters. The second step has been implemented in the *TCT* R package (available from github.com/florianstijven/TCT). We study the asymptotic properties and efficiency of these two-stage estimators and evaluate them in simulations.

The remainder of this paper is organized as follows. In Section 2, we introduce a general class of target parameters that quantify treatment effects on the time scale together with identifying assumptions. We introduce two-step estimators and corresponding methods for inference in Section 3 and study their finite-sample properties through simulations in Section 4. We apply the methods to the data that motivated this work in Section 5 and end the paper with a discussion of the proposed methods and possible extensions.

## 2 TARGET PARAMETERS

Raket ^4^ defined target parameters that quantify treatment effects on the time scale through non-linear parametric models for longitudinal data. We extend that work by defining non- and semi-parametric target parameters (i.e., parameters that are defined without having to assume a particular parametric form for the data-generating process) with a similar interpretation within a causal framework.

### 2.1 Notation

Throughout this paper, we consider the setting where patients get assigned to one of two treatments at baseline. The variable *Z* captures the assigned treatment, where *Z* = 0 denotes the control treatment (e.g., placebo or active control) and *Z* = 1 denotes the experimental treatment. After treatment assignment, patients are followed up longitudinally up to time *c*. We let *Y*_*t*_(*z*) be the potential outcome at time *t* after randomization had the patient been assigned treatment *z*. Because each patient is followed from *t* = 0 to *t* = *c*, each patient has two potential functions *t* → *Y*_*t*_(0) and *t* → *Y*_*t*_(1) as “potential outcomes”, reflecting how the response variable (e.g., a clinical scale) would have evolved over time had the patient been assigned treatment *z* = 0 or *z* = 1. Viewing these functions as sample paths of stochastic processes, the potential outcomes correspond to two stochastic processes indexed by time: {*Y*_*t*_(0) : *t* ∈ [0, *c*]} and {*Y*_*t*_(1) : *t* ∈ [0, *c*]}. We let the corresponding distribution functions at a given time point *t* be *F*_*Y*(0),*t*_ and *F*_*Y*(1),*t*_, respectively.

In the remainder of this paper, we consider causal target parameters that are specific functionals of the potential stochastic processes defined above. To this end, we define potential trajectories, which are function-valued summaries of these stochastic processes.

#### Definition 1

(Trajectory). Let *F* → *h*(*F*) denote a functional on the set of univariate distribution functions or a subset thereof. The *potential trajectory* for treatment group *Z* = *z* is defined as the following real-valued function:

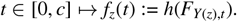

In the simulations and data application, we focus on trajectories where *h* is the mean functional. However, one could consider other choices for *h*, such as the median or other quantiles.

We further refer to *f*_0_ as the *reference trajectory* because treatment effects on the time scale will be defined treating *f*_0_ as the reference.

### 2.2 Causal Target Parameters

The definition of the causal target parameters centers around the *time mapping function* that maps time points from the reference trajectory to *f*_1_.

#### Definition 2

(Time mapping function). Let *t* → *f*_0_(*t*) and *t* → *f*_1_(*t*) be the potential trajectories as defined before. A time mapping function *t* → *g*(*t*) is a real-valued function that satisfies *f*_1_(*t*) = *f*_0_ (*g*(*t*)) for *t* ∈ [0, *c*].

The time mapping function is uniquely defined if and only if *f*_1_(*t*) = *f*_0_(*x*) has a unique solution in *x*, for every *t* ∈ [0, *c*]. We, therefore, make the following two assumptions under which the time mapping function is uniquely defined and can be written as 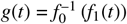.

#### Assumption 1

(Strictly monotone reference trajectory). The reference trajectory *t* → *f*_0_(*t*) is strictly monotone.

#### Assumption 2

(Range restriction). The range of *f*_1_ is contained in the range of *f*_0_: *f*_1_([0, *c*]) ⊆ *f*_0_([0, *c*]).

The time mapping function essentially captures the entire treatment effect on the time scale. For instance, *g*(1 year) = 0.5 years means that 1 year under active treatment corresponds to 6 months under control treatment in terms of disease progression in the trial’s patient population. Hence, progression has been slowed with 50% or has been delayed by 6 months after one year of treatment.

*Remark 1*. The time mapping function is defined through the comparison of trajectories, which are population-level summaries of the potential outcome processes. The time mapping function nonetheless also has an individual-level interpretation: For instance, the treatment effect *g*(1 year) = 0.5 years corresponds (in terms of the functional *h*) to “slowing” each individual’s potential function with 50% after 1 year; more specifically, this corresponds to the following equality: *E*{*Y*_1 year_(1)} = *E*{*Y*_0.5 years_(0)} when *h* is the mean functional. Note, however, that *E*{*Y*_1 year_(1)} = *E*{*Y*_X years_(0)} = *E*{*Y*_0.5 years_(0)} may also hold for *X* a non-degenerate random variable (likely centered around a value close to 0.5 years). Hence, *g*(1 year) = 0.5 years does not imply that each individual’s potential function is slowed with 50% (but the implication holds in the other direction).

If measurements are taken at fixed time points, *t* → *g*(*t*) may only be identified at certain time points because *f*_0_(*t*) can only be identified if measurements were taken at time *t* (and other identifying assumptions hold). On the other hand, if the timing of measurements is random, *g*(*t*) may not be estimable at root-*n* rate. Both issues are solved by parameterizing *t* → *g*(*t*) by a finite-dimensional parameter ***γ*** ∈ ℝ^*p*^. We discuss several choices for this parameterization next.

#### 2.2.1 Time-Specific Acceleration Factors

If there are *K* + 1 fixed measurement occasions, including a baseline measurement, *f*_0_ and *f*_1_ can only be identified at the corresponding times {*t*_0_ = 0, *t*_1_, …, *t*_*K*_}. If we further assume that *g*(0) = 0, which holds if treatment cannot have an instantaneous effect (as treatment is assigned at *t*_0_ = 0), then we may hope to identify *g*(*t*_1_) to *g*(*t*_*K*_). In this respect, we parametrize *g* by ***γ*** = (*γ*_1_, …, *γ*_*K*_)^⊤^ such that

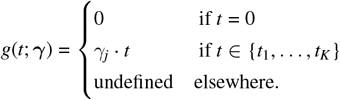

Note that this parameterization does not restrict the distribution of the potential outcomes. We further term *γ*_*j*_ the *time-specific acceleration factor* at *t*_*j*_ because the interpretation is similar to that of an acceleration factor in accelerated failure time models. For instance, *γ*_*j*_ = 0.4 means that the progression rate in the active treatment group, between randomization and *t*_*j*_ = 1 year, was only 40% of the progression rate in the control group. In other words, after 1 year of treatment, we expect patients to only progress 0.40 · 12 = 4.8 months instead of 12 months if they receive the active treatment. This time mapping is illustrated in Figure 1.

**FIGURE 1.**
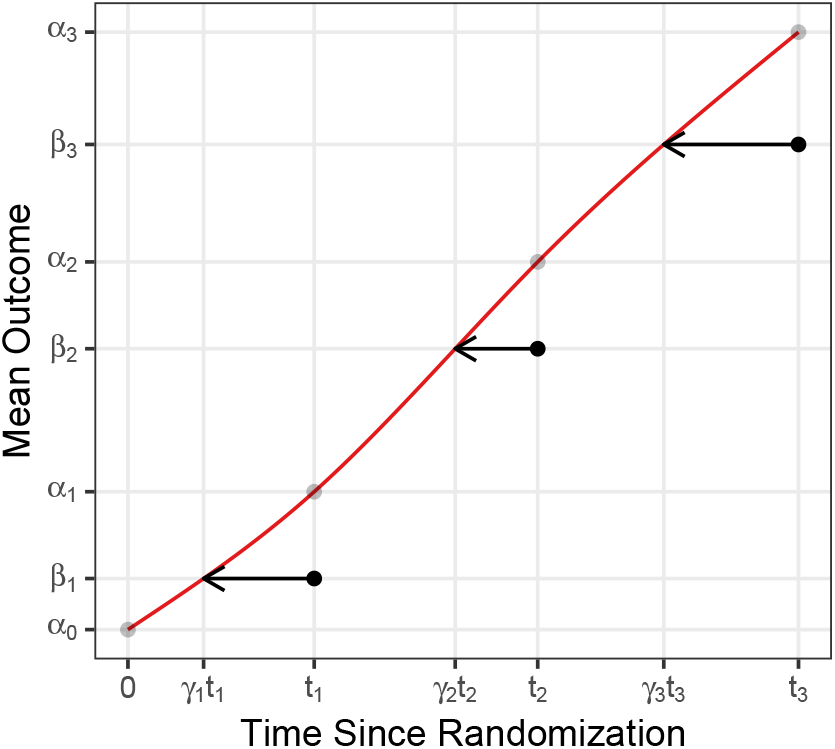
Illustration of the time mapping that underlies the time-specific acceleration factors. The red line is the reference trajectory *f*_0_. The gray dots on this line are the mean outcomes in the control group at the measurement occasions. The black dots are the corresponding mean outcomes in the active treatment group. The arrows visualize the time mapping from *t*_*j*_ to *γ*_*j*_*t*_*j*_. The length of these arrows corresponds to “the amount of time saved” after *t*_*j*_ time units of treatment. The y-axis plots the mean outcome; hence, *h* is the mean functional.

Effective DMTs slow progression and thus imply that *γ*_*j*_ ∈ (0, 1) ∀ *j*. Treatments that reverse progression would have *γ*_*j*_ < 0, but we do not consider this possibility further and, anyhow, Assumption 2 does not allow for a reversal of progression.

#### 2.2.2 Proportional Slowing of Disease Progression

To reduce the number of parameters that describe the treatment effect, we could assume that the treatment causes a *proportional slowing* of disease progression, which corresponds to the following restriction on the time mapping function: *g*(*t*) = *γ* · *t*. In relation to the time-specific acceleration factors, this implies that *γ*_1_ = · · · = *γ*_*K*_, which obviously restricts the distribution of the potential outcomes. This generally has testable implications.

If the proportional-slowing assumption holds and *γ* ∈ (0, 1), the treatment can be said to *proportionally* slow the disease progression, although this provides no guarantee that the disease will continue to progress *more slowly* beyond the clinical trial’s time window. Using the terminology of accelerated failure time models, we now have a constant acceleration factor *γ* which we refer to as the *common acceleration factor*.

### 2.3 Observed Data and Identification

The causal target parameters introduced above are functionals of the potential stochastic processes. However, the observed data are not sampled from these stochastic processes. The observed data are assumed to be (*X, Z*, ***Y***) where *X* ∈ *X* are baseline covariates, *Z* ∈ {0, 1} is the assigned treatment at baseline, and 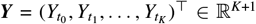 are the observed responses at fixed time points *t*_0_ to *t*_*K*_ where we assume that *t*_0_ = 0. Next, we list a set of untestable assumptions that identify the time mapping function at the measurement occasions with a statistical parameter (i.e., a function of the observed-data distribution *P*_0_).

The first step towards identification of the time mapping function is the identification of the trajectories. Under the following standard causal-inference assumptions ^9^, the trajectories *f*_0_ and *f*_1_ evaluated in *t* ∈ {*t*_0_, …, *t*_*K*_} are identified.

#### Assumption 3

(Conditional exchangeability).

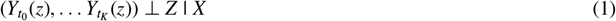

#### Assumption 4

(Positivity of treatment assignment). For *P*_0_ almost all *x*,

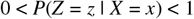

#### Assumption 5

(Consistency). If an individual receives treatment *Z* = *z*, then their observed outcome 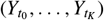 is equal to their potential outcome under treatment *z*:

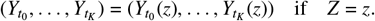

Finally, the following interpolation assumption is required for identifying the trajectories on [0, *t*_*K*_].

#### Assumption 6

(Interpolation). Given *f*_0_ and *f*_1_ evaluated at the measurement occasions {*t*_0_, …, *t*_*K*_}, a known interpolation method yields the correct trajectories on [0, *c*].

*Remark 2*. This interpolation assumption will not hold in practice as there exist many interpolation methods and there is no strong reason to prefer one of them. The time mapping function *g* will generally be less sensitive to the interpolation method if there are more measurement occasions and, for a sufficiently large number of measurement occasions, we expect the methods presented in the next section to be relatively insensitive to the interpolation method.

Under Assumptions 3–6, *f*_0_ and *f*_1_ are identified with the observed-data distribution. The specific statistical parameters that identify these trajectories at a given point depend on the functional *h* used in Definition 1. For instance, for mean trajectories, the g-computation formula ^9^ identifies *f*_*z*_(*t*) for *t* ∈ {*t*_0_, …, *t*_*K*_} as follows:

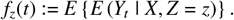

The trajectories evaluated in *t* ∈ [0, *c*] \ {*t*_0_, …, *t*_*K*_} are identified by interpolation.

Given that the trajectories *f*_0_ and *f*_1_ are identified on [0, *c*], the time mapping function is identified as 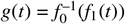 under Assumptions 1 and 2.

## 3 ESTIMATION

In the previous section, we introduced causal target parameters, which are identified with statistical parameters under certain assumptions. In this section, we continue with the setting where the response is measured at fixed measurement occasions and we assume that estimates for (*f*_0_ (*t*_0_), *f*_0_ (*t*_*K*_), …, *f*_0_ (*t*_*K*_)) =:(*α*_0,0_, *α*_1,0_, …, *α*_*K,0*_)=: ***α***_***0***_^⊤^ and (*f*_1_ (*t*_1_), …,*f*_1_ (*t*_*K*_)) =: (*β*_1,0_, …, *β*_*K,0*_)=: ***β***_***0***_ ^⊤^ are available. These estimates constitute the first step of our two-stage procedure and are denoted by 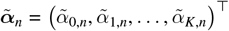 and 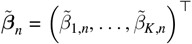, where *n* is the sample size. We assume that 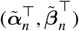 converges to the true values 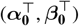 at root-*n* rate and is asymptotically normal.

### Assumption 7.

The estimator 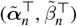 is root-*n* consistent and asymptotically normal:

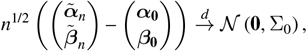

where 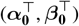 are the true values and 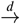 denotes convergence in distribution. A consistent estimator for the positive definite matrix Σ_0_ is available and is denoted by 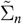

The trajectories evaluated in *t* ∈ [0, *t*_*K*_ ]\{*t*_0_, …, *t*_*K*_} can be estimated by interpolating between 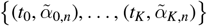 for *f*_0_ and 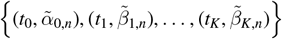 for *f*_1_. Hence, we can write the estimated trajectories as functions parameterized by these estimates: 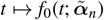 and 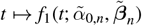. We can similarly write the true trajectories as *t* → *f*_0_(*t*; ***α***_**0**_) and *t* →*f*_1_(*t*; *α*_0,0_, ***β***_0_).

In the remainder of this section, we introduce two-stage estimators and inferential procedures. In the first step, 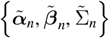 are computed using any method satisfying Assumption 7. The second step depends on the data only through 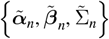 and is explained in the remainder of this section. Estimators obtained in the second step will be denoted by a hat (instead of a tilde). We also provide heuristic justifications for the presented procedures, but technical results and proofs are deferred to Appendix A.

### 3.1 Estimation and Inference Based on Contrast Functions

In this subsection, we discuss estimation and inference based on contrast functions that contrast the estimated trajectories with what would have been expected for a particular value of the time-specific acceleration factors ***γ*** = (*γ*_1_, …, *γ*_*K*_)′. Estimators are defined as the roots of these contrast functions. Inference relies on the contrast functions evaluated in the true value for ***γ*** being asymptotically mean-zero normal.

#### 3.1.1 Contrast Functions

To motivate the contrast functions, we start from the following null hypothesis regarding the time-specific acceleration factor at 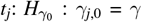. Under this null hypothesis and assuming we know ***α***_**0**_, the mean in the experimental treatment group at *t*_*j*_ is *f*_0_(*γ* · *t*_*j*_; ***α***_**0**_). Consequently, a test for *H*_0_ : *β*_*j*,0_ = *f*_0_(*γ* · *t*_*j*_; ***α***_**0**_) is also a valid test for 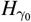. Tests are often available for *H*_0_ : *β*_*j*,0_ = *f*_0_(*γ* · *t*_*j*_; ***α***_**0**_) (e.g., *F*-tests in a linear mixed model ^10^), but they cannot be applied directly since ***α***_**0**_ is not known but estimated. This motivates the following definition of the contrast function:

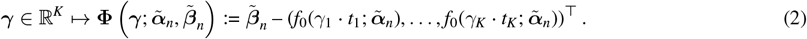

For a given vector of time-specific acceleration factors ***γ***, the contrast function can also be seen as the following function: (***α***^⊤^, ***β***^⊤^)^⊤^ → **Φ**(***γ***; ***α, β***). If the latter function is differentiable at 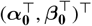, it follows from the delta method that

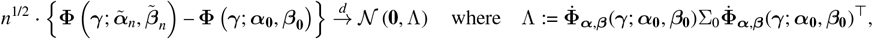

and 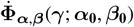 is the Jacobian of (***α***^⊤^, ***β***^⊤^)^⊤^ → **Φ**(***γ***; ***α, β***) evaluated in 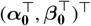.

If ***γ***_**0**_ is the true value for the time-specific acceleration factors, we have that **Φ**(***γ***_**0**_; ***α***_0_, ***β***_0_) = **0**. Hence, a test for *H*_0_ : ***γ***_**0**_ = ***γ*** can be based on the following distributional result:

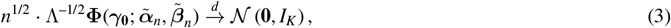

where *I*_*K*_ is the *K × K* identity matrix. In practice, Λ is unknown but can be consistently estimated by 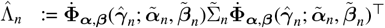 under regularity conditions. By Slutsky’s theorem, we can replace Λ with a consistent estimator without affecting the limiting distribution.

#### 3.1.2 Time-Specific Acceleration Factors

The root of 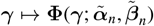, denoted by 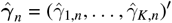, is a consistent and asymptotically normal estimator for the time-specific acceleration factors under regularity conditions (Lemma A.3 in Appendix A). We can use the estimated standard error for 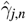 to construct confidence intervals (CIs) based on the normal approximation. Because acceleration factors are expected to be in the unit interval while the estimates may be outside this interval, the normal approximation may be poor. We, therefore, also consider another type of CI.

CIs can also be constructed based on the duality of hypothesis tests and CIs. First note that the *j*’th element of 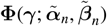 only depends on ***γ*** = (*γ*_1_, …, *γ*_*K*_)′ through *γ*_*j*_. Hence, we can write the *j*’th element as 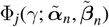 and a test statistic for *H*_0_ : *γ*_*j*,0_ = *γ* follows from the distributional result in (3):

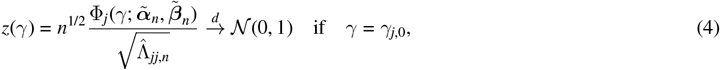

where 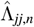 is the *j*’th diagonal element of 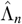 (and 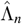 is consistent for Λ). A 1 – *α* CI is obtained as 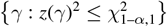 where 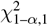 is the 1 – *α* percentile of the chi-squared distribution with one degree of freedom.

Hypothesis tests based on the contrast function in (2) and hypothesis tests based on 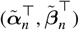 are equivalent for testing the null hypothesis of no treatment effect. This hypothesis, *H*_0_ : *β*_*j*,0_ = *α*_*j*,0_ ∀ *j* = 1, …, *K*, corresponds to the following linear hypothesis:

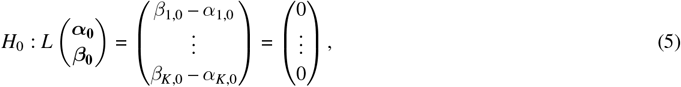

where *L* is an appropriate contrast matrix. A Wald test for (5), or corresponding sub-hypotheses, is based on the following distributional result:

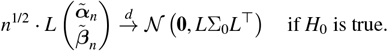

Because **Φ**(***γ***; ***α, β***) = (*β*_1_ – *α*_1_, …, *β*_*K*_ – *α*_*K*_)^⊤^ for ***γ*** = (1, …, 1)^⊤^, we have that

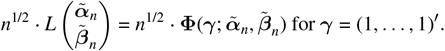

Consequently, Wald-type test statistics for *H*_0_ : *β*_*j*,0_ = *α*_*j*,0_ ∀ *j* = 1, …, *K* are equivalent to corresponding test statistics based on 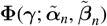 for *γ* = (1,…,1)^┬^.

#### 3.1.3 Common Acceleration Factor

We now describe a class of estimators (defined up to a vector of weights) for the common acceleration factor based on the previously introduced contrast functions. The weights-based estimator for the common acceleration factor, denoted by 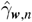, is the root of

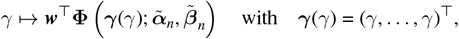

where ***w*** ∈ ℝ^*K*^ is a vector of predefined weights. Lemma A.4 in Appendix A shows that 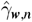 is consistent and asymptotically normal under regularity conditions with a limiting variance that depends on ***w***. Test statistics and CIs can be derived similarly as in Section 3.1.2.

Ideally, one would use weights that minimize the limiting variance; however, these optimal weights are unknown because they depend on unknown parameters. Lemma A.5 in Appendix A shows that we can replace the predefined weights with estimated weights 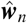 without affecting the limiting distribution as long as the estimated weights converge to fixed weights ***w***_**0**_. Specifically, the limiting distributions using 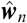 or the unknown ***w***_**0**_ as weights are the same. In the simulations and data application, we define 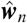 as the weights that minimize the estimated variance of the corresponding weights-based estimator, 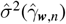:

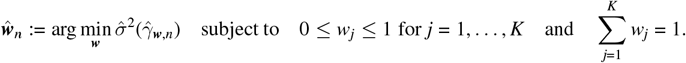

We term the estimator that uses these estimated weights the *adaptive weights-based estimator*.

### 3.2 Estimation and Inference Based on Generalized Least Squares

We propose a second approach based on minimizing a generalized least-squares (GLS) criterion. We can estimate the time-specific acceleration factors by treating 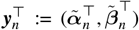 as the *observed data* and fitting a non-linear regression function, parameterized by (***α***^⊤^, ***γ***^⊤^) with ***γ***^⊤^ = (*γ*_1_, …, *γ*_*K*_), to these data:

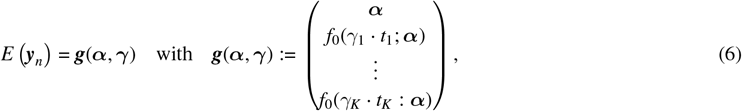

where ***α***^⊤^ = (*f*_0_(*t*_0_; ***α***), …, *f*_0_(*t*_*K*_; ***α***)) because *f*_0_ is an interpolating function. The regression function’s true parameters, (***α***_**0**_^⊤^, ***γ***_**0**_^⊤^), are estimated by minimizing the estimated GLS criterion:

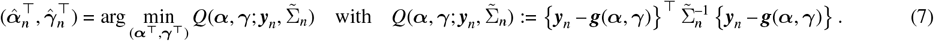

The estimator for the time-specific acceleration factors defined above is equivalent to the contrast-based estimators described before, and 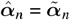.

The common acceleration factor can be estimated by minimizing the same criterion as in (7), but under the constraint that *γ*_1_ = *γ*_2_ = · · · = *γ*_*K*_. This corresponds to redefining the regression function in (6) as follows:

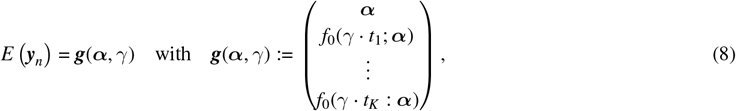

where the regression function’s true parameters, (***α***_**0**_^⊤^, *γ*_0_), are estimated by minimizing the estimated GLS criterion:

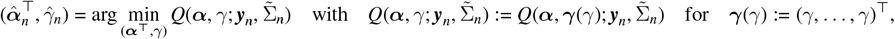

where now 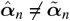.

Lemma A.9 in Appendix A shows that this estimator is consistent and asymptotically normal under regularity conditions. The estimated standard error can be used for constructing CIs based on the normal approximation, but as explained before, the normal approximation may be poor. We, therefore, also consider another type of CI based on the minimized GLS criterion.

If for a fixed *n*, the regression model in (8) would (i) have multivariate normal residuals with a known covariance matrix and (ii) be linear in (***α***^⊤^, *γ*), exact inference based on the minimized GLS criterion would be possible ^11^. Because the residuals are not normal and the regression function is not linear in its parameters, exact inference is not possible. However, Lemmas A.10 and A.11 in Appendix A show that asymptotically valid inference based on the minimized GLS criterion is possible if the residuals are asymptotically normal (i.e., under Assumption 7) and some regularity conditions. Specifically, we show that

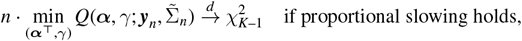

and

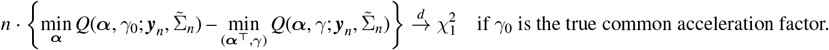

The result in the first display can be used to test for proportional slowing. The result in the second display can be used for testing whether the common acceleration factor equals *γ*_0_, and for constructing CIs based on the duality of hypothesis tests and CIs.

### 3.3 Parametric Bootstrap

The inferential methods described before are all based on asymptotic arguments that essentially rely on Taylor expansions. Such methods may perform poorly in finite samples if the functions involved are highly non-linear around the true parameter values. As a second general strategy for inferences—that may handle non-linearities better but remains only asymptotically valid—we propose a parametric bootstrap procedure that does not require individual-patient data. Specifically, we resample 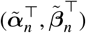 from the estimated multivariate normal sampling distribution:

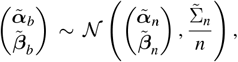

where 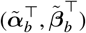 is the *b*’th bootstrap replicate. The *b*’th bootstrap replicate of the common acceleration factor 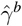 (and similarly for the time-specific acceleration factors) is obtained by applying the estimator to 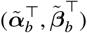 and 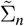 (the latter being kept fixed across bootstrap replications). In the simulations and data application, we subsequently use the percentile interval, but other types of bootstrap intervals could be considered as well.

### 3.4 Efficiency

In Appendix B, we introduce a particular notion of efficiency and corresponding theoretical results for the setting where ***y***_**0**_ ∈ ℝ ^*k*^ is a true vector and one is interested in estimating a particular function *ψ* : ℳ ⊆ ℝ^*k*^ → ℝ evaluated in ***y***_**0**_, and one only observes a random variable ***y***_*n*_ ∈ ℝ^*k*^ such that

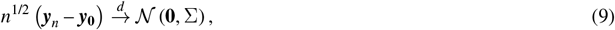

where Σ is assumed to be known. In the above developments, we had that 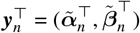 and 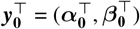. The efficiency theory presented in Appendix B resembles the efficiency theory as presented by van der Vaart ^12^ and Tsiatis ^13^, but these efficiency theories are nonetheless different. However, the efficiency theory in Appendix B is simpler because (9) is, arguably, a simple distributional setup.

In Appendix B.4.1, we show that there can only be one regular and asymptotically linear (RAL) estimator for the time-specific acceleration factors up to asymptotic equivalence. (Regularity and asymptotic linearity are defined in Appendix B.) Consequently, the estimators for the time-specific acceleration factors presented above are efficient. Note, however, that there may exist asymptotically equivalent estimators with different—possibly even better—finite-sample properties.

While there can only be one RAL estimator (up to asymptotic equivalence) for the time-specific acceleration factors, there exist multiple RAL estimators for the common acceleration factor. This class of RAL estimators is characterized in Appendix B. We further show in Appendix B.4.2 that the GLS estimator for the common acceleration factor has the smallest asymptotic variance of all estimators in this class; hence, it is efficient.

## 4 SIMULATIONS

In this section, we summarize the results of our simulation study for evaluating the proposed estimators and inferential procedures. This simulation study has three goals, ordered by importance:

1. Assess to what extent the asymptotic results hold up in finite samples.
2. Identify finite-sample settings where the proposed methods perform poorly.
3. Assess the correctness of the implementation in the *TCT* R package.

### 4.1 Data-Generating Model

The data-generating model for the simulations is based on data from the Alzheimer’s disease neuroimaging initiative (ADNI) ^14^, closely resembling the data-generating model used by Raket ^4^. We use the ADAS-Cog scale as outcome; this is a 13-item cognitive subscale of the Alzheimer’s disease assessment scale, where lower scores indicate less impairment ^15^. Specifically, the data-generating model for the control group is based on a selection of 556 patients from the ADNI for whom the ADAS-Cog scores are available at baseline and 6, 12, 18, 24, and 36 months after baseline. To allow for equally-spaced measurement occasions in our data-generating model, we added a measurement at 30 months. Given these seven potential measurement occasions, we consider the following three measurement patterns:

- *24 Months*. One follow-up visit every 6 months until 24 months after randomization.
- *36 Months*. One follow-up visit every 6 months until 36 months after randomization.
- *36(-30) Months*. Same pattern as 36 Months but leaving out the measurement at 30 months.

We consider a normal and fast progression rate in the placebo group and use the mean functional for defining the trajectories (see Definition 1). The normal progression rate is based on the estimated means from the ADNI patients augmented with the fictitious mean ADAS-Cog score at 30 months, leading to the following mean vector: (19.6, 20.5, 20.9, 22.7, 23.8, 25.8, 27.4). The fast progression rate is not based on data; here, we used the following mean vector: (18.0, 19.7, 20.9, 22.7, 24.7, 27.1, 29.2). In all scenarios, the covariance matrix for the 36 Months pattern is obtained from the following vector of variances and correlation matrix:

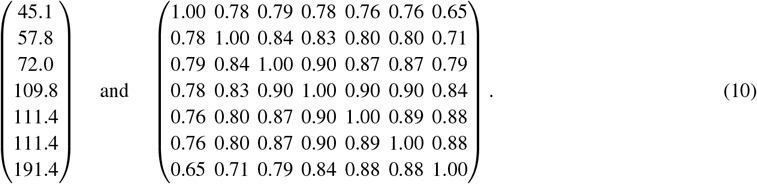

This is the estimated covariance matrix from the ADNI data augmented with an extra row and column for the measurement at 30 months. Note the strong correlation between the measurements across time and the increasing variance; this is realistic but may make inferences harder. The covariance matrices for the other measurement patterns are the corresponding subsets of this matrix.

**FIGURE 2.**
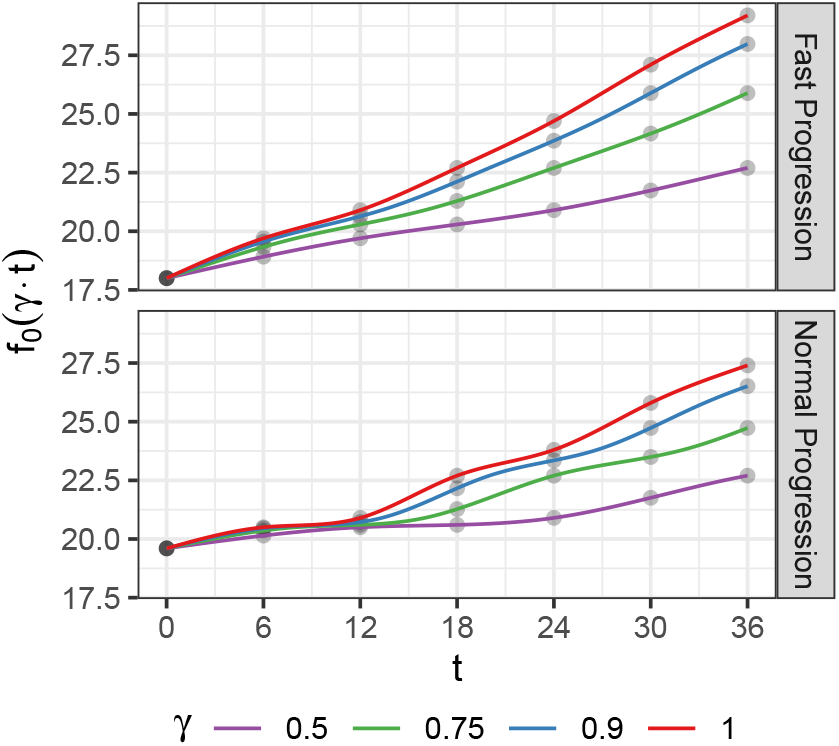
Trajectories underlying the data-generating models. An acceleration factor equal to 1 corresponds to no treatment effect. The dots correspond to potential measurement occasions, and the lines connecting the dots are based on natural cubic spline interpolation.

To simulate proportional slowing, we first consider the reference trajectory, *f*_0_. We have up to now only specified the mean responses in the control group at the seven potential measurement occasions; we interpolate between these points with natural cubic spline interpolation to obtain *f*_0_. The interpolated reference trajectories are plotted as red lines in Figure 2. To simulate data for the treated group (*Z* = 1), we consider trajectories of the following form:

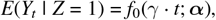

where *γ* ∈ {0.50, 0.75, 0.90, 1}, representing four different treatment effects consistent with proportional slowing.

Finally, we consider four different samples sizes: *n* ∈ {50, 200, 500, 1000}, where *n* is the total sample size and 1:1 randomization is assumed. The data are sampled from multivariate normal distributions with the means determined by the above trajectory and with the covariance matrix defined by (10) or subsets thereof as covariance matrix.

### 4.2 Results

In this section, the results of the simulation study are summarized for the adaptive weights-based and GLS estimators of the common acceleration factor described in Section 3 using natural cubic splines for interpolation (i.e., the “correct” interpolation method). Additional results about these estimators using natural cubic spline or linear interpolation (such that Assumption 6 is violated) are available in Appendix D. To obtain 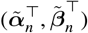 and 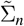—this is step 1 of the two-step estimator—we used a mixed model for repeated measures (MMRM). This is a correctly specified linear mixed model and is a common method for analyzing these type of longitudinal data ^7^. We use the *mmrm* R package to fit these models ^16^; more details on this model are provided in Appendix D.1. We perform 2000 Monte-Carlo replications for every setting, leading to a standard error of 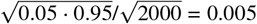 for estimating the empirical coverage of the 95% CIs.

In Figure 3, the means of the estimated acceleration factors are plotted as functions of the sample size. There is a non-negligible bias for the smaller sample sizes, but the bias decreases to almost 0 as the sample size increases to 1000, except for the 24 Months measurement pattern. The latter can be explained by a subtle model misspecification if the measurement pattern in the analysis (i.e., 24 Months) does not match the measurement pattern in the data-generating model (i.e., 36 Months). This is further explained in Appendix D.3.

**FIGURE 3.**
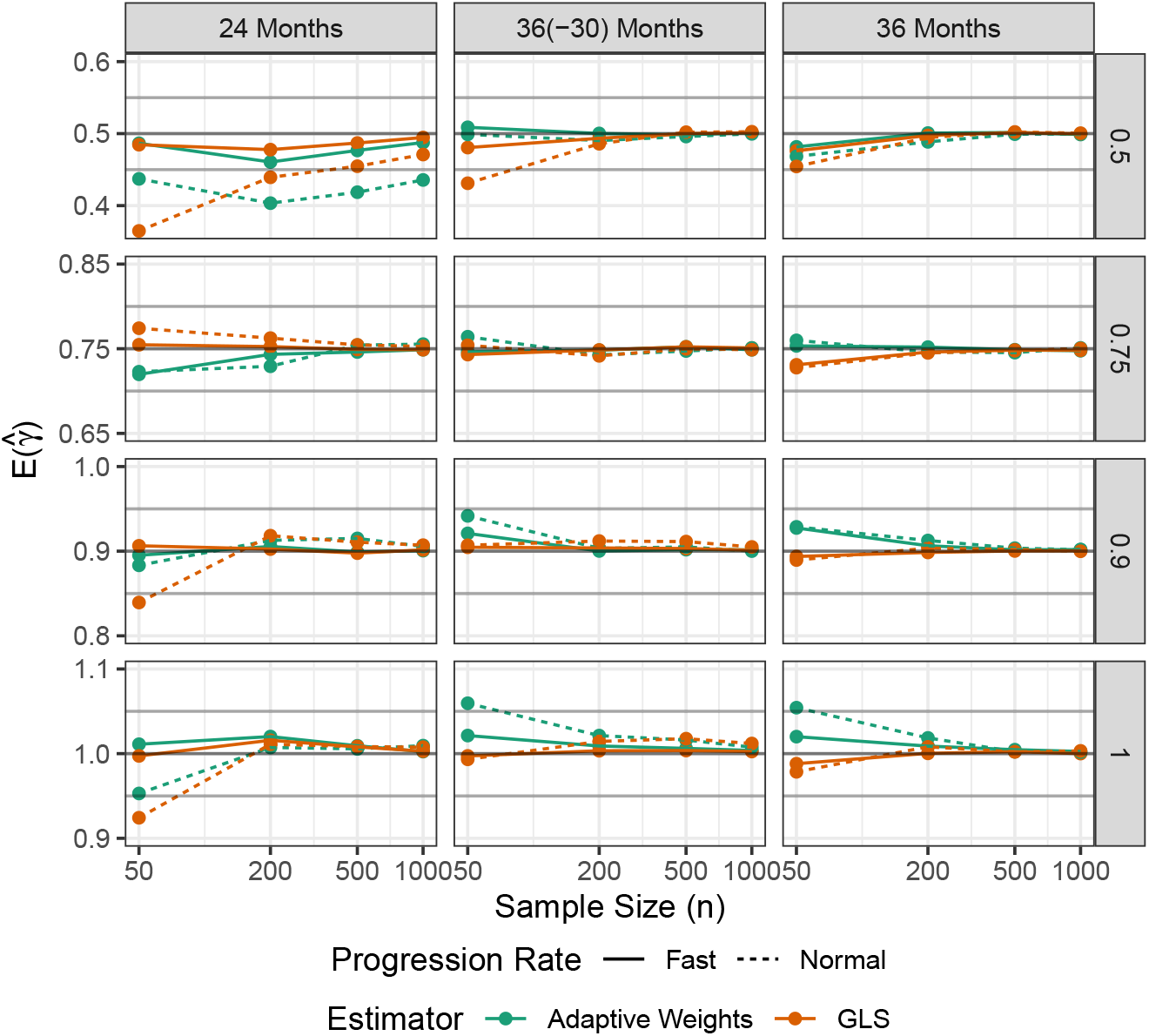
The means of the estimated common acceleration factors as functions of the sample size. The rows correspond to the true common acceleration factors, the columns correspond to the measurement patterns. In each subplot, a black horizontal line represents the true acceleration factor and gray horizontal lines represent the 0.05-margin around the true value. GLS: generalized least squares.

Figure 3 also shows that, when there is bias, the bias is smaller (i) for the fast progression rate and (ii) when there are more measurement occasions. Anyhow, the bias is mostly smaller than 0.05 when Assumption 6 is satisfied. This assumption is not testable, however.

The empirical coverage rates of the 95% CIs (based on inverting hypothesis tests) are plotted in Figure 4. This reveals that there is close to nominal coverage for the adaptive weights-based estimator across all scenarios. For the GLS estimator, there is undercoverage for the sample sizes between 50 and 500, but coverage is nominal or close to nominal for *n* = 1000.

**FIGURE 4.**
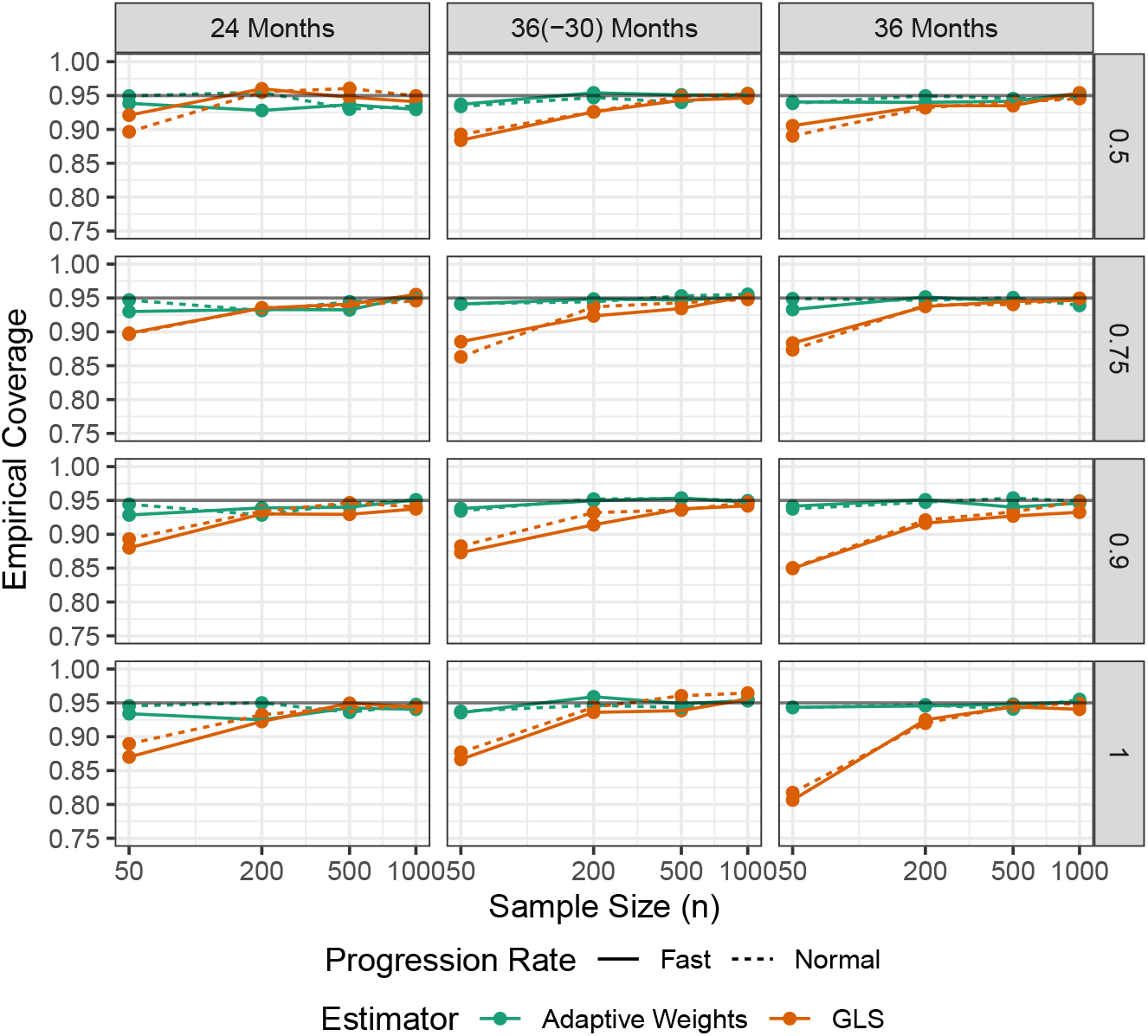
The empirical coverage rates of the confidence intervals (based on inverting hypothesis tests) as functions of the sample size. The rows correspond to the true acceleration factors, the columns correspond to the measurement patterns. The black horizontal lines indicate 95% coverage. GLS: generalized least squares.

In Figure 5, the empirical coverage rates are presented for the 95% percentile CIs based on the parametric bootstrap. To limit the computational burden, we only used *B* = 500 bootstrap replications throughout and only reanalyzed the normal progression scenarios (because the largest deviations from nominal were observed there). Figure 5 reveals close-to-nominal coverage across all scenarios for the adaptive weights-based estimator and an overcoverage that disappears with an increasing sample size for the GLS estimator. These results indicate that the parametric bootstrap permits valid, but sometimes conservative, inferences in small samples.

**FIGURE 5.**
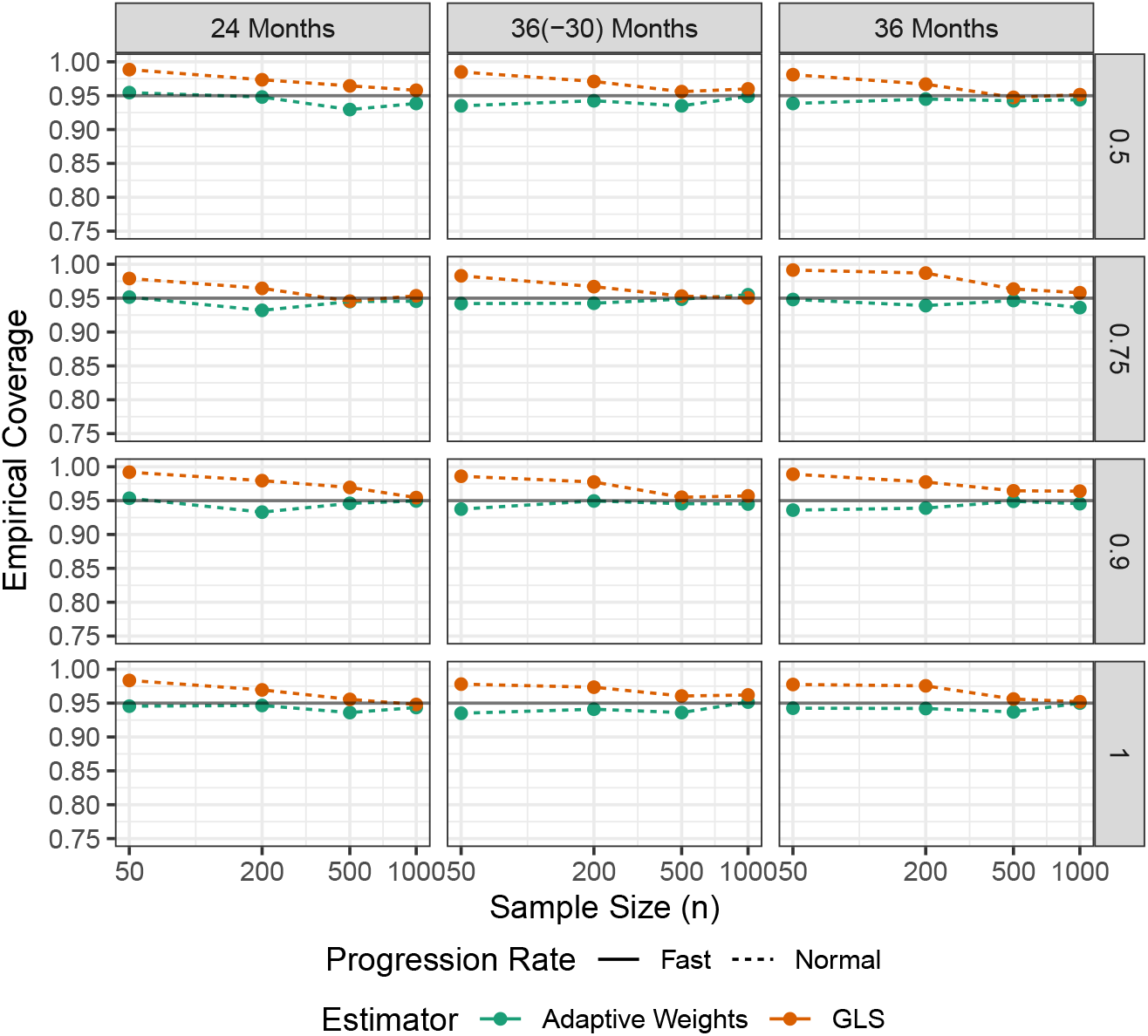
The empirical coverage rates of the percentile confidence intervals based on the parametric bootstrap as functions of the sample size. The rows correspond to the true acceleration factors, the columns correspond to the measurement patterns. The black horizontal lines indicate 95% coverage. GLS: generalized least squares.

The remaining simulation results presented in Appendix D reveal that tests based on the common acceleration factor do not increase power (as compared to the standard F-test in a MMRM) except in a few settings. Additionally, the estimator based on linear interpolation remains slightly biased (which is expected as consistency is not guaranteed when Assumption 6 is violated).

## 5 DATA APPLICATION

### 5.1 Motivating Data

The motivating data come from a multi-center phase 2/3 clinical trial (NCT03823404) in AD where a potentially DMT was compared to placebo ^17^. Our analysis of these data focused on a pre-specified subgroup of patients in which, based on the mechanism of action, it was thought the drug would be most effective. Within this subgroup, 148 patients were randomized to active treatment and 79 to placebo. The primary outcome measure is the ADAS-Cog scale measured at baseline and at 12, 24, 40, and 48 weeks after randomization.

### 5.2 Mixed Model for Repeated Measures

The type of data described above are usually analyzed with a MMRM, typically adjusted for baseline covariates. We fitted a MMRM using REML to the motivating data where the vector of ADAS-Cog scores at baseline, 12, 24, 40, and 48 weeks after randomization, ***Y*** = (*Y*_0_, *Y*_12_, *Y*_24_, *Y*_40_, *Y*_48_), is the response variable and the covariance matrix is unstructured but common to all patients. We also added an interaction between time and ApoE4 status, and a main effect for the study site. The latter is necessary to maintain the independent observations assumption. More specifically, the following parametric model is assumed for these data:

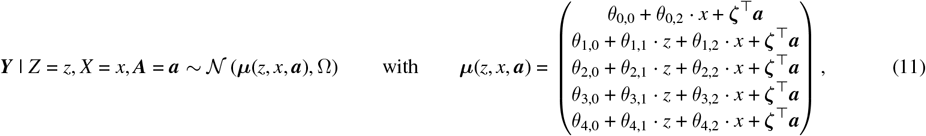

where *x* is an indicator for ApoeE4 status and ***a*** is a vector of dummy variables for the study site. Note that the mean function ***µ***(*x, z*, ***a***) does not allow for a treatment effect at baseline; this holds by randomization at baseline.

The null hypothesis of no treatment effect, *H*_0_ : *θ*_1,1_ = *θ*_2,1_ = *θ*_3,1_ = *θ*_4,1_ = 0, is tested using the *F*-test with the Kenward-Roger degrees-of-freedom approximation, which results in *p* = 0.015. Thus, there is a significant treatment effect, but it is unclear how relevant this effect really is.

A first step towards interpreting the magnitude of the treatment effect is looking at the estimated mean differences in ADAS-Cog scores. Because we have included baseline covariates in the MMRM, we have to carefully define the treatment group-specific mean ADAS-Cog scores at the various measurement occasions. In what follows, we consider the estimated marginal means (EMMs) where the centers and the ApoE4 levels receive weights proportional to their observed frequencies in the data. These are valid estimators for the potential-outcome means, *E*(*Y*_*t*_(*z*)), if the parametric assumptions hold.

The EMMs with proportional weights and the corresponding contrasts by measurement occasion are estimated with emmeans() and contrast() from the *emmeans* R package ^18^ and are presented in Table 1. These contrasts indicate that the treatment effect increases with time, but it is difficult to appreciate the relevance of these effects: is a 2 or 3 points mean difference on the ADAS-Cog scale meaningful? This question cannot be answered without also taking into account the duration of treatment and the disease severity (as minimal clinically important differences also depend on these factors) ^19^.

**TABLE 1.** Estimated marginal means and the corresponding treatment contrasts by measurement occasion, and the corresponding estimated time-specific acceleration factors. The penultimate column contains the *t*-statistic for testing *H*_0_ : *β*_*j*,0_ = *α*_*j*,0_; this corresponds to the *z*-statistic for *H*_0_ : *γ*_*j*,0_ = 1 as explained in Section 3.1.2. 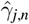 : estimated time-specific acceleration factor; CI: confidence interval.

| Measurement Occasion | Active Treatment (95% CI)<sup>a</sup> | Placebo (95% CI)<sup>a</sup> | Contrast (95% CI)<sup>a</sup> | $t$-statistic (df) | $\hat{\gamma}_{j,n}$ (95% CI)<sup>b</sup> |
| --- | --- | --- | --- | --- | --- |
| Baseline | - | 23.2 (22.3, 24.2) | - | - | - |
| Week 12 | 23.4 (22.2, 24.6) | 24.1 (22.7, 25.5) | -0.68 (-1.94, 0.59) | -1.06 (217) | 0.32 (-0.39, 1.37) |
| Week 24 | 24.5 (23.1, 25.9) | 26.0 (24.4, 27.6) | -1.43 (-2.91, 0.06) | -1.89 (208) | 0.65 (0.19, 1.01) |
| Week 40 | 25.4 (24.0, 26.9) | 28.2 (26.5, 29.9) | -2.76 (-4.36, -1.16) | -3.41 (201) | 0.52 (0.29, 0.76) |
| Week 48 | 26.4 (24.7, 28.2) | 29.0 (26.9, 31.0) | -2.54 (-4.60, -0.48) | -2.43 (184) | 0.56 (0.35, 0.83) |

<sup>a</sup> based on the $t$ -distribution with Kenward-Roger approximate degrees of freedom
<sup>b</sup> based on inverting the contrast-based tests

### 5.3 Acceleration Factors

We now use the EMMs from the previous section to estimate the acceleration factors using the *TCT* R package. First, we extract the EMMs at all time-treatment combinations, 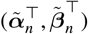. We also need the corresponding estimated covariance matrix, 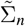. The emmeans() function returns these objects. Second, we use the TCT_meta() function to estimate the time-specific acceleration factors; these estimates are presented in the last column of Table 1. The estimated time-specific acceleration factors do not contradict proportional slowing: the CIs overlap considerably. We additionally performed the test for proportional slowing based on the minimized GLS criterion, which was not significant (*p* = 0.600). A visual check for proportional slowing also provides no evidence against proportional slowing (see Appendix C.2).

We use the adaptive weights-based and GLS estimators for the common acceleration factor. Because the sample size is relatively small, we further use the parametric bootstrap (*B* = 2000) with percentile intervals for inference. The adaptive weights-based estimate (95% CI) is 0.54 (0.33, 0.80), the GLS estimate (95% CI) is 0.54 (0.36, 0.90). Both estimates correspond to a slowing of disease progression with 50%—a very relevant effect. For completeness, the 95% CIs based on inverting hypothesis tests are (0.32, 0.77) and (0.36, 0.77) for tests based on the adaptively weighted contrasts and minimized GLS criterion, respectively.

## 6 DISCUSSION AND CONCLUSIONS

In this paper, we introduced a class of causal target parameters that quantify treatment effects on the time scale together with a set of identifying assumptions. We further focused on the time-specific and common acceleration factors as target parameters and developed two-step estimators that only require summary-level information. We theoretically studied the asymptotic behavior of these estimators and corresponding tests, and studied finite-sample behavior in a simulation study based on the ADNI data. These methods were further applied to data from a phase 2/3 clinical trial in AD, revealing a significant and clinically relevant treatment effect. In contrast, the analysis with an MMRM revealed a statistically significant effect, but left the question of clinical relevance unaddressed.

The methods presented in this paper complement the methods in Raket ^4^. The latter rely on non-linear parametric models— these can only be fitted with individual-patient data, rely on parametric assumptions, and are specific to the type of data (e.g., extending these methods from continuous to binary outcomes is certainly not trivial). The target parameters defined in this paper are causal and model free; therefore, they remain well defined for observational data under a non-parametric model for the time-specific acceleration factors and semi-parametric model for the common acceleration factor. Hence, as long as one can estimate *E*(*Y*_*t*_(*z*)) for *t* ∈ {0, *t*_1_, …, *t*_*K*_} and *z* ∈ {0, 1} (e.g., using non-parametric methods like targeted maximum likelihood ^20^ and debiased machine learning ^21^), our methods can be applied to estimate the acceleration factors. In a randomized trial, other methods for longitudinal data like generalized estimating equations ^22^ or longitudinal quantile regression ^23^ could be used instead of a MMRM with no modification to how we estimate and draw inferences about the acceleration factors.

The simulation study showed that inference is difficult for small to medium sample sizes, especially if progression is slow and measurements are sparse. Raket ^4^ encountered similar issues with their non-linear regression models for individual-patient data. Our parametric bootstrap solves some of these issues at the cost of being conservative in small samples. Note, however, that the finite-sample behavior of our two-step estimators strongly depends on the finite-sample behavior of 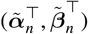 and 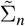. If the latter estimators converge very slowly to the normal limiting distribution and to Σ_0_, respectively, then this will translate to poor finite-sample behavior of the proposed two-step estimators.

The common acceleration factor and our proposed two-step estimators are especially useful for meta-analysis for two reasons. First, the two-step estimators only requires summary-level information (i.e., 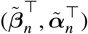 and 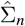 as defined in the text). The estimated covariance matrix 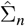 is, however, usually not reported in publications and cooperation from the owners of the data would, therefore, still be required as they have to supply this matrix. Second, the common acceleration factor is an interpretable target parameter whose scale is shared across trials with different designs (e.g., different timing of measurements or different clinical scales). It may, therefore, be a useful target parameter to compare across trials.

## Supporting information

Web Appendices

## Data Availability

The data described cannot be made available, but
the GitHub repo contains a simulated mock dataset based on the case study (and code to
analyze these mock data).

https://www.github.com/florianstijven/TCT

https://www.github.com/florianstijven/meta-TCT-simulations

## ACKNOWLEDGMENTS

We thank Lighthouse Pharma for allowing us to use their data in this paper.

The resources and services used in this work were provided by the VSC (Flemish Supercomputer Center), funded by the Research Foundation – Flanders (FWO) and the Flemish Government.

## SUPPORTING INFORMATION

The *TCT* R package is available from github.com/florianstijven/TCT. The code used for the simulations and the data application are available from github.com/florianstijven/meta-TCT-simulations. The data described in Section 5 cannot be made available, but the GitHub repo contains a simulated mock dataset based on the case study (and code to analyze this mock dataset).

The Appendix referenced throughout the paper is available online.

