## Supplementary material for "Time-Scale Target Parameters and Two-Step Estimation in Longitudinal Trials for Progressive Diseases": Web Appendices

### A Technical Results

This appendix contains technical results that support the estimators and inferential procedures proposed on the main text. We first introduce some preliminary theory and assumptions that are used in the second and third part of this appendix, where the asymptotics of, respectively, the contrast-based and GLS estimators are studied.

#### A.1 Preliminaries

We first introduce a general theorem that is used multiple times for proving asymptotic results later on. Next, we discuss some regularity conditions on the inter- and extrapolation methods used in practice.

##### A.1.1 General Theory

The following theorem contains a general result that will be used in the proofs of the lemmas about the estimators described in the main text. This lemma is essentially a variation on the delta method where the "differentiable function" is defined implicitly as the root of an estimating function that is indexed by (i) an asymptotically normal random variable and (ii) a nuisance parameter that converges to a fixed element at an arbitrarily slow rate.

**Theorem A.1.** *Let $\boldsymbol{\theta} \in \Theta \subseteq \mathbb{R}^p \mapsto \boldsymbol{\Phi}(\boldsymbol{\theta}; \boldsymbol{y}, \eta) \in \mathbb{R}^p$ be an estimating function, indexed by $\boldsymbol{y} \in \mathbb{R}^d$ and $\eta \in \Gamma$. Assume that $\boldsymbol{\Phi}(\boldsymbol{\theta}; \boldsymbol{y_0}, \eta_0) = \boldsymbol{0}$ for some $\boldsymbol{\theta_0}$ in the interior of $\Theta$, $\boldsymbol{y_0} \in \mathbb{R}^d$, and $\eta_0 \in \Gamma$. Further assume the following conditions:*

*(i)* $\{\boldsymbol{y}_n\}$ *is a sequence of random variables such that* $n^{1/2}(\boldsymbol{y}_n - \boldsymbol{y_0}) \xrightarrow{d} \mathcal{N}(\boldsymbol{0}, \Sigma_0)$

*(ii)* $\|\boldsymbol{\Phi}(\boldsymbol{\theta}_n; \boldsymbol{y_0}, \eta_0)\|_2 \to 0$ *implies that* $\boldsymbol{\theta}_n \to \boldsymbol{\theta_0}$ *and* $\|\boldsymbol{\Phi}(\boldsymbol{\theta_0}; \boldsymbol{y_0}, \eta)\|_2 = 0$ *for* $\eta$ *in a neighborhood of* $\eta_0$.

*(iii)* $\boldsymbol{\theta} \mapsto \boldsymbol{\Phi}(\boldsymbol{\theta}; \boldsymbol{y}, \eta)$ *is continuously differentiable on* $\Theta$ *for all* $\boldsymbol{y}$ *and* $\eta$ *in a neighborhood of* $\boldsymbol{y_0}$ *and* $\eta_0$*, respectively. The corresponding matrix of partial derivatives is denoted by* $\dot{\boldsymbol{\Phi}}_{\boldsymbol{\theta}}(\boldsymbol{\theta}; \boldsymbol{y}, \eta)$.

*(iv)* $\boldsymbol{y} \to \boldsymbol{\Phi}(\boldsymbol{\theta_0}; \boldsymbol{y}, \eta)$ *is continuously differentiable in a neighborhood of* $\boldsymbol{y_0}$ *for all* $\eta$ *in a neighborhood around* $\eta_0$*. The corresponding matrix of partial derivatives is denoted by* $\dot{\boldsymbol{\Phi}}_{\boldsymbol{y}}(\boldsymbol{\theta_0}; \boldsymbol{y}, \eta)$.

*(v)* $(\boldsymbol{\theta}, \boldsymbol{y}, \eta) \to \dot{\boldsymbol{\Phi}}_{\boldsymbol{\theta}}(\boldsymbol{\theta}; \boldsymbol{y}, \eta)$ *is continuous at* $(\boldsymbol{\theta_0}, \boldsymbol{y_0}, \eta_0)$*,* $\dot{\boldsymbol{\Phi}}_{\boldsymbol{\theta}}(\boldsymbol{\theta}_0; \boldsymbol{y_0}, \eta_0)$ *is invertible, and* $(\boldsymbol{y}, \eta) \mapsto \dot{\boldsymbol{\Phi}}_{\boldsymbol{y}}(\boldsymbol{\theta_0}; \boldsymbol{y}, \eta)$ *is continuous at* $(\boldsymbol{y_0}, \eta_0)$.

*(vi) If* $\boldsymbol{y} \to \boldsymbol{y_0}$ *and* $\eta \to \eta_0$*, then* $\sup_{\boldsymbol{\theta} \in \Theta} \|\boldsymbol{\Phi}(\boldsymbol{\theta}; \boldsymbol{y}, \eta) - \boldsymbol{\Phi}(\boldsymbol{\theta}; \boldsymbol{y_0}, \eta_0)\|_2 \to 0$.

*(vii)* $\{\eta_n\}$ *is a sequence of random elements such that* $\eta_n \xrightarrow{P} \eta_0$

*Let $\{\hat{\boldsymbol{\theta}}_n\}$ be a sequence of random variables for which holds that $\boldsymbol{\Phi}(\hat{\boldsymbol{\theta}}_n; \boldsymbol{y}_n, \eta_n) = o_P(n^{-1/2})$. Under the above conditions, $\hat{\boldsymbol{\theta}}_n$ is consistent and asymptotically normal:*

$$n^{1/2}(\hat{\boldsymbol{\theta}}_n - \boldsymbol{\theta_0}) \xrightarrow{d} \mathcal{N}(\boldsymbol{0}, \Omega),$$

*where*

$$\Omega = \dot{\boldsymbol{\Phi}}_{\boldsymbol{\theta}}^{-1}(\boldsymbol{\theta_0}; \boldsymbol{y_0}, \eta_0) \cdot \dot{\boldsymbol{\Phi}}_{\boldsymbol{y}}(\boldsymbol{\theta_0}; \boldsymbol{y_0}, \eta_0) \cdot \Sigma_0 \cdot \dot{\boldsymbol{\Phi}}_{\boldsymbol{y}}(\boldsymbol{\theta_0}; \boldsymbol{y_0}, \eta_0)^\top \cdot \dot{\boldsymbol{\Phi}}_{\boldsymbol{\theta}}^{-1}(\boldsymbol{\theta_0}; \boldsymbol{y_0}, \eta_0).$$

*Proof.* Because $\boldsymbol{y}_n \overset{P}{\to} \boldsymbol{y_0}$ and $\eta_n \overset{P}{\to} \eta_0$ by, respectively, conditions (i) and (vii), $\boldsymbol{y}_n$ and $\eta_n$ are in any neighborhood of $\boldsymbol{y_0}$ and $\eta_0$, respectively, with probability tending to one. In the following derivations, we assume that $\boldsymbol{y}_n$ and $\eta_n$ are in all neighborhoods mentioned in the Theorem's conditions.

**Consistency.** By arguing along subsequences (Kosorok, 2008, Lemma 7.21(ii)), condition (ii) implies that $\hat{\boldsymbol{\theta}}_n$ is consistent if $\|\boldsymbol{\Phi}(\hat{\boldsymbol{\theta}}_n; \boldsymbol{y_0}, \eta_0)\|_2 = o_P(1)$ where $\|\cdot\|_2$ is the Euclidean norm. We now show that $\|\boldsymbol{\Phi}(\hat{\boldsymbol{\theta}}_n; \boldsymbol{y_0}, \eta_0)\|_2 = o_P(1)$:

$$
\begin{aligned}
\|\boldsymbol{\Phi}(\hat{\boldsymbol{\theta}}_n; \boldsymbol{y_0}, \eta_0)\|_2 &\le \|\boldsymbol{\Phi}(\hat{\boldsymbol{\theta}}_n; \boldsymbol{y_0}, \eta_0) - \boldsymbol{\Phi}(\hat{\boldsymbol{\theta}}_n; \boldsymbol{y}_n, \eta_n)\|_2 + \|\boldsymbol{\Phi}(\hat{\boldsymbol{\theta}}_n; \boldsymbol{y}_n, \eta_n)\|_2 && \text{(triangle inequality)} \\
&\le \sup_{\boldsymbol{\theta}\in\Theta} \|\boldsymbol{\Phi}(\boldsymbol{\theta}; \boldsymbol{y_0}, \eta_0) - \boldsymbol{\Phi}(\boldsymbol{\theta}; \boldsymbol{y}_n, \eta_n)\|_2 + o_P(1) && \text{(definition of } \hat{\boldsymbol{\theta}}_n) \\
&\le o_P(1) && \text{(condition (vi))}
\end{aligned}
$$

For the last inequality, we can again argue along subsequences.

**Asymptotic Normality.** By condition (iii) and $\boldsymbol{\theta_0}$ being an interior point of $\Theta$, we can apply the mean value theorem to each element of $\boldsymbol{\theta} \mapsto \boldsymbol{\Phi}(\boldsymbol{\theta}; \boldsymbol{y}_n, \eta_n)$ where $\tilde{\boldsymbol{\theta}}_n$ is an intermediate value between $\hat{\boldsymbol{\theta}}_n$ and $\boldsymbol{\theta_0}$. Because $\hat{\boldsymbol{\theta}}_n$ is consistent and $\tilde{\boldsymbol{\theta}}_n$ is an intermediate value, $\tilde{\boldsymbol{\theta}}_n$ is also consistent. Note that the intermediate value may be different for each element of the vector-valued function $\boldsymbol{\Phi}$, but we ignore this in the notation.

$$
\begin{aligned}
\boldsymbol{\Phi}(\boldsymbol{\theta_0}; \boldsymbol{y}_n, \eta_n) &= -\dot{\boldsymbol{\Phi}}_{\boldsymbol{\theta}}(\tilde{\boldsymbol{\theta}}_n; \boldsymbol{y}_n, \eta_n) \cdot (\hat{\boldsymbol{\theta}}_n - \boldsymbol{\theta}_0) + \boldsymbol{\Phi}(\hat{\boldsymbol{\theta}}_n; \boldsymbol{y}_n, \eta_n) \\
&= -\dot{\boldsymbol{\Phi}}_{\boldsymbol{\theta}}(\tilde{\boldsymbol{\theta}}_n; \boldsymbol{y}_n, \eta_n) \cdot (\hat{\boldsymbol{\theta}}_n - \boldsymbol{\theta}_0) + o_P(n^{-1/2}) && \text{(definition of } \hat{\boldsymbol{\theta}}_n) \\
&= -\Big(\dot{\boldsymbol{\Phi}}_{\boldsymbol{\theta}}(\boldsymbol{\theta_0}; \boldsymbol{y_0}, \eta_0) + o_P(1)\Big) \cdot (\hat{\boldsymbol{\theta}}_n - \boldsymbol{\theta}_0) + o_P(n^{-1/2})
\end{aligned}
$$

The third equality follows from condition (v) and the consistency of $\tilde{\boldsymbol{\theta}}_n$, $\boldsymbol{y}_n$, and $\eta_n$ (together with the continuous mapping theorem).

By condition (iv), we can apply the mean value theorem to $\boldsymbol{\Phi}(\boldsymbol{\theta_0}; \boldsymbol{y}_n, \eta_n)$ where $\tilde{\boldsymbol{y}}_n$ is an intermediate value between $\boldsymbol{y}_n$ and $\boldsymbol{y_0}$. Because $\boldsymbol{y}_n$ is consistent and $\tilde{\boldsymbol{y}}_n$ is an intermediate value, $\tilde{\boldsymbol{y}}_n$ is also consistent. Note that the intermediate value may be different for each element of the vector-valued function $\boldsymbol{\Phi}$, but we ignore this in the notation.

$$
\begin{aligned}
n^{1/2} \cdot \boldsymbol{\Phi}(\boldsymbol{\theta_0}; \boldsymbol{y}_n, \eta_n) &= n^{1/2} \cdot \dot{\boldsymbol{\Phi}}_{\boldsymbol{y}}(\boldsymbol{\theta_0}; \tilde{\boldsymbol{y}}_n, \eta_n) \cdot (\boldsymbol{y}_n - \boldsymbol{y_0}) + \boldsymbol{\Phi}(\boldsymbol{\theta_0}; \boldsymbol{y_0}, \eta_n) \\
&= \Big\{ n^{1/2} \cdot \dot{\boldsymbol{\Phi}}_{\boldsymbol{y}}(\boldsymbol{\theta_0}; \boldsymbol{y_0}, \eta_0) + o_P(1) \Big\} \cdot (\boldsymbol{y}_n - \boldsymbol{y_0})
\end{aligned}
$$

where $\|\boldsymbol{\Phi}(\boldsymbol{\theta_0}; \boldsymbol{y_0}, \eta_n)\|_2 = 0$ by condition (ii). We also used condition (v) and the consistency of $\tilde{\boldsymbol{y}}_n$ and $\eta_n$ in the last equality (together with the continuous mapping theorem).

Hence, we have that $\boldsymbol{\Phi}(\boldsymbol{\theta_0}; \boldsymbol{y}_n, \eta_n) = O_P(n^{-1/2})$ which then implies that $n^{1/2}(\hat{\boldsymbol{\theta}}_n - \boldsymbol{\theta_0}) = O_P(1)$. It now follows that

$$
\begin{aligned}
n^{1/2}(\hat{\boldsymbol{\theta}}_n - \boldsymbol{\theta_0}) &= -n^{1/2} \cdot \dot{\boldsymbol{\Phi}}_{\boldsymbol{\theta}}^{-1}(\boldsymbol{\theta_0}; \boldsymbol{y_0}, \eta_0) \cdot \boldsymbol{\Phi}(\boldsymbol{\theta_0}; \boldsymbol{y}_n, \eta_n) + o_P(1) \\
&= -n^{1/2} \cdot \dot{\boldsymbol{\Phi}}_{\boldsymbol{\theta}}^{-1}(\boldsymbol{\theta_0}; \boldsymbol{y_0}, \eta_0) \cdot \dot{\boldsymbol{\Phi}}_{\boldsymbol{y}}(\boldsymbol{\theta_0}; \boldsymbol{y_0}, \eta_0) \cdot (\boldsymbol{y}_n - \boldsymbol{y_0}) + o_P(1).
\end{aligned}
$$

From condition (i) now follows that $n^{1/2}(\hat{\boldsymbol{\theta}}_n - \boldsymbol{\theta_0})$ converges to a mean-zero normal distribution with variance matrix

$$
\Omega = \dot{\boldsymbol{\Phi}}_{\boldsymbol{\theta}}^{-1}(\boldsymbol{\theta_0}; \boldsymbol{y_0}, \eta_0) \cdot \dot{\boldsymbol{\Phi}}_{\boldsymbol{y}}(\boldsymbol{\theta_0}; \boldsymbol{y_0}, \eta_0) \cdot \Sigma_0 \cdot \dot{\boldsymbol{\Phi}}_{\boldsymbol{y}}(\boldsymbol{\theta_0}; \boldsymbol{y_0}, \eta_0)^\top \cdot \dot{\boldsymbol{\Phi}}_{\boldsymbol{\theta}}^{-1}(\boldsymbol{\theta_0}; \boldsymbol{y_0}, \eta_0).
$$

□

##### A.1.2 Interpolation and Extrapolation

For the estimators described in the main text to work, we need to make regularity assumptions about the interpolation method mentioned in Assumption 6 and the extrapolation method used in the implementation[1]. These regularity assumptions[2] hold for most reasonable interpolation and extrapolation methods.

**Assumption A.1** (Regular interpolation and extrapolation method)**.** *Let* $0 < t_1 < \cdots < t_K$ *be a set of finite values and* $\boldsymbol{y} = (y_0, y_1, \ldots, y_1)' \in \mathbb{R}^{K+1}$*. Let* $t \in \mathbb{R} \mapsto f(t; \boldsymbol{y})$ *be an interpolating function between* $\{(0, y_0), (t_1, y_1), \ldots, (t_K, y_K)\}$ *extended to the real line by some extrapolation method. This function satisfies the following properties, for some* $\delta > 0$ *not depending on* $\boldsymbol{y}$*:*

*(i) It is continuously differentiable for* $t \in [0, t_K + \delta]$*.*

*(ii) For any* $\boldsymbol{y} \in \mathbb{R}^{K+1}$ *and* $\boldsymbol{y}_n \to \boldsymbol{y}$*, it holds that*

$$\sup_{t \in [0, t_K + \delta]} |f(t; \boldsymbol{y}_n) - f(t; \boldsymbol{y})| \to 0 \quad \text{and} \quad \sup_{t \in [0, t_K + \delta]} \left| \frac{\partial f(t; \boldsymbol{y}_n)}{\partial t} - \frac{\partial f(t; \boldsymbol{y})}{\partial t} \right| \to 0.$$

*(iii)* $\boldsymbol{y} \mapsto f(t; \boldsymbol{y})$ *is continuously differentiable for all* $t \in [0, t_K]$*.*

*(iv) If* $f(\cdot; \boldsymbol{y}) : [0, t_K] \to \mathbb{R}$ *is strictly monotone, then* $f(\cdot; \boldsymbol{y}) : [0, t_K + \delta] \to \mathbb{R}$ *is also strictly monotone.*

#### A.2 Estimation and Inference Based on Contrast Functions

We first define the contrast functions, derive the partial derivatives required in Theorem A.1, and prove a technical lemma related to these contrast functions and their partial derivatives. We then study the asymptotics of the estimators based on the contrast functions:

1. contrast-based estimator of the time-specific acceleration factors
2. weights-based estimator of the common acceleration factor
3. adaptive weights-based estimator of the common acceleration factor.

##### A.2.1 Contrast Functions

The contrast function was defined in the main text as

$$\boldsymbol{\Phi}(\boldsymbol{\gamma}; \hat{\boldsymbol{\alpha}}_n, \hat{\boldsymbol{\beta}}_n) = \begin{pmatrix} \hat{\beta}_{1,n} - f_0(\gamma_1 \cdot t_1; \hat{\boldsymbol{\alpha}}_n) \\ \vdots \\ \hat{\beta}_{K,n} - f_0(\gamma_K \cdot t_K; \hat{\boldsymbol{\alpha}}_n) \end{pmatrix} = \hat{\boldsymbol{\beta}}_n - \boldsymbol{f_0}(\boldsymbol{\gamma t}; \hat{\boldsymbol{\alpha}}_n), \tag{1}$$

where $\boldsymbol{\gamma} = (\gamma_1, \ldots, \gamma_K)^\top$ and $\boldsymbol{t} = (t_1, \ldots, t_K)^\top$. This notation is very similar to the notation in Theorem A.1. This is on purpose as $\boldsymbol{\Phi}$ defined above is an estimating function for $\boldsymbol{\gamma}$ (which is $\boldsymbol{\theta}$ in

[1] Extrapolation is not necessary for identification (see Section 2 of the main text); however, extrapolation is needed in any implementation because $\max_j \hat{\beta}_j > \max_j \hat{\alpha}_j$ and $\min_j \hat{\beta}_j < \min_j \hat{\alpha}_j$ may occur in finite samples. The extrapolation method only affects the asymptotic distribution of the estimators if $\gamma_0 = 1$.

[2] The regularity assumptions listed here are actually much stronger than required. However, weakening these assumptions would make them more difficult to understand.

Theorem A.1) indexed by $(\hat{\boldsymbol{\alpha}}_n, \hat{\boldsymbol{\beta}}_n)'$ (which is $\boldsymbol{y}_n$ in Theorem A.1). (There is no nuisance parameter $\eta$ here.)

The corresponding matrices of partial derivatives mentioned in Theorem A.1 are as follows:

$$\dot{\boldsymbol{\Phi}}_{\boldsymbol{\gamma}}(\boldsymbol{\gamma}; \boldsymbol{\alpha}, \boldsymbol{\beta}) = \begin{pmatrix} t_1 \cdot \frac{\partial f_0(t;\boldsymbol{\alpha})}{\partial t}\Big|_{t=\gamma_1 \cdot t_1} & \cdots & 0 \\ \vdots & \ddots & \vdots \\ 0 & \cdots & t_K \cdot \frac{\partial f_0(t;\boldsymbol{\alpha})}{\partial t}\Big|_{t=\gamma_K \cdot t_K} \end{pmatrix}$$

and

$$\dot{\boldsymbol{\Phi}}_{\boldsymbol{\alpha},\boldsymbol{\beta}}(\boldsymbol{\gamma}; \boldsymbol{\alpha}, \boldsymbol{\beta}) = \begin{pmatrix} A & I_K \end{pmatrix} \quad \text{where} \quad A = \begin{pmatrix} -\frac{\partial f_0}{\partial \alpha_0}(\gamma \cdot t_1; \boldsymbol{\alpha}) & \dots & -\frac{\partial f_0}{\partial \alpha_K}(\gamma \cdot t_1; \boldsymbol{\alpha}) \\ \vdots & \ddots & \vdots \\ -\frac{\partial f_0}{\partial \alpha_0}(\gamma \cdot t_K; \boldsymbol{\alpha}) & \dots & -\frac{\partial f_0}{\partial \alpha_K}(\gamma \cdot t_K; \boldsymbol{\alpha}) \end{pmatrix},$$

and $I_K$ is the $(K \times K)$ identity matrix.

In the proofs of some of the following lemmas, we use the following technical result.

**Lemma A.2.** *Under Assumptions 1, 2, and A.1, conditions (iii–vi) of Theorem A.1 are satisfied for $\boldsymbol{\Phi}(\boldsymbol{\gamma}; \boldsymbol{\alpha}, \boldsymbol{\beta})$ defined in (1) where $\boldsymbol{\theta}$ corresponds to $\boldsymbol{\gamma}$, and $(\boldsymbol{\alpha}^\top, \boldsymbol{\beta}^\top)^\top$ corresponds to $\boldsymbol{y}$. We let $\Theta \subseteq [0, 1 + \delta/t_K]$. There is no $\eta$.*

*Proof.* **Condition (iii) and first part of condition (v).** $\dot{\boldsymbol{\Phi}}_{\boldsymbol{\gamma}}(\boldsymbol{\gamma}; \boldsymbol{\alpha}; \boldsymbol{\beta})$ is diagonal and only contains entries of the form $t_j \cdot \frac{\partial f_0(t;\boldsymbol{\alpha})}{\partial t}\big|_{t=\gamma_j \cdot t_j}$. By Assumption A.1, $t \mapsto \frac{\partial f_0(t;\boldsymbol{\alpha})}{\partial t}$ is continuous; hence, the entries of $\dot{\boldsymbol{\Phi}}_{\boldsymbol{\gamma}}(\boldsymbol{\gamma}; \boldsymbol{\alpha}; \boldsymbol{\beta})$ are continuous in $\boldsymbol{\gamma}$ as well.

The first part of condition (v) follows if $\boldsymbol{\alpha}_n \to \boldsymbol{\alpha}_{\mathbf{0}}$ and $t_n \to t$ implies that $\frac{\partial f_0(t;\boldsymbol{\alpha}_n)}{\partial t}(t_n) \to \frac{\partial f_0(t;\boldsymbol{\alpha}_{\mathbf{0}})}{\partial t}(t)$ for all $t \in [0, t_K + \delta]$. For a given $t \in [0, t_K + \delta]$, we have that

$$\left|\frac{\partial f_0(t;\boldsymbol{\alpha}_n)}{\partial t}(t_n) - \frac{\partial f_0(t;\boldsymbol{\alpha}_{\mathbf{0}})}{\partial t}(t)\right| \leq \left|\frac{\partial f_0(t;\boldsymbol{\alpha}_n)}{\partial t}(t_n) - \frac{\partial f_0(t;\boldsymbol{\alpha}_{\mathbf{0}})}{\partial t}(t_n)\right| + \left|\frac{\partial f_0(t;\boldsymbol{\alpha}_{\mathbf{0}})}{\partial t}(t_n) - \frac{\partial f_0(t;\boldsymbol{\alpha}_{\mathbf{0}})}{\partial t}(t)\right|$$

The first part is $o(1)$ because of condition (ii) in Assumption A.1 and the second part is $o(1)$ because $t \mapsto f(t; \boldsymbol{\alpha}_{\mathbf{0}})$ is continuously differentiable (condition (i) of Assumption A.1) and, hence, continuous.

Further, $\dot{\boldsymbol{\Phi}}_{\boldsymbol{\gamma}}(\boldsymbol{\gamma}; \boldsymbol{\alpha}; \boldsymbol{\beta})$ is invertible if $\frac{\partial f_0(t;\boldsymbol{\alpha})}{\partial t}$ is non-zero for $t \in \{\gamma_{1,0} \cdot t_1, \dots, \gamma_{K,0} \cdot t_K\}$ and $\boldsymbol{\alpha} = \boldsymbol{\alpha}_{\mathbf{0}}$, which holds by Assumptions 1 and 2.

**Condition (iv) and second part condition (v).** $\dot{\boldsymbol{\Phi}}_{\boldsymbol{\alpha},\boldsymbol{\beta}}(\boldsymbol{\gamma}; \boldsymbol{\alpha}, \boldsymbol{\beta})$ only contains constants and entries of the form $\frac{\partial f_0(\gamma \cdot t_j;\boldsymbol{\alpha})}{\partial \alpha_{j'}}$. The latter entries are continuous in $\boldsymbol{\alpha}$ by Assumption A.1, condition (iii) and by Assumption 2 (which ensures that each element of $\boldsymbol{\gamma}_{\mathbf{0}}\boldsymbol{t}$ is in $[0, t_K]$).

Since there is no $\eta$, the second part of condition (v) follows from condition (iv).

**Condition (vi).** We can write the sup in condition (vi) as follows, where for $(\boldsymbol{\alpha}_n, \boldsymbol{\beta}_n) \to (\boldsymbol{\alpha}_{\mathbf{0}}, \boldsymbol{\beta}_{\mathbf{0}})$:

$$\begin{aligned}
\sup_{\boldsymbol{\gamma}\in\Theta} |\boldsymbol{\Phi}(\boldsymbol{\gamma}; \boldsymbol{\alpha}_n, \boldsymbol{\beta}_n) - \boldsymbol{\Phi}(\boldsymbol{\gamma}; \boldsymbol{\alpha}_{\mathbf{0}}, \boldsymbol{\beta}_{\mathbf{0}})\|_2 &= \sup_{\boldsymbol{\gamma}\in\Theta} \|\boldsymbol{\beta}_n - \boldsymbol{\beta}_{\mathbf{0}} + \boldsymbol{f}_{\mathbf{0}}(\boldsymbol{\gamma}\boldsymbol{t}; \boldsymbol{\alpha}_n) - \boldsymbol{f}_{\mathbf{0}}(\boldsymbol{\gamma}\boldsymbol{t};, \boldsymbol{\alpha}_{\mathbf{0}})\|_2 \\
&\leq \|\boldsymbol{\beta}_n - \boldsymbol{\beta}_{\mathbf{0}}\|_2 + \sup_{\boldsymbol{\gamma}\in\Theta} \|\boldsymbol{f}_{\mathbf{0}}(\boldsymbol{\gamma}\boldsymbol{t}; \boldsymbol{\alpha}_n) - \boldsymbol{f}_{\mathbf{0}}(\boldsymbol{\gamma}\boldsymbol{t}; \boldsymbol{\alpha}_{\mathbf{0}})\|_2 \quad \text{(triangle inequality)} \\
&\leq o(1) + K \cdot \sup_{t\in[0,t_K+\delta]} |f_0(t; \boldsymbol{\alpha}_n) - f_0(t; \boldsymbol{\alpha}_{\mathbf{0}})| \\
&= o(1)
\end{aligned}$$

The last equality follows from condition (ii) of Assumption A.1. □

##### A.2.2 Time-Specific Acceleration Factors

The following lemma contains the asymptotic distribution of the contrast-based estimator of the time-specific acceleration factors.

**Lemma A.3.** *Under Assumptions 1, 2, 6, 7, and A.1, the estimator for the time-specific acceleration factors, defined as the root*[3] *of*

$$\boldsymbol{\gamma} \in [0, 1 + \delta/t_K]^K \mapsto \boldsymbol{\Phi}(\boldsymbol{\gamma}; \hat{\boldsymbol{\alpha}}_n, \hat{\boldsymbol{\beta}}_n),$$

*and denoted by* $\hat{\boldsymbol{\gamma}}_n$*, is consistent and asymptotically normal:*

$$n^{1/2}(\hat{\boldsymbol{\gamma}}_n - \boldsymbol{\gamma_0}) \xrightarrow{d} \mathcal{N}(\mathbf{0}, \Omega_0),$$

*where*

$$\Omega = \dot{\boldsymbol{\Phi}}_{\boldsymbol{\gamma}}^{-1}(\boldsymbol{\gamma_0}; \boldsymbol{\alpha_0}, \boldsymbol{\beta_0}) \cdot \dot{\boldsymbol{\Phi}}_{\boldsymbol{\alpha},\boldsymbol{\beta}}(\boldsymbol{\gamma_0}; \boldsymbol{\alpha_0}, \boldsymbol{\beta_0}) \cdot \Sigma_0 \cdot \dot{\boldsymbol{\Phi}}_{\boldsymbol{\alpha},\boldsymbol{\beta}}(\boldsymbol{\gamma_0}; \boldsymbol{\alpha_0}, \boldsymbol{\beta_0})^\top \cdot \dot{\boldsymbol{\Phi}}_{\boldsymbol{\gamma}}^{-1}(\boldsymbol{\gamma_0}; \boldsymbol{\alpha_0}, \boldsymbol{\beta_0}).$$

*Proof.* We can apply Theorem A.1 (where there is no nuisance parameter $\eta$). Hence, we only need to verify conditions (i–vii) from Theorem A.1 to complete the proof.

**Condition (i).** This follows directly from Assumption 7.

**Condition (ii).** Assumptions 1 and A.1 imply that $t \mapsto f_0(t; \boldsymbol{\alpha_0})$ is strictly monotone and continuous on $[0, t_K + \delta]$, which in turn implies that $\gamma \mapsto \beta_{j,0} - f_0(\gamma \cdot t_j; \boldsymbol{\alpha_0})$ is strictly monotone and continuous on $[0, 1 + \delta/t_K]$. Hence, if this function has a root, it is unique. By Assumption 2, this function has a root, which we denote by $\gamma_{j,0}$. (This is the true time-specific acceleration factor by Assumption 6.) Hence, $\beta_{j,0} - f_0(\gamma_n \cdot t_j; \boldsymbol{\alpha_0}) \to 0$ implies that $\gamma_n \to \gamma_{j,0}$. This applies to each element of $\boldsymbol{\Phi}(\boldsymbol{\gamma}; \boldsymbol{\alpha_0}; \boldsymbol{\beta_0})$; hence, condition (ii) is satisfied.

**Conditions (iii–vi).** This follows from Lemma A.2.

**Condition (vii).** This is trivially satisfied because there is no nuisance parameter $\eta$ in the current setting. □

The following corollary can be used to construct confidence intervals based on the duality of hypothesis testing and confidence intervals.

**Corollary A.3.1.** *Let* $\boldsymbol{\gamma_0}$ *equal the true vector of acceleration factors, Lemma A.3 then implies the following:*

$$n^{1/2} \cdot \boldsymbol{\Phi}(\boldsymbol{\gamma_0}; \hat{\boldsymbol{\alpha}}_n, \hat{\boldsymbol{\beta}}_n) \xrightarrow{d} \mathcal{N}(\mathbf{0}, \Lambda_0)$$

*where* $\Lambda_0 = \dot{\boldsymbol{\Phi}}_{\boldsymbol{\alpha},\boldsymbol{\beta}}(\boldsymbol{\gamma_0}; \boldsymbol{\alpha_0}, \boldsymbol{\beta_0}) \cdot \Sigma_0 \cdot \dot{\boldsymbol{\Phi}}_{\boldsymbol{\alpha},\boldsymbol{\beta}}(\boldsymbol{\gamma_0}; \boldsymbol{\alpha_0}, \boldsymbol{\beta_0})^\top$.

The univariate test statistic for the acceleration factor at $t_j$,

$$\gamma \mapsto z_j(\gamma) := n^{1/2} \frac{\Phi_j(\gamma; \hat{\boldsymbol{\alpha}}_n, \hat{\boldsymbol{\beta}}_n)}{\Lambda_{jj,0}} = n^{1/2} \frac{\hat{\beta}_{j,n} - f_0(\gamma \cdot t_j; \hat{\boldsymbol{\alpha}}_n)}{\Lambda_{jj,0}},$$

where $\Phi_j$ is the $j$'th element of $\boldsymbol{\Phi}$ and $\Lambda_{jj,0}$ the $j$'th diagonal element of $\Lambda_0$, is a function of the acceleration factor at $t_j$ only. This follows from the definition of the contrast function in (1). A $1 - \alpha$ confidence interval for the time-specific acceleration factor at $t_j$ is then be obtained as

$$\left\{ \gamma : z_j(\gamma)^2 \leq \chi^2_{1-\alpha,1} \right\},$$

[3] This function has a unique root with probability tending to 1 under the stated assumptions. If for a given $(\hat{\boldsymbol{\alpha}}_n, \hat{\boldsymbol{\beta}}_n)$, there is no root (or more than 1), we can set $\hat{\boldsymbol{\gamma}}_n = 1$.

where $\chi^2_{1-\alpha,1}$ is the $1-\alpha$ percentile of the $\chi^2_1$-distribution and $\Lambda_{jj,0}$ has been replaced with a consistent estimate.

Generally, $z_j(\gamma)$ depends on $\gamma$ in a complex manner that depends on the interpolation and extrapolation method. Analytical solutions for the confidence interval limits may be available for some interpolation and extrapolation methods, but not for others. We, therefore, find the confidence intervals numerically in the *TCT* R package. The limits of the confidence interval are computed by solving $z(\gamma)^2 = \chi^2_{1-\alpha,1}$ for $\gamma$. This equation is expected to have two or fewer solutions[4]: maximally one where $z(\gamma) > 0$ and maximally one where $z(\gamma) < 0$. Hence, we expect $z(\gamma) = \sqrt{\chi^2_{1-\alpha,1}}$ and $z(\gamma) = -\sqrt{\chi^2_{1-\alpha,1}}$ to each have maximally one solution. Finding these solutions is a standard numerical problem.

**Remark 1.** *As mentioned in the text, we replace $\Lambda_{jj,0}$ with a consistent estimate $\hat{\Lambda}_{jj,n}$ in practice. By Slutsky's theorem, the limiting distribution does not change because the estimator is consistent. This estimate, $\hat{\Lambda}_{jj,n}$, is a constant given the data, but depends on the estimate $\hat{\gamma}_{j,n}$. In principle, one could also let the estimate of $\Lambda_{jj,0}$ depend on the $\gamma$ in which $z_j(\gamma)$ is evaluated. The corresponding test statistic would then be*

$$n^{1/2}\frac{\hat{\beta}_{j,n} - f_0(\gamma \cdot t_j; \hat{\boldsymbol{\alpha}}_n)}{\hat{\Lambda}_{jj,n}(\gamma)}.$$

##### A.2.3 Weights-Based Estimator of Common Acceleration Factor

**Lemma A.4.** *Under Assumptions 1, 2, 6, 7, A.1, and assuming proportional slowing, the weights-based estimator $\hat{\gamma}_{\boldsymbol{w},n}$ for the common acceleration factor, defined as the root[5] of*

$$\gamma \in [0, 1+\delta/t_K] \mapsto \boldsymbol{w}^\top \cdot \boldsymbol{\Phi}(\gamma; \hat{\boldsymbol{\alpha}}_n, \hat{\boldsymbol{\beta}}_n)$$

*for a fixed vector of weights $\boldsymbol{w} \in \mathbb{R}^K$, is consistent and asymptotically normal:*

$$n^{1/2}(\hat{\gamma}_{\boldsymbol{w},n} - \gamma_0) \xrightarrow{d} \mathcal{N}(\boldsymbol{0}, \Omega_{\boldsymbol{w},0}),$$

*where*

$$\Omega_{\boldsymbol{w},0} = \left(\boldsymbol{w}^\top \cdot \dot{\boldsymbol{\Phi}}_{\boldsymbol{\gamma}}(\boldsymbol{\gamma_0}; \boldsymbol{\alpha_0}, \boldsymbol{\beta_0}) \cdot \boldsymbol{1}\right)^{-2} \cdot \boldsymbol{w}^\top \cdot \dot{\boldsymbol{\Phi}}_{\boldsymbol{\alpha},\boldsymbol{\beta}}(\boldsymbol{\gamma_0}; \boldsymbol{\alpha_0}, \boldsymbol{\beta_0}) \cdot \Sigma_0 \cdot \dot{\boldsymbol{\Phi}}_{\boldsymbol{\alpha},\boldsymbol{\beta}}(\boldsymbol{\gamma_0}; \boldsymbol{\alpha_0}, \boldsymbol{\beta_0})^\top \cdot \boldsymbol{w}.$$

*Proof.* We can apply Theorem A.1 where there is no nuisance parameter $\eta$. Hence, we only need to verify conditions (i–vii) from Theorem A.1 to complete the proof.

**Condition (i).** This follows directly from Assumption 7.

**Condition (ii).** Assumptions 1 and A.1 imply that $t \mapsto f_0(t; \boldsymbol{\alpha_0})$ is strictly monotone and continuous in $[0, t_K + \delta]$, which in turn implies that $\gamma \mapsto \beta_{j,0} - f_0(\gamma \cdot t_j; \boldsymbol{\alpha_0})$ is strictly monotone and continuous in $[0, 1 + \delta/t_K]$. Furthermore, these functions are either strictly increasing or strictly decreasing for all $j$. For $\boldsymbol{w}$ a vector of weights (and hence positive values), $\gamma \mapsto \sum_j w_j \cdot (\beta_{j,0} - f_0(\gamma \cdot t_j; \boldsymbol{\alpha_0}))$ is, therefore, also strictly monotone and continuous. Hence, if the latter function has a root, it is unique. By Assumption 2 and proportional slowing, this function has a

[4] $\gamma \mapsto \hat{\beta}_{j,n} - f_0(\gamma \cdot t_j; \hat{\boldsymbol{\alpha}}_n)$ is expected to be strictly monotone by Assumption 1; although this may fail in finite samples. If it is strictly monotone, then there can still be fewer than two solutions because the latter function is bounded. If it is not strictly monotone, more than two solutions are possible.

[5] This function has a unique root with probability tending to 1 under the stated assumptions. If for a given $(\hat{\boldsymbol{\alpha}}_n, \hat{\boldsymbol{\beta}}_n)$, there is no root (or more than 1), we can set $\hat{\gamma}_{\boldsymbol{w},n} = 1$.

root, which we denote by $\gamma_0$. (This is the common acceleration factor by Assumption 6.) Hence, $\sum_j w_j \cdot (\beta_{j,0} - f_0(\gamma_n \cdot t_j; \boldsymbol{\alpha_0})) \to 0$ implies that $\gamma_n \to \gamma_0$. Hence, condition (ii) is satisfied.

**Conditions (iii–vi).** This follows from Lemma A.2 and the fact that these conditions being satisfied for $\boldsymbol{\Phi}$ implies that the same conditions are satisfied for $\boldsymbol{w}^\top \cdot \boldsymbol{\Phi}$ where $\boldsymbol{w}$ is any vector of weights.

**Condition (vii).** This is trivially satisfied because there is no nuisance parameter $\eta$ in the current setting. □

The following corollary can be used to construct confidence intervals based on the duality of hypothesis testing and confidence intervals.

**Corollary A.4.1.** *Under proportional slowing and letting $\gamma_0$ be the true common acceleration factor, corollary A.3.1 then implies the following:*

$$n^{1/2} \cdot \boldsymbol{w}^\top \cdot \boldsymbol{\Phi}(\boldsymbol{\gamma_0}; \hat{\boldsymbol{\alpha}}_n, \hat{\boldsymbol{\beta}}_n) \xrightarrow{d} \mathcal{N}\left(\boldsymbol{0}, \boldsymbol{w}^\top \cdot \Lambda_0 \cdot \boldsymbol{w}\right)$$

*where $\Lambda_0$ is defined in corollary A.3.1.*

A confidence interval can be obtained in the same way as explained in Appendix A.2.2, but now using the following univariate test statistic for the common acceleration factor:

$$\gamma \mapsto z(\gamma) := n^{1/2} \frac{\boldsymbol{w}^\top \cdot \boldsymbol{\Phi}(\gamma; \hat{\boldsymbol{\alpha}}_n, \hat{\boldsymbol{\beta}}_n)}{\boldsymbol{w}^\top \cdot \Lambda_0 \cdot \boldsymbol{w}},$$

where, as before, $\Lambda_0$ is replaced with a consistent estimate in practice. Remark 1 also applies here if we change $\Lambda_{jj,0}$, $\hat{\Lambda}_{jj,n}$, and $\hat{\Lambda}_{jj,n}(\gamma)$ to, respectively, $\Lambda_0$, $\hat{\Lambda}_n$, and $\hat{\Lambda}_n(\gamma)$.

##### A.2.4 Adaptive Weights-Based Estimator

The following lemma shows that the asymptotic distribution of the weights-based estimator in the case where the weights are fixed, is the same as in the case where they were estimated. The only additional requirement is that the estimated weights vector converges to a fixed vector, but this convergence can be arbitrarily slow.

**Lemma A.5.** *Under the assumptions of Lemma A.4 and additionally assuming that $\hat{\boldsymbol{w}}_n \xrightarrow{P} \boldsymbol{w}_0$ for some weight vector $\boldsymbol{w}_0$, the adaptive weights-based estimator $\hat{\gamma}_{\hat{\boldsymbol{w}}_n,n}$ for the common acceleration factor, defined as the root[6] of*

$$\gamma \in [0, 1 + \delta/t_K] \mapsto \hat{\boldsymbol{w}}_n^\top \cdot \boldsymbol{\Phi}(\gamma; \hat{\boldsymbol{\alpha}}_n, \hat{\boldsymbol{\beta}}_n)$$

*is consistent and asymptotically normal:*

$$n^{1/2}(\hat{\gamma}_{\hat{\boldsymbol{w}}_n,n} - \gamma_0) \xrightarrow{d} \mathcal{N}(\boldsymbol{0}, \Omega_{\boldsymbol{w}_0}),$$

*where $\Omega_{\boldsymbol{w}_0}$ is defined in Lemma A.4.*

[6] This function has a unique root with probability tending to 1 under the stated assumptions. If for a given $(\hat{\boldsymbol{\alpha}}_n, \hat{\boldsymbol{\beta}}_n)$, there is no root (or more than 1), we can set $\hat{\gamma}_{\hat{\boldsymbol{w}}_n,n} = 1$.

*Proof.* We can apply Lemma A.1 where $\eta_n = \hat{\boldsymbol{w}}_n$. Hence, we only need to verify conditions (i–vii) from lemma A.1 to complete the proof.

**Condition (i).** This follows directly from Assumption 7.

**Condition (ii).** Part 1 follows from the proof of Lemma A.4. Part 2 follows from the proportional slowing assumption. Indeed, by proportional slowing we have that $\beta_{j,0} - f_0(\gamma_0 \cdot t_j; \boldsymbol{\alpha_0}) = 0$ for all $j$. Any weighted average (including with $\hat{\boldsymbol{w}}_n$) is a linear combination of zeroes and, hence, zero.

**Conditions (iii–vi).** This follows from Lemma A.2 and the fact that these conditions being satisfied for $\boldsymbol{\Phi}$ implies that the same conditions are satisfied for $\boldsymbol{w}^\top \cdot \boldsymbol{\Phi}$ where $\boldsymbol{w}$ is any vector of weights.

**Condition (vii).** This holds by assumption. □

**Remark 2.** *The fundamental reason why the sampling variability in $\hat{\boldsymbol{w}}_n$ can be ignored, is the same as for Z-estimators where the target and nuisance parameters are orthogonal (van der Vaart, 2000, p. 61). Under the proportional slowing assumption, any weight vector will yield a consistent estimator for the common acceleration factor. By contrast, if the proportional slowing assumption is violated, the limiting value of the estimator will depend on the weights and the orthogonality argument breaks down. Consequently, the asymptotic distribution may then depend on the estimator of the weights.*

Lemma A.5 only requires that the estimated weights, $\hat{\boldsymbol{w}}_n$, converge to some fixed weights vector. Under proportional slowing, the corresponding estimator for the common acceleration factor is consistent and asymptotically normal regardless of the limiting weights vector. Of course, the limiting value may affect efficiency. We, therefore, aim to estimate the following optimal weights:

$$\boldsymbol{w}_{\text{opt}} := \arg\min_{\boldsymbol{w}} \Omega_{\boldsymbol{w}}.$$

As made explicit in Lemma A.4, $\Omega_{\boldsymbol{w},0}$ depends $\gamma_0$, $(\boldsymbol{\alpha_0}, \boldsymbol{\beta_0})$, and $\Sigma_0$. Hence, we can write it as $\Omega(\boldsymbol{w}, \gamma_0, \boldsymbol{\alpha_0}, \boldsymbol{\beta_0}, \Sigma_0)$ and the corresponding estimate, for a given $\boldsymbol{w}$, as $\Omega(\boldsymbol{w}, \hat{\gamma}_{\boldsymbol{w},n}, \hat{\boldsymbol{\alpha}}_n, \hat{\boldsymbol{\beta}}_n, \hat{\Sigma}_n)$. The estimated optimal weights are defined as follows:

$$\hat{\boldsymbol{w}}_n := \arg\min_{\boldsymbol{w}} \Omega(\boldsymbol{w}, \hat{\gamma}_{\boldsymbol{w},n}, \hat{\boldsymbol{\alpha}}_n, \hat{\boldsymbol{\beta}}_n, \hat{\Sigma}_n) \quad \text{subject to} \quad 0 \leq w_j \leq 1 \text{ for } j = 1, \dots, K \quad \text{and} \quad \sum_{j=1}^{K} w_j = 1.$$

The above defined estimator for the optimal weights is implemented in the *TCT* R package using the following algorithm:

**Algorithm 1:** Computation of Estimated Optimal Weights $\hat{\boldsymbol{w}}_n$

**Input:** Initial weight vector $\tilde{\boldsymbol{w}}_0$ (e.g., a vector of ones); tolerance thresholds $\epsilon_1$, $\epsilon_2$
**Output:** Estimated optimal weights $\hat{\boldsymbol{w}}_n$

Compute $\hat{\gamma}_{\tilde{\boldsymbol{w}}_0,n}$ using the initial weights $\tilde{\boldsymbol{w}}_0$;
$\hat{\sigma}_0^2 \leftarrow \Omega(\tilde{\boldsymbol{w}}_0, \hat{\gamma}_{\tilde{\boldsymbol{w}}_0,n}, \hat{\boldsymbol{\alpha}}_n, \hat{\boldsymbol{\beta}}_n, \hat{\Sigma}_n)$;
Set $k \leftarrow 0$;
**repeat**
  $k \leftarrow k + 1$;
  $\tilde{\boldsymbol{w}}_k \leftarrow \arg\min_{\boldsymbol{w}} \Omega(\boldsymbol{w}, \hat{\gamma}_{\tilde{\boldsymbol{w}}_{k-1},n}, \hat{\boldsymbol{\alpha}}_n, \hat{\boldsymbol{\beta}}_n, \hat{\Sigma}_n)$ s.t. $0 \leq w_j \leq 1$ and $\sum_{j=1}^{K} w_j = 1$;
  Compute $\hat{\gamma}_{\tilde{\boldsymbol{w}}_k,n}$;
  $\hat{\sigma}_k^2 \leftarrow \Omega(\tilde{\boldsymbol{w}}_k, \hat{\gamma}_{\tilde{\boldsymbol{w}}_k,n}, \hat{\boldsymbol{\alpha}}_n, \hat{\boldsymbol{\beta}}_n, \hat{\Sigma}_n)$;
**until** $\|\tilde{\boldsymbol{w}}_k - \tilde{\boldsymbol{w}}_{k-1}\|_2^2 < \epsilon_1$ ***or*** $(\hat{\sigma}_k^2 - \hat{\sigma}_{k-1}^2)^2 < \epsilon_2$;
**return** $\tilde{\boldsymbol{w}}_k$

**Remark 3.** *A simpler estimator for the optimal weights would use* $(\hat{\boldsymbol{\alpha}}_n, \hat{\boldsymbol{\beta}}_n)$ *and* $\hat{\Sigma}_n$*, and some initial estimate for* $\gamma_0$*; for instance,* $\hat{\gamma}_{\boldsymbol{w},n}$ *where* $\boldsymbol{w}$ *is a vector of ones. As long as the initial estimate for* $\gamma_0$ *is consistent, this will not change the asymptotic distribution of* $\hat{\gamma}_{\hat{\boldsymbol{w}}_n,n}$ *although finite-sample behavior will be affected.*

##### A.3 Inference Based on Generalized Least Squares

We first define the generalized least-squares (GLS) criterion, discuss some of its properties, derive its partial derivatives, and introduce an additional assumption regarding inter- and extrapolation. We then study the asymptotics of the GLS estimator of the common acceleration factor. In the last subsection, we discuss how the minimized GLS criterion can be used for testing.

###### A.3.1 Preliminaries

We first introduce some additional notations, consistent with the notation in Theorem A.1. We let $\boldsymbol{\theta} := (\boldsymbol{\alpha}^\top, \gamma)^\top$, $\boldsymbol{g}(\boldsymbol{\theta}) := \left(\boldsymbol{\alpha}^\top, \boldsymbol{f_0}(\gamma \cdot \boldsymbol{t}; \boldsymbol{\alpha})^\top\right)^\top$, $\boldsymbol{f_0}(\gamma \cdot \boldsymbol{t}; \boldsymbol{\alpha}) := (f_0(\gamma \cdot t_1; \boldsymbol{\alpha}), \dots, f_0(\gamma \cdot t_K; \boldsymbol{\alpha}))^\top$, and $\boldsymbol{y}_n := (\hat{\boldsymbol{\alpha}}_n^\top, \hat{\boldsymbol{\beta}}_n^\top)^\top$. We rewrite the GLS estimator (as presented in the main text), now denoted by $\hat{\boldsymbol{\theta}}_n$ or $(\tilde{\boldsymbol{\alpha}}_n^\top, \tilde{\gamma}_n)^\top$, as follows:

$$\hat{\boldsymbol{\theta}}_n := \arg\min_{\boldsymbol{\theta} \in \Theta} Q\left(\boldsymbol{\theta}; \boldsymbol{y}_n, \hat{\Sigma}_n\right) \quad \text{where} \quad Q\left(\boldsymbol{\theta}; \boldsymbol{y}_n, \hat{\Sigma}_n\right) := (\boldsymbol{y}_n - g(\boldsymbol{\theta}))^\top \hat{\Sigma}_n^{-1} (\boldsymbol{y}_n - g(\boldsymbol{\theta})). \tag{2}$$

It follows that $\boldsymbol{\theta_0} = (\boldsymbol{\alpha_0}, \gamma_0)$ is the unique global minimizer of $\boldsymbol{\theta} \mapsto Q(\boldsymbol{\theta}; \boldsymbol{y_0}, \Sigma_0)$, where $Q(\boldsymbol{\theta_0}; \boldsymbol{y_0}, \Sigma_0) = 0$, if (i) $t \mapsto f(t; \boldsymbol{\alpha_0})$ is strictly monotone and (ii) the model is correctly specified. Indeed, we have that

$$\boldsymbol{g}(\boldsymbol{\theta_0}) = (\boldsymbol{\alpha_0}, \boldsymbol{f_0}(\gamma_0 \cdot \boldsymbol{t}; \boldsymbol{\alpha_0})) = (\boldsymbol{\alpha_0}, \boldsymbol{\beta_0}) = \boldsymbol{y_0},$$

for a correctly specified model (i.e., proportional slowing and correct interpolation as stated in Assumption 6). We further have that $Q(\boldsymbol{\theta}) > 0$ for $\boldsymbol{\theta} \neq \boldsymbol{\theta_0}$, which directly follows from $Q(\boldsymbol{\theta})$ being a quadratic form of the positive definite matrix $\Sigma^{-1}$ and the fact that $\boldsymbol{g}(\boldsymbol{\theta}) \neq \boldsymbol{g}(\boldsymbol{\theta_0})$ for $\boldsymbol{\theta} \neq \boldsymbol{\theta_0}$ if $t \mapsto f(t; \boldsymbol{\alpha_0})$ is strictly monotone.

By Assumption A.1 and for $\Sigma$ invertible, the function $\boldsymbol{\theta} \mapsto Q(\boldsymbol{\theta}; \boldsymbol{y}, \Sigma)$ is differentiable in $\mathbb{R}^{K+1} \times [0, 1 + \delta/t_K]$ with the following vector of partial derivatives:

$$\boldsymbol{\Phi}(\boldsymbol{\theta}; \boldsymbol{y}, \Sigma) := -2 \cdot J_{\boldsymbol{g}}(\boldsymbol{\theta})^\top \Sigma^{-1} (\boldsymbol{y} - \boldsymbol{g}(\boldsymbol{\theta})), \tag{3}$$

where

$$J_{\boldsymbol{g}}(\boldsymbol{\theta}) = \begin{pmatrix} I_{K+1} & \boldsymbol{0_{K+1}} \\ \frac{\partial f_0(\gamma \cdot t_1; \boldsymbol{\alpha})}{\partial \boldsymbol{\alpha}'} & \frac{\partial f_0(\gamma \cdot t_1; \boldsymbol{\alpha})}{\partial \gamma} \\ \vdots & \vdots \\ \frac{\partial f_0(\gamma \cdot t_K; \boldsymbol{\alpha})}{\partial \boldsymbol{\alpha}'} & \frac{\partial f_0(\gamma \cdot t_K; \boldsymbol{\alpha})}{\partial \gamma} \end{pmatrix}. \tag{4}$$

Under additionally Assumptions 1 and 2, we have that $J_{\boldsymbol{g}}(\boldsymbol{\theta_0})$ is of full rank because $\left.\frac{\partial f_0(\gamma \cdot t_j; \boldsymbol{\alpha_0})}{\partial \gamma}\right|_{\gamma = \gamma_0} \neq 0$ under these assumptions.

For deriving the asymptotic distribution of the GLS estimator in (7), we will need to make one additional regularity assumption about the inter- and extrapolation method.

**Assumption A.2.** *Use the same notation as in Assumption A.1. We assume that* $t \mapsto f(t; \boldsymbol{y})$ *is twice continuously differentiable for* $t \in [0, t_K + \delta]$ *and* $(t, \boldsymbol{y}) \mapsto \frac{\partial f(t; \boldsymbol{y})}{\partial \boldsymbol{y}}$ *is continuous on* $[0, t_K + \delta] \times \mathbb{R}^{K+1}$ *for some* $\delta > 0$ *(not depending on* $\boldsymbol{y}$*).*

Under assumption A.2, the Hessian $\nabla^2_{\boldsymbol{\theta}} Q(\boldsymbol{\theta}; \boldsymbol{y_0}, \Sigma_0)$ exists and can be expressed as follows:

$$\nabla^2_{\boldsymbol{\theta}} Q(\boldsymbol{\theta}; \boldsymbol{y_0}, \Sigma_0) = 2 \cdot J_{\boldsymbol{g}}(\boldsymbol{\theta})^\top \Sigma_0^{-1} J(\boldsymbol{\theta}) - 2 \cdot \sum_{i}^{2K+1} \sum_{j}^{2K+1} (y_{i,0} - g_i(\boldsymbol{\theta}))\, \Sigma^{-1}_{ij,0} \nabla^2_{\boldsymbol{\theta}} g_j(\boldsymbol{\theta}), \tag{5}$$

where $\Sigma^{-1}_{ij,0}$ is the $(i,j)$ element of $\Sigma_0^{-1}$. For $\boldsymbol{\theta} = \boldsymbol{\theta_0}$ and a correctly specified model, this expression simplifies as follows:

$$\nabla^2_{\boldsymbol{\theta}} Q(\boldsymbol{\theta_0}; \boldsymbol{y_0}, \Sigma_0) = 2 \cdot J_{\boldsymbol{g}}(\boldsymbol{\theta_0})^\top \Sigma_0^{-1} J_{\boldsymbol{g}}(\boldsymbol{\theta_0}).$$

As expected, the hessian evaluated in the true parameter, $\nabla^2_{\boldsymbol{\theta}} Q(\boldsymbol{\theta_0}; \boldsymbol{y_0}, \Sigma_0)$, is positive definite (if $J_{\boldsymbol{g}}(\boldsymbol{\theta_0})$ is full rank, which is $K+2$). However, the hessian may not be positive definite for all $\boldsymbol{\theta}$.

**Remark 4.** *The estimating function $\boldsymbol{\theta} \mapsto \boldsymbol{\Phi}(\boldsymbol{\theta}; \boldsymbol{y_0}, \Sigma_0)$, defined above, may not uniquely define the target parameter because $\boldsymbol{\theta} \mapsto \boldsymbol{\Phi}(\boldsymbol{\theta}; \boldsymbol{y_0}, \Sigma_0)$ may have more than one root. Indeed, the target parameter $\boldsymbol{\theta_0} := (\boldsymbol{\alpha_0}, \gamma_0)$ is a root because $\boldsymbol{y_0} - \boldsymbol{g}(\boldsymbol{\theta_0}) = \boldsymbol{0}$ under a correctly specified model. However, there may exist a $\boldsymbol{\theta^*} \neq \boldsymbol{\theta_0}$ such that $\boldsymbol{\Phi}(\boldsymbol{\theta^*}; \boldsymbol{y_0}, \Sigma_0) = \boldsymbol{0}$ (e.g., in a local optimum). Consequently, condition (ii) of Theorem A.1 may not hold and additional work is required to prove the consistency and asymptotic normality of the GLS estimator (7). This is done in the next subsection.*

##### A.3.2 Asymptotic Distribution

We first state some known technical results that will be used for proving consistency and asymptotic normality of the GLS estimator (7).

**Lemma A.6** (Problem 5.27 (van der Vaart, 2000))**.** *Let $\boldsymbol{\theta} \in \Theta \mapsto Q(\boldsymbol{\theta})$ be continuous where $\Theta \subset \mathbb{R}^p$ is compact and let $\boldsymbol{\theta_0}$ be the unique minimizer that is an interior point of $\Theta$. Then for every open $G \ni \boldsymbol{\theta_0}$, it holds that*

$$Q(\boldsymbol{\theta_0}) < \inf_{\boldsymbol{\theta} \notin G} Q(\boldsymbol{\theta}). \tag{6}$$

*Proof.* Suppose that the conditions hold, but that (6) does not hold. Then there exists a $G$ such that

$$Q(\boldsymbol{\theta_0}) \geq \inf_{\boldsymbol{\theta} \notin G} Q(\boldsymbol{\theta}).$$

The subset $G^c \subset \Theta$ is compact because a closed subset of a compact set is compact. The minimum of a continuous function over a compact set exists; hence, there exists a $\boldsymbol{\theta^*} \in G^c$ such that $Q(\boldsymbol{\theta^*}) = \inf_{\boldsymbol{\theta} \notin G} Q(\boldsymbol{\theta}) \leq Q(\boldsymbol{\theta_0})$. Hence, $\boldsymbol{\theta_0}$ is not the unique minimizer, which is a contradiction. Hence, (6) must hold. □

**Corollary A.6.1.** *Let $Q(\boldsymbol{\theta}; \boldsymbol{y}, \Sigma)$ be as defined in (2) and let $\Theta$ be a compact set such that*

*(i) $\Theta \subseteq \mathbb{R}^{K+1} \times [0, 1 + t_K/\delta]$ for $\delta > 0$ defined in Assumption A.1*

*(ii) $\boldsymbol{\theta_0}$ is an interior point of $\Theta$.*

*Under proportional slowing and Assumptions 1, 6, 7, and A.1, it follows that for every open $G \ni \boldsymbol{\theta_0}$*

$$Q(\boldsymbol{\theta_0}; \boldsymbol{y_0}, \Sigma_0) < \inf_{\boldsymbol{\theta} \notin G} Q(\boldsymbol{\theta}; \boldsymbol{y_0}, \Sigma_0).$$

*Proof.* From proportional slowing and Assumptions 1 and 6, follows that $\boldsymbol{\theta_0}$ is the unique minimizer (as explained in Appendix A.3.1 where $\Sigma_0$ is invertible by Assumption 7). We can now apply Lemma A.6 if $\boldsymbol{\theta} \mapsto Q(\boldsymbol{\theta}; \boldsymbol{y_0}, \Sigma)$ is continuous. The latter follows if $(\gamma, \boldsymbol{\alpha}) \mapsto f_0(\gamma \cdot t_j; \boldsymbol{\alpha})$ is continuous in $\Theta$ for all $j$, which follows from Assumption A.1 as follows: Let $(\gamma_n, \boldsymbol{\alpha}_n) \to (\gamma, \boldsymbol{\alpha}) \in \Theta$, then it follows that

$$\begin{aligned} |f_0(\gamma_n \cdot t_j; \boldsymbol{\alpha}_n) - f_0(\gamma \cdot t_j; \boldsymbol{\alpha})| &\leq |f_0(\gamma_n \cdot t_j; \boldsymbol{\alpha}_n) - f_0(\gamma_n \cdot t_j; \boldsymbol{\alpha})| + |f_0(\gamma_n \cdot t_j; \boldsymbol{\alpha}) - f_0(\gamma \cdot t_j; \boldsymbol{\alpha})| \\ &\leq \sup_{t \in [0, t_K + \delta]} |f_0(t; \boldsymbol{\alpha}_n) - f_0(t; \boldsymbol{\alpha})| + |f_0(\gamma_n \cdot t_j; \boldsymbol{\alpha}) - f_0(\gamma \cdot t_j; \boldsymbol{\alpha})| \,. \end{aligned}$$

Condition (ii) of Assumption A.1 implies that $\sup_{t \in [0, t_K + \delta]} |f_0(t; \boldsymbol{\alpha}_n) - f_0(t; \boldsymbol{\alpha})| = o(1)$. By assumption, we have that $\gamma \in [0, 1 + t_K/\delta]$ and, consequently, $\gamma \cdot t_j \in [0, t_K + \delta]$. This implies that $|f_0(\gamma_n \cdot t_j; \boldsymbol{\alpha}) - f_0(\gamma \cdot t_j; \boldsymbol{\alpha})| = o(1)$ because $t \mapsto f_0(t; \boldsymbol{\alpha})$ is continuous in $[0, t_K + \delta]$ by Assumption A.1. Hence, the expression in the above display is $o(1)$. □

**Lemma A.7** (Uniform convergence). *Let $\boldsymbol{\theta} \in \Theta \mapsto Q(\boldsymbol{\theta}; \boldsymbol{y}, \Sigma)$ be as defined in (2) and $\Theta$ compact. Under Assumptions 7 and A.1, it follows that*

$$\sup_{\boldsymbol{\theta} \in \Theta} \left| Q(\boldsymbol{\theta}; \boldsymbol{y}_n, \hat{\Sigma}_n) - Q(\boldsymbol{\theta}; \boldsymbol{y_0}, \Sigma_0) \right| \xrightarrow{P} 0.$$

*Proof.* Note that

$$Q(\boldsymbol{\theta}; \boldsymbol{y}_n, \hat{\Sigma}_n) = \sum_{i=1}^{2K+1} \sum_{j=1}^{2K+1} \hat{p}_{ij,n} \cdot (y_{i,n} - g_i(\boldsymbol{\theta})) \cdot (y_{j,n} - g_j(\boldsymbol{\theta}))$$

and

$$Q(\boldsymbol{\theta}; \boldsymbol{y_0}, \Sigma_0) = \sum_{i=1}^{2K+1} \sum_{j=1}^{2K+1} p_{ij,0} \cdot (y_{i,0} - g_i(\boldsymbol{\theta})) \cdot (y_{j,0} - g_j(\boldsymbol{\theta}))$$

where $\hat{p}_{ij,n}$ is the $(i,j)$ element of $\Sigma^{-1}$, and similarly for $p_{ij,0}$ and $\Sigma_0^{-1}$. It follows that

$$\begin{aligned}
\sup_{\boldsymbol{\theta}\in\Theta}|Q(\boldsymbol{\theta};\boldsymbol{y_n},\hat{\Sigma}_n)-Q(\boldsymbol{\theta};\boldsymbol{y_0},\Sigma_0)| &\le \sup_{\boldsymbol{\theta}\in\Theta}\left|\sum_{i=1}^{2K+1}\sum_{j=1}^{2K+1}p_{ij,0}\cdot A_{ij,n}(\boldsymbol{\theta})\right| \\
&\quad+\sup_{\boldsymbol{\theta}\in\Theta}\left|\sum_{i=1}^{2K+1}\sum_{j=1}^{2K+1}(\hat{p}_{ij,n}-p_{ij,0})\cdot(y_{i,n}-g_i(\boldsymbol{\theta}))\cdot(y_{j,n}-g_j(\boldsymbol{\theta}))\right| \\
&\quad(A_{ij,n}(\boldsymbol{\theta})\text{ defined below}) \\
&\le \sum_{i=1}^{2K+1}\sum_{j=1}^{2K+1}|p_{ij,0}|\cdot\sup_{\boldsymbol{\theta}\in\Theta}\left|A_{ij,n}(\boldsymbol{\theta})\right| \\
&\quad+\sum_{i=1}^{2K+1}\sum_{j=1}^{2K+1}|(\hat{p}_{ij,n}-p_{ij,0})|\cdot\sup_{\boldsymbol{\theta}\in\Theta}|(y_{i,n}-g_i(\boldsymbol{\theta}))\cdot(y_{j,n}-g_j(\boldsymbol{\theta}))| \\
&\le O(1)\sum_{i=1}^{2K+1}\sum_{j=1}^{2K+1}\sup_{\boldsymbol{\theta}\in\Theta}\left|A_{ij,n}(\boldsymbol{\theta})\right| \\
&\quad+o_P(1)\sum_{i=1}^{2K+1}\sum_{j=1}^{2K+1}\sup_{\boldsymbol{\theta}\in\Theta}|(y_{i,n}-g_i(\boldsymbol{\theta}))\cdot(y_{j,n}-g_j(\boldsymbol{\theta}))| \\
&\le O(1)\sum_{i=1}^{2K+1}\sum_{j=1}^{2K+1}\sup_{\boldsymbol{\theta}\in\Theta}\left|A_{ij,n}(\boldsymbol{\theta})\right|+o_P(1)O(1)\sup_{\boldsymbol{\theta}\in\Theta}\|\boldsymbol{y}_n-\boldsymbol{g}(\boldsymbol{\theta})\|_2^2 \\
&\le O(1)\sum_{i=1}^{2K+1}\sum_{j=1}^{2K+1}\sup_{\boldsymbol{\theta}\in\Theta}\left|A_{ij,n}(\boldsymbol{\theta})\right|+o_P(1)\sup_{\boldsymbol{\theta}\in\Theta}\left(\|\boldsymbol{y}_n\|_2^2+\|\boldsymbol{g}(\boldsymbol{\theta})\|_2^2\right) \\
&\le O(1)\sum_{i=1}^{2K+1}\sum_{j=1}^{2K+1}\sup_{\boldsymbol{\theta}\in\Theta}\left|A_{ij,n}(\boldsymbol{\theta})\right|+o_P(1)O_P(1) \\
&\quad(\|\boldsymbol{y}_n\|_2=O_P(1)\text{ and }\boldsymbol{g}(\boldsymbol{\theta})\text{ is bounded})
\end{aligned}$$

where

$$\begin{aligned}
A_{ij,n}(\boldsymbol{\theta}) &:= (y_{i,n}-g_i(\boldsymbol{\theta}))\cdot(y_{j,n}-g_j(\boldsymbol{\theta}))-(y_{i,0}-g_i(\boldsymbol{\theta}))\cdot(y_{j,0}-g_j(\boldsymbol{\theta})) \\
&= (y_{i,n}-g_i(\boldsymbol{\theta}))\cdot(y_{j,n}-g_j(\boldsymbol{\theta}))-\{(y_{i,0}-y_{i,n})+(y_{i,n}-g_i(\boldsymbol{\theta}))\}\cdot(y_{j,0}-g_j(\boldsymbol{\theta})) \\
&= (y_{i,n}-g_i(\boldsymbol{\theta}))\cdot\{(y_{j,n}-g_j(\boldsymbol{\theta}))-(y_{j,0}-g_j(\boldsymbol{\theta}))\}-(y_{i,0}-y_{i,n})\cdot(y_{j,0}-g_j(\boldsymbol{\theta})) \\
&= (y_{i,n}-g_i(\boldsymbol{\theta}))\cdot(y_{j,n}-y_{j,0})-(y_{i,0}-y_{i,n})\cdot(y_{j,0}-g_j(\boldsymbol{\theta}))
\end{aligned}$$

Further let

$$A^1_{ij,n}(\boldsymbol{\theta}) := (y_{i,n}-g_i(\boldsymbol{\theta}))\cdot(y_{j,n}-y_{j,0})$$

and

$$A^2_{ij,n}(\boldsymbol{\theta}) := (y_{i,0}-y_{i,n})\cdot(y_{j,0}-g_j(\boldsymbol{\theta}))\,.$$

Because $|A_{ij,n}(\boldsymbol{\theta})|=|A^1_{ij,n}(\boldsymbol{\theta})-A^2_{ij,n}(\boldsymbol{\theta})|\le|A^1_{ij,n}(\boldsymbol{\theta})|+|A^2_{ij,n}(\boldsymbol{\theta})|$ by the triangle inequality, we can complete the proof by showing that $\sup_{\boldsymbol{\theta}\in\Theta}\left|A^1_{ij,n}(\boldsymbol{\theta})\right|=o_P(1)$ and $\sup_{\boldsymbol{\theta}\in\Theta}\left|A^2_{ij,n}(\boldsymbol{\theta})\right|=o_P(1)$. We now show the former; the latter follows analogously.

$$\begin{aligned}\sup_{\boldsymbol{\theta}\in\Theta}\left|A^1_{ij,n}(\boldsymbol{\theta})\right| &\leq \|\boldsymbol{y}_n-\boldsymbol{y_0}\|_2\cdot\sup_{\boldsymbol{\theta}\in\Theta}\|\boldsymbol{y}_n-\boldsymbol{g}(\boldsymbol{\theta})\|_2\\ &\leq o_P(1)\cdot\sup_{\boldsymbol{\theta}\in\Theta}\{\|\boldsymbol{y}_n\|_2+\|\boldsymbol{g}(\boldsymbol{\theta})\|_2\}\\ &\leq o_P(1)\cdot\left\{O_P(1)+\sup_{\boldsymbol{\theta}\in\Theta}\|\boldsymbol{g}(\boldsymbol{\theta})\|_2\right\}\\ &= o_P(1)\cdot O_P(1)=o_P(1),\end{aligned}$$

where (i) $\|\boldsymbol{y}_n-\boldsymbol{y_0}\|_2=o_P(1)$ and $\|\boldsymbol{y}_n\|_2=O_P(1)$ by Assumption 7 and (ii) $\sup_{\boldsymbol{\theta}\in\Theta}\|\boldsymbol{g}(\boldsymbol{\theta})\|_2<\infty$ by the compactness of $\Theta$ and Assumption A.1. □

**Lemma A.8** (Consistency)**.** *Let $\hat{\boldsymbol{\theta}}_n$ be the estimator defined in (7) that minimizes over a compact set $\Theta$ such that*

*(i) $\Theta\subseteq\mathbb{R}^{K+1}\times[0,1+\delta/t_K]$ for $\delta>0$ defined in Assumption A.1*

*(ii) $\boldsymbol{\theta_0}$ is an interior point of $\Theta$.*

*Under proportional slowing and Assumptions 1, 6, 7, and A.1, this estimator is consistent: $\hat{\boldsymbol{\theta}}_n\xrightarrow{P}\boldsymbol{\theta_0}$.*

*Proof.* Theorem 5.7 in van der Vaart (2000) shows that $\hat{\boldsymbol{\theta}}_n\xrightarrow{P}\boldsymbol{\theta_0}$ if

$$\sup_{\boldsymbol{\theta}\in\Theta}|Q(\boldsymbol{\theta};\boldsymbol{y}_n,\hat{\Sigma}_n)-Q(\boldsymbol{\theta};\boldsymbol{y_0},\Sigma_0)|\xrightarrow{P}0\quad\text{and}\quad\inf_{\boldsymbol{\theta}:\|\boldsymbol{\theta}-\boldsymbol{\theta_0}\|_2\geq\epsilon}Q(\boldsymbol{\theta_0};\boldsymbol{y_0},\Sigma_0)<Q(\boldsymbol{\theta};\boldsymbol{y_0},\Sigma_0)\quad\text{for every}\quad\epsilon>0.$$

The former follows from Lemma A.7 if $\Theta$ is compact and Assumptions 7 and A.1 hold. The latter follows from Corollary A.6.1 if $\Theta$ is compact and satisfies conditions (i–ii) of this Lemma, Assumptions 1, 6, and A.1 hold, and proportional slowing. □

**Lemma A.9** (Asymptotic normality)**.** *Let $\hat{\boldsymbol{\theta}}_n$ be the estimator defined in (7) that minimizes over a compact set $\Theta$ such that*

*(i) $\Theta\subseteq\mathbb{R}^{K+1}\times[0,1+\delta/t_K]$ for $\delta>0$ defined in Assumptions A.1 and A.2*

*(ii) $\boldsymbol{\theta_0}$ is an interior point of $\Theta$.*

*Under proportional slowing and Assumptions 1, 2, 6, 7, A.1, and A.2, this estimator is consistent and asymptotically normal:*

$$n^{1/2}\left(\hat{\boldsymbol{\theta}}_n-\boldsymbol{\theta_0}\right)\xrightarrow{d}\mathcal{N}\left(\boldsymbol{0},\Omega_0\right)\quad\textit{where}\quad\Omega:=\left(J_{\boldsymbol{g}}(\boldsymbol{\theta_0})^\top\Sigma_0^{-1}J_{\boldsymbol{g}}(\boldsymbol{\theta_0})\right)^{-1}.$$

*Proof.* We can apply Theorem A.1 (where $\eta_n=\Sigma_n$) without the first part of condition (ii) and condition (vi) if we can show consistency separately. Consistency of $\hat{\boldsymbol{\theta}}_n$ follows from Lemma A.8; hence, we only need to verify the remaining conditions of Theorem A.1 where the estimating function is defined in (3). The partial derivative matrices in the current setting are as follows:

$$\dot{\boldsymbol{\Phi}}_{\boldsymbol{\theta}}(\boldsymbol{\theta};\boldsymbol{y},\Sigma)=\nabla^2_{\boldsymbol{\theta}}Q(\boldsymbol{\theta};\boldsymbol{y},\Sigma)\quad\text{and}\quad\dot{\boldsymbol{\Phi}}_{\boldsymbol{y}}(\boldsymbol{\theta};\boldsymbol{y},\Sigma)=-2\cdot J_{\boldsymbol{g}}(\boldsymbol{\theta})^\top\Sigma^{-1}.$$

The former exists by Assumptions A.1 and A.2. The latter exists by Assumption A.1 only. (We implicitly assumed here that $\Sigma$ is invertible.)

We now verify the conditions of Theorem A.1, except the first part of condition (ii) and condition (vi).

**Condition (i).** This follows directly from Assumption 7.

**Condition (ii), second part.** We have that $\mathbf{\Phi}(\boldsymbol{\theta_0}; \boldsymbol{y_0}, \Sigma) = \mathbf{0}$ for any invertible matrix $\Sigma$ because $\boldsymbol{y_0} = \boldsymbol{g}(\boldsymbol{\theta_0})$ for a correctly specified model (i.e., under proportional slowing and Assumption 6).

**Condition (iii).** The matrix-valued function $\boldsymbol{\theta} \mapsto \dot{\mathbf{\Phi}}_{\boldsymbol{\theta}}(\boldsymbol{\theta}; \boldsymbol{y}, \Sigma)$ consists of products and sums of the following types of terms, as can be seen from (4–5),

$$\frac{\partial f_0(\gamma \cdot t_j; \boldsymbol{\alpha})}{\partial \alpha_{j'}} \quad \text{and} \quad \frac{\partial f_0(\gamma \cdot t_j; \boldsymbol{\alpha})}{\partial \gamma} \quad \text{and} \quad g_j(\boldsymbol{\theta}) \quad \text{and} \quad \nabla^2_{\boldsymbol{\theta}} g_j(\boldsymbol{\theta}).$$

The first term is continuous in $\boldsymbol{\theta} \in \Theta$ by Assumption A.2. The second term is continuous in $\boldsymbol{\theta} \in \Theta$ if $(t, \boldsymbol{\alpha}) \mapsto \frac{\partial f_0(t;\boldsymbol{\alpha})}{\partial t}$ is continuous for $t \in [0, t_K + \delta]$ (because $\gamma$ is restricted to $[0, 1 + \delta/t_K]$). This is shown to hold under Assumption A.1 in the proof of Lemma A.2. The third term is continuous in $\boldsymbol{\theta} \in \Theta$ if $(t, \boldsymbol{\alpha}) \mapsto f_0(t; \boldsymbol{\alpha})$ is continuous for $t \in [0, t_K + \delta]$. This follows from the mean value theorem because $(t, \boldsymbol{\alpha}) \mapsto \frac{\partial f_0(t;\boldsymbol{\alpha})}{\partial t}$ and $(t, \boldsymbol{\alpha}) \mapsto \frac{\partial f_0(t;\boldsymbol{\alpha})}{\partial \boldsymbol{\alpha}}$ are continuous for $t \in [0, t_K + \delta]$ (by the results for the first two terms). The fourth term is continuous by Assumption A.2.

Since the above four terms are continuous in $\boldsymbol{\theta}$, we have that $\boldsymbol{\theta} \mapsto \dot{\mathbf{\Phi}}_{\boldsymbol{\theta}}(\boldsymbol{\theta}; \boldsymbol{y}, \Sigma)$ is continuous as well (for any $\boldsymbol{y}$ and invertible $\Sigma$).

**Condition (iv).** The matrix-valued function $\boldsymbol{\theta} \mapsto \dot{\mathbf{\Phi}}_{\boldsymbol{y}}(\boldsymbol{\theta}; \boldsymbol{y}, \Sigma)$ consists of sums of the following two types of elements:

$$\frac{\partial f_0(\gamma \cdot t_j; \boldsymbol{\alpha})}{\partial \alpha_{j'}} \quad \text{and} \quad \frac{\partial f_0(\gamma \cdot t_j; \boldsymbol{\alpha})}{\partial \gamma}.$$

These were shown above to be continuous in $\boldsymbol{\theta}$.

**Condition (v).** Because $\dot{\mathbf{\Phi}}_{\boldsymbol{\theta}}(\boldsymbol{\theta}; \boldsymbol{y}, \Sigma)$ is the product of (i) the four terms mentioned above, which depend on $\boldsymbol{\theta}$ only and are continuous in $\boldsymbol{\theta}$, (ii) elements of $\boldsymbol{y}$, and (iii) elements of $\Sigma^{-1}$, we have that $(\boldsymbol{\theta}, \boldsymbol{y}, \Sigma) \mapsto \dot{\mathbf{\Phi}}_{\boldsymbol{\theta}}(\boldsymbol{\theta}; \boldsymbol{y}, \Sigma)$ is continuous at $(\boldsymbol{\theta_0}, \boldsymbol{y_0}, \Sigma_0)$ if $\Sigma_0$ is positive definite. Furthermore, we showed in Appendix A.3.1 that $\dot{\mathbf{\Phi}}_{\boldsymbol{\theta}}(\boldsymbol{\theta_0}; \boldsymbol{y}, \Sigma_0)$ is positive definite and, hence, invertible. This proves part 1 of condition (v).

Part 2 of condition (v) follows trivially from condition (iv) together with the fact that $\Sigma_0$ is positive definite.

**Condition (vi).** This condition is only required for the consistency result in Theorem A.1.

**Condition (vii).** This follows from Assumption 7.

From Theorem A.1 now follows that $\hat{\boldsymbol{\theta}}_n$ is asymptotically normal:

$$n^{1/2} \left( \hat{\boldsymbol{\theta}}_n - \boldsymbol{\theta_0} \right) \xrightarrow{d} \mathcal{N}\left( \mathbf{0}, \Omega_0 \right),$$

where

$$\begin{aligned}
\Omega_0 &= \left( \nabla^2_{\boldsymbol{\theta}} Q(\boldsymbol{\theta_0}; \boldsymbol{y_0}, \Sigma_0) \right)^{-1} \dot{\mathbf{\Phi}}_{\boldsymbol{y}}(\boldsymbol{\theta}; \boldsymbol{y_0}) \Sigma_0 \dot{\mathbf{\Phi}}_{\boldsymbol{y}}(\boldsymbol{\theta}; \boldsymbol{y_0})^\top \left( \nabla^2_{\boldsymbol{\theta}} Q(\boldsymbol{\theta_0}; \boldsymbol{y_0}, \Sigma_0) \right)^{-1} \\
&= \left( 2 \cdot J_{\boldsymbol{g}}(\boldsymbol{\theta_0})^\top \Sigma^{-1} J_{\boldsymbol{g}}(\boldsymbol{\theta_0}) \right)^{-1} \cdot 2 \cdot J_{\boldsymbol{g}}(\boldsymbol{\theta_0})^\top \Sigma_0^{-1} \Sigma_0 \cdot 2 \cdot \Sigma_0^{-1} J_{\boldsymbol{g}}(\boldsymbol{\theta_0}) \left( 2 \cdot J_{\boldsymbol{g}}(\boldsymbol{\theta_0})^\top \Sigma_0^{-1} J(\boldsymbol{\theta_0}) \right)^{-1} \\
&= \left( J_{\boldsymbol{g}}(\boldsymbol{\theta_0})^\top \Sigma_0^{-1} J_{\boldsymbol{g}}(\boldsymbol{\theta_0}) \right)^{-1} J_{\boldsymbol{g}}(\boldsymbol{\theta_0})^\top \Sigma_0^{-1} J_{\boldsymbol{g}}(\boldsymbol{\theta_0}) \left( J_{\boldsymbol{g}}(\boldsymbol{\theta_0})^\top \Sigma_0^{-1} J_{\boldsymbol{g}}(\boldsymbol{\theta_0}) \right)^{-1} \\
&= \left( J_{\boldsymbol{g}}(\boldsymbol{\theta_0})^\top \Sigma_0^{-1} J_{\boldsymbol{g}}(\boldsymbol{\theta_0}) \right)^{-1}.
\end{aligned}$$

□

##### A.3.3 Generalized Least Squares Criterion for Testing

The estimator in (7) and the distributional result in Lemma A.9 can be used to construct Wald-type hypothesis tests and confidence intervals. In this section, we show how the minimized GLS criterion can also be used for inferences. As will become clear, this resembles tests based on the residual sum of squares in linear models or based on the likelihood ratio in more general parametric models.

We first present some general results and then apply them to the GLS setting described before.

**General Theory** In a general linear model $Y = X\boldsymbol{\beta} + \boldsymbol{\epsilon}$ with fixed design matrix $X$ and normal residuals $\boldsymbol{\epsilon} \sim \mathcal{N}(\mathbf{0}, \Sigma)$ and $\Sigma$ known, exact inference is possible. In the current setting, however, we only have asymptotic normality of $\boldsymbol{y}_n$ (Assumption 7) and the model is not linear (i.e., we cannot write $\boldsymbol{g}(\boldsymbol{\theta})$ as $X\boldsymbol{\theta}$ for some fixed design matrix). Nonetheless, the exact results from general linear models may translate to useful asymptotic results under some conditions, as described in the following two lemmas.

**Lemma A.10.** *Consider the following general linear model with asymptotically normal residuals:*

$$\boldsymbol{y}_n = X\boldsymbol{\beta} + \boldsymbol{\epsilon}_n \quad \textit{where} \quad n^{1/2}\boldsymbol{\epsilon}_n \xrightarrow{d} \mathcal{N}(\mathbf{0}, \Sigma) \quad \textit{as} \quad n \to \infty,$$

*and*

- $\boldsymbol{y}_n \in \mathbb{R}^k$*: response vector,*
- $X \in \mathbb{R}^{k \times p}$*: full rank design matrix,*
- $\boldsymbol{\beta} \in \mathbb{R}^p$*: parameter vector,*
- $\Sigma \in \mathbb{R}^{k \times k}$*: known, positive-definite covariance matrix.*

*Let* $\hat{\boldsymbol{\beta}}_n := \left(X^\top \Sigma^{-1} X\right)^{-1} X^\top \Sigma^{-1} \boldsymbol{y}_n + o_P(n^{-1/2})$ *be an estimator of* $\boldsymbol{\beta_0}$*, the true coefficients vector, that is asymptotically equivalent to the generalized least-squares estimator. It then follows that*

$$n \cdot \left(\boldsymbol{y}_n - X\hat{\boldsymbol{\beta}}_n\right)^\top \Sigma^{-1} \left(\boldsymbol{y}_n - X\hat{\boldsymbol{\beta}}_n\right) \xrightarrow{d} \chi^2_{k-p} \quad \textit{as} \quad n \to \infty,$$

*where* $\chi^2_{k-p}$ *is a chi-squared distribution with* $k - p$ *degrees of freedom.*

*Proof.* We first transform the outcome variable and design matrix as follows:

$$\boldsymbol{z}_n := \Sigma^{-1/2}\boldsymbol{y}_n \quad \text{and} \quad W := \Sigma^{-1/2}X.$$

This yields the following linear model with (asymptotically) independent standard normal errors:

$$\boldsymbol{z}_n = W\boldsymbol{\beta} + \boldsymbol{e}_n \quad \text{where} \quad n^{1/2}\boldsymbol{e}_n \xrightarrow{d} \mathcal{N}(\mathbf{0}, I_k).$$

The corresponding projection matrix and residuals are as follows:

$$P_W := W\left(W^\top W\right)^{-1} W^\top \qquad \boldsymbol{r}_n := n^{1/2}(I - P_W)\,\boldsymbol{z}_n.$$

Because (i) $P_W$ is the projection matrix onto the column space of $W$ and (ii) $\boldsymbol{z}_n = W\boldsymbol{\beta_0} + \boldsymbol{e}_n$, we also have that $\boldsymbol{r}_n = n^{1/2}(I - P_W)\,\boldsymbol{e}_n$. Note that $P_W$ is symmetric and idempotent (Rencher & Schaalje, 2008, p. 228); consequently, $I - P_W$ is also symmetric and idempotent. Furthermore,

$I - P_W$ has rank $k-p$ because $P_W$ projects onto the column space of the rank $p$ matrix $W$. The corresponding residual sum of squares (RSS) are

$$\begin{aligned}\boldsymbol{r}_n^\top \boldsymbol{r}_n &= \left(n^{1/2}\boldsymbol{e}_n\right)^\top (I-P_W)^\top (I-P_W)\left(n^{1/2}\boldsymbol{e}_n\right)\\ &= \left(n^{1/2}\boldsymbol{e}_n\right)^\top (I-P_W)\left(n^{1/2}\boldsymbol{e}_n\right) \qquad (I-P_W \text{ is symmetric idempotent}),\end{aligned}$$

where $n^{1/2}\boldsymbol{e}_n \xrightarrow{d} \mathcal{N}(\boldsymbol{0}, I_k)$ by assumption. Since the above product is a continuous function of $n^{1/2}\boldsymbol{e}_n$, by the continuous mapping theorem, the distribution of $\boldsymbol{r}_n^\top \boldsymbol{r}_n$ converges to that of $Z^\top(I - P_W)Z$ where $Z \sim \mathcal{N}(\boldsymbol{0}, I_k)$. From the theory of quadratic forms (Rencher & Schaalje, 2008, Theorem 5.5) follows that $Z^\top(I-P_W)Z \sim \chi^2_{k-p}$. This implies that $\boldsymbol{r}_n^\top \boldsymbol{r}_n \xrightarrow{d} \chi^2_{k-p}$.

The proof is complete if we can show that

$$n \cdot \left(\boldsymbol{y}_n - X\hat{\boldsymbol{\beta}}_n\right)^\top \Sigma^{-1}\left(\boldsymbol{y}_n - X\hat{\boldsymbol{\beta}}_n\right) = \boldsymbol{r}_n^\top \boldsymbol{r}_n + o_P(1),$$

where the left-hand side is the RSS in the general linear model using an estimator $\hat{\boldsymbol{\beta}}_n$ that is asymptotically equivalent to the GLS estimator. We prove the above display in two steps: First, we show that $\boldsymbol{r}_n^\top \boldsymbol{r}_n$ equals the RSS in the general linear model using the GLS estimator, $\tilde{\boldsymbol{\beta}}_n$:

$$\begin{aligned}\boldsymbol{r}_n^\top \boldsymbol{r}_n &= n \cdot \{\boldsymbol{z}_n - P_W \boldsymbol{z}_n\}^\top \{\boldsymbol{z}_n - P_W\boldsymbol{z}_n\}\\ &= n \cdot \left\{\boldsymbol{z}_n - W\left(W^\top W\right)^{-1} W^\top \boldsymbol{z}_n\right\}^\top \left\{\boldsymbol{z}_n - W\left(W^\top W\right)^{-1} W^\top \boldsymbol{z}_n\right\}\\ &= n \cdot \left\{\Sigma^{-1/2}\boldsymbol{y}_n - \Sigma^{-1/2}X\left(X^\top\Sigma^{-1}W\right)^{-1} X^\top \Sigma^{-1}\boldsymbol{y}_n\right\}^\top \left\{\Sigma^{-1/2}\boldsymbol{y}_n - \Sigma^{-1/2}X\left(X^\top\Sigma^{-1}W\right)^{-1} X^\top \Sigma^{-1}\boldsymbol{y}_n\right\}\\ &= n \cdot \left(\Sigma^{-1/2}\boldsymbol{y}_n - \Sigma^{-1/2}X\tilde{\boldsymbol{\beta}}_n\right)^\top \left(\Sigma^{-1/2}\boldsymbol{y}_n - \Sigma^{-1/2}X\tilde{\boldsymbol{\beta}}_n\right)\\ &= n \cdot \left(\boldsymbol{y}_n - X\tilde{\boldsymbol{\beta}}_n\right)^\top \Sigma^{-1}\left(\boldsymbol{y}_n - X\tilde{\boldsymbol{\beta}}_n\right) =: \text{RSS}.\end{aligned}$$

Second, we show that the RSS based on $\tilde{\boldsymbol{\beta}}_n$ and $\hat{\boldsymbol{\beta}}_n$ are asymptotically equivalent:

$$\begin{aligned}n \cdot \left(\boldsymbol{y}_n - X\hat{\boldsymbol{\beta}}_n\right)^\top \Sigma^{-1}\left(\boldsymbol{y}_n - X\hat{\boldsymbol{\beta}}_n\right) &= n \cdot \left(\boldsymbol{y}_n - X\tilde{\boldsymbol{\beta}}_n + o_P(n^{-1/2})\right)^\top \Sigma^{-1}\left(\boldsymbol{y}_n - X\tilde{\boldsymbol{\beta}}_n + o_P(n^{-1/2})\right)\\ &= n \cdot \left(\boldsymbol{y}_n - X\tilde{\boldsymbol{\beta}}_n\right)^\top \Sigma^{-1}\left(\boldsymbol{y}_n - X\tilde{\boldsymbol{\beta}}_n\right)\\ &\quad + 2n \cdot o_P(n^{-1/2}) \cdot \Sigma^{-1}\left(\boldsymbol{y}_n - X\tilde{\boldsymbol{\beta}}_n\right)\\ &\quad + n \cdot o_P(n^{-1/2}) \cdot \Sigma^{-1} \cdot o_P(n^{-1/2})\\ &= \text{RSS} + o_P(1) \cdot n^{1/2} \cdot \left(\boldsymbol{y}_n - X\tilde{\boldsymbol{\beta}}_n\right) + o_P(1)\\ &= \text{RSS} + o_P(1)O_P(1) + o_P(1) = \text{RSS} + o_P(1).\end{aligned}$$

In the penultimate equality, we used the following result:

$$\begin{aligned}n^{1/2}\left(\boldsymbol{y}_n - X\tilde{\boldsymbol{\beta}}_n\right) &= n^{1/2}\left(\boldsymbol{y}_n - X\boldsymbol{\beta}_{\boldsymbol{0}}\right) + X \cdot n^{1/2}\left(\boldsymbol{\beta}_{\boldsymbol{0}} - \tilde{\boldsymbol{\beta}}_n\right)\\ &= n^{1/2} \cdot \boldsymbol{\epsilon}_n + X \cdot O_P(1) = O_P(1).\end{aligned}$$

□

**Lemma A.11.** *Consider the same model as in Lemma A.10 and let $X_1$ and $X_2$ be, respectively, $k \times p$ and $k \times l$ full-rank design matrices such that $\mathbf{C}(X_1) \subset \mathbf{C}(X_2)$ where $\mathbf{C}(\cdot)$ denotes the column space. Consider the following estimators:*

$$\hat{\boldsymbol{\beta}}_{1,n} := \left(X_1^\top \Sigma^{-1} X_1\right)^{-1} X_1^\top \Sigma^{-1} \boldsymbol{y}_n + o_P(n^{-1/2}) \quad \text{and} \quad \hat{\boldsymbol{\beta}}_{2,n} := \left(X_2^\top \Sigma^{-1} X_2\right)^{-1} X_2^\top \Sigma^{-1} \boldsymbol{y}_n + o_P(n^{-1/2}).$$

*Letting $\boldsymbol{\beta_{1,0}}$ and $\boldsymbol{\beta_{2,0}}$ be the true coefficient vectors. If $X_1\boldsymbol{\beta_{1,0}} = X_2\boldsymbol{\beta_{2,0}}$, it follows that*

$$n \cdot \left(\boldsymbol{y}_n - X_1\hat{\boldsymbol{\beta}}_{1,n}\right)^\top \Sigma^{-1} \left(\boldsymbol{y}_n - X_1\hat{\boldsymbol{\beta}}_{1,n}\right) - n \cdot \left(\boldsymbol{y}_n - X_2\hat{\boldsymbol{\beta}}_{2,n}\right)^\top \Sigma^{-1} \left(\boldsymbol{y}_n - X_2\hat{\boldsymbol{\beta}}_{2,n}\right) \xrightarrow{d} \chi^2_{l-p},$$

*where $\chi^2_{l-p}$ is a chi-squared distribution with $l-p$ degrees of freedom.*

*Proof.* As in the proof of Lemma A.10, we first transform the general linear models to linear models with asymptotically independent standard normal errors:

$$\boldsymbol{z}_n := \Sigma^{-1/2}\boldsymbol{y}_n \quad \text{and} \quad W_1 := \Sigma^{-1/2}X_1 \quad \text{and} \quad W_2 := \Sigma^{-1/2}X_2$$

$$\boldsymbol{z}_n = W_1\boldsymbol{\beta_1} + \boldsymbol{e}_n \quad \text{and} \quad \boldsymbol{z}_n = W_2\boldsymbol{\beta_2} + \boldsymbol{e}_n \quad \text{where} \quad n^{1/2}\boldsymbol{e}_n \xrightarrow{d} \mathcal{N}(\mathbf{0}, I_k).$$

From the proof of Lemma A.10 follows that the residual sums of squares (RSS) based on the GLS estimators in the above two models are asymptotically equivalent to the RSS in the general linear models using $\hat{\boldsymbol{\beta}}_{1,n}$ and $\hat{\boldsymbol{\beta}}_{2,n}$. Hence, the proof is complete if we can show that the difference of the RSSs based on the GLS estimators is chi-squared with $l-p$ degrees of freedom.

Define the following two projection matrices, corresponding to the two linear models defined in the last display:

$$P_{W_1} := W_1^\top \left(W_1^\top W_1\right)^{-1} W_1 \quad \text{and} \quad P_{W_2} := W_2^\top \left(W_2^\top W_2\right)^{-1} W_2,$$

and the corresponding residuals as follows:

$$\boldsymbol{r}_{1,n} := n^{1/2}\left(I - P_{W_1}\right)\boldsymbol{z}_n \quad \text{and} \quad \boldsymbol{r}_{2,n} := n^{1/2}\left(I - P_{W_2}\right)\boldsymbol{z}_n.$$

Following the same arguments as in the proof of Lemma A.10, we have that $\boldsymbol{r}_{1,n} = n^{1/2}\left(I - P_{W_1}\right)\boldsymbol{e}_n$ and $\boldsymbol{r}_{2,n} = n^{1/2}\left(I - P_{W_2}\right)\boldsymbol{e}_n$. Hence, the difference in RSS is as follows:

$$\begin{aligned}
\boldsymbol{r}_{1,n}^\top\boldsymbol{r}_{1,n} - \boldsymbol{r}_{2,n}^\top\boldsymbol{r}_{2,n} &= \left(n^{1/2}\boldsymbol{e}_n\right)^\top \left\{\left(I - P_{W_1}\right)^\top\left(I - P_{W_1}\right) - \left(I - P_{W_2}\right)^\top\left(I - P_{W_2}\right)\right\}\left(n^{1/2}\boldsymbol{e}_n\right) \\
&= \left(n^{1/2}\boldsymbol{e}_n\right)^\top \left\{\left(I - P_{W_1}\right) - \left(I - P_{W_2}\right)\right\}\left(n^{1/2}\boldsymbol{e}_n\right) \\
&= \left(n^{1/2}\boldsymbol{e}_n\right)^\top \left(P_{W_2} - P_{W_1}\right)\left(n^{1/2}\boldsymbol{e}_n\right).
\end{aligned}$$

The matrix $P_{W_2} - P_{W_1}$ is symmetric because $P_{W_1}$ and $P_{W_2}$ are symmetric, and is idempotent:

$$\begin{aligned}
\left(P_{W_2} - P_{W_1}\right)\left(P_{W_2} - P_{W_1}\right) &= P_{W_2}P_{W_2} - P_{W_2}P_{W_1} - P_{W_1}P_{W_2} + P_{W_1}P_{W_1} \\
&= P_{W_2} - P_{W_1} - P_{W_1} + P_{W_1} = P_{W_2} - P_{W_1},
\end{aligned}$$

where the second equality follows from $P_{W_1}$ and $P_{W_2}$ being projection matrices onto nested subspaces. Furthermore, $P_{W_2} - P_{W_1}$ is of rank $l$ because it is the projection matrix onto $\mathbf{C}(W_2)\backslash\mathbf{C}(W_1)$.

From Rencher and Schaalje (2008, Theorem 5.5) now follows that $Z^\top\left(P_{W_2} - P_{W_1}\right)Z \sim \chi^2_{l-p}$. Consequently, we can apply the continuous mapping theorem to obtain that

$$\boldsymbol{r}_{1,n}^\top\boldsymbol{r}_{1,n} - \boldsymbol{r}_{2,n}^\top\boldsymbol{r}_{2,n} \xrightarrow{d} \chi^2_{l-p}.$$

□

In practice, we do not know the true covariance matrix $\Sigma$, but we generally do have a consistent estimator $\hat{\Sigma}_n$ (e.g., as assumed in Assumption 7). Replacing $\Sigma$ with $\hat{\Sigma}_n$ does not change the limiting distribution, which follows from the continuous mapping theorem and the fact that $(X_n, \hat{\Sigma}_n) \xrightarrow{d} (X, \Sigma)$ if $X_n$ converges in distribution to $X$ and $\hat{\Sigma}_n$ converges in probability to a constant $\Sigma$.

**Application** Lemmas A.10 and A.11 can be applied to the GLS estimator described in Lemma A.9. Indeed, from the proof of the latter lemma (and Theorem A.1) follows that

$$n^{1/2}\left(\hat{\boldsymbol{\theta}}_n - \boldsymbol{\theta}_{\mathbf{0}}\right) = \left(J_{\boldsymbol{g}}(\boldsymbol{\theta}_{\mathbf{0}})^\top \Sigma_0^{-1} J_{\boldsymbol{g}}(\boldsymbol{\theta}_{\mathbf{0}})\right)^{-1} J_{\boldsymbol{g}}(\boldsymbol{\theta}_{\mathbf{0}})^\top \Sigma_0^{-1} \cdot n^{1/2}\left(\boldsymbol{y}_n - \boldsymbol{y}_{\mathbf{0}}\right) + o_P(1).$$

We can drop $n^{1/2}$ on both sides and reparameterize everything as follows:

$$\boldsymbol{\beta}(\boldsymbol{\theta}) := \boldsymbol{\theta} - \boldsymbol{\theta}_{\mathbf{0}} \quad \text{and} \quad \boldsymbol{\beta}_{\mathbf{0}} := \boldsymbol{\beta}(\boldsymbol{\theta}_{\mathbf{0}}) = \mathbf{0} \quad \text{and} \quad \hat{\boldsymbol{\beta}}_n := \boldsymbol{\beta}(\hat{\boldsymbol{\theta}}_n) = \hat{\boldsymbol{\theta}}_n - \boldsymbol{\theta}_{\mathbf{0}}.$$

Under this reparameterization, we have that

$$\hat{\boldsymbol{\beta}}_n = \left(J_{\boldsymbol{g}}(\boldsymbol{\theta}_{\mathbf{0}})^\top \Sigma^{-1} J_{\boldsymbol{g}}(\boldsymbol{\theta}_{\mathbf{0}})\right)^{-1} J_{\boldsymbol{g}}(\boldsymbol{\theta}_{\mathbf{0}})^\top \Sigma^{-1}\left(\boldsymbol{y}_n - \boldsymbol{y}_{\mathbf{0}}\right) + o_P(n^{-1/2})$$

is asymptotically equivalent to the GLS estimator in the following general linear model with asymptotically normal residuals:

$$\boldsymbol{y}_n - \boldsymbol{y}_{\mathbf{0}} = X\boldsymbol{\beta} + \boldsymbol{\epsilon}_n \quad \text{where} \quad n^{1/2}\boldsymbol{\epsilon}_n \xrightarrow{d} \mathcal{N}(\mathbf{0}, \Sigma) \quad \text{and} \quad X := J_{\boldsymbol{g}}(\boldsymbol{\theta}_{\mathbf{0}}).$$

Hence, we can apply Lemma A.10. This is useful for testing the proportional slowing assumption, as explained in the next remark.

**Remark 5** (Test for proportional slowing)**.** *From Lemma A.10 follows that*

$$n \cdot Q(\hat{\boldsymbol{\theta}}_n; \boldsymbol{y}_n, \hat{\Sigma}_n) = n \cdot \left(\boldsymbol{y}_n - \boldsymbol{g}(\hat{\boldsymbol{\theta}}_n)\right)^\top \hat{\Sigma}_n \left(\boldsymbol{y}_n - \boldsymbol{g}(\hat{\boldsymbol{\theta}}_n)\right) \xrightarrow{d} \chi^2_{K-1} \quad \text{as} \quad n \to \infty,$$

*where $\hat{\boldsymbol{\theta}}_n$ is the GLS estimator (7), if the conditions of Lemma A.9 hold. If either proportional slowing or Assumption 6 is violated, the limiting distribution will not be $\chi^2_{K-1}$. The statistic $n \cdot Q(\hat{\boldsymbol{\theta}}_n; \boldsymbol{y}_n, \hat{\Sigma}_n)$ can thus be used for testing these assumptions.*

From the following argument also follows that we can apply Lemma A.11. Let $\{\boldsymbol{g}(\boldsymbol{\theta}_1, \boldsymbol{c}) : \boldsymbol{\theta}_1 \in \Theta_1 \subset \mathbb{R}^p\}$ be a $p$-dimensional model for $\boldsymbol{y}_{\mathbf{0}}$ that contains the truth (i.e., $\boldsymbol{y}_{\mathbf{0}} = \boldsymbol{g}(\boldsymbol{\theta}_1)$ for some $\boldsymbol{\theta}_1 \in \Theta_1$). Further let $\left\{g(\boldsymbol{\theta}_1, \boldsymbol{\theta}_2) : (\boldsymbol{\theta}_1, \boldsymbol{\theta}_2) \in \Theta_1 \times \Theta_2 \subset \mathbb{R}^l\right\}$ be an $l$-dimensional model that contains the former (where $p < l$). Assume that $\boldsymbol{g}$ is differentiable. The Jacobian for the larger model is

$$J_{\boldsymbol{g}}(\boldsymbol{\theta}_1, \boldsymbol{\theta}_2) = \begin{pmatrix} \nabla^\top_{\boldsymbol{\theta}_1} g_1(\boldsymbol{\theta}_1, \boldsymbol{\theta}_2) & \nabla^\top_{\boldsymbol{\theta}_2} g_1(\boldsymbol{\theta}_1, \boldsymbol{\theta}_2) \\ \vdots & \vdots \\ \nabla^\top_{\boldsymbol{\theta}_1} g_k(\boldsymbol{\theta}_1, \boldsymbol{\theta}_2) & \nabla^\top_{\boldsymbol{\theta}_2} g_k(\boldsymbol{\theta}_1, \boldsymbol{\theta}_2) \end{pmatrix}.$$

We let $X_2 := J_{\boldsymbol{g}}(\boldsymbol{\theta}_1, \boldsymbol{\theta}_2)$ for $(\boldsymbol{\theta}_1, \boldsymbol{\theta}_2) = \boldsymbol{\theta}_{\mathbf{0}}$ be the design matrix corresponding to the larger model. From the above display follows that the first $p$ columns of $X_2$ correspond to the Jacobian for the smaller model evaluated in the true parameter, which we denote by $X_1$. Hence, the column space of $X_1$ is nested in the column space of $X_2$. So, if $X_2$ is of full rank, we can apply Lemma A.11.

**Remark 6** (Test of $H_0 : \gamma = \gamma_0$)**.** *Consider the model described in Appendix A.3.1 parameterized by $\boldsymbol{\theta} = (\boldsymbol{\alpha}, \gamma)$ for which the Jacobian is given in (4). Under the conditions of Lemma A.9, we can test $H_0 : \gamma = \gamma_0$ by comparing the minimized GLS criterion when minimizing over $\boldsymbol{\theta} = (\boldsymbol{\alpha}, \gamma)$ with the minimized criterion when $\gamma$ is fixed at $\gamma_0$.*

### B Efficiency

In this appendix, we introduce a particular notion of efficiency and corresponding theoretical results for the setting where $\boldsymbol{y_0} \in \mathbb{R}^k$ is a true vector and one is interested in estimating a particular function $\psi : \mathcal{M} \subseteq \mathbb{R}^k \to \mathbb{R}$ evaluated in $\boldsymbol{y_0}$. One only observes $\boldsymbol{y}_n \in \mathbb{R}^k$, which is a sample from $P_{0,n}$ such that

$$n^{1/2} (\boldsymbol{y}_n - \boldsymbol{y_0}) \underset{P_{0,n}}{\overset{d}{\to}} \mathcal{N}(\boldsymbol{0}, \Sigma_0), \tag{7}$$

where $\Sigma_0$ is assumed to be known. (In the main text and the other appendices, $\boldsymbol{y}_n$ is an estimator for $\boldsymbol{y_0}$.) The definitions and theoretical results presented next closely resemble the (semi-parametric) efficiency theory as presented in van der Vaart (2000). Our results are, however, not a special case of that theory as the above setup differs from that considered by van der Vaart (2000) (i.e., $n$ i.i.d. observations where $n \to \infty$). Our results are also not a generalization of the results in van der Vaart (2000), but our results and corresponding proofs are simpler because (7) is, arguably, a simple distributional setup.

#### B.1 Definitions and Setup

The following definitions have a straightforward counterpart in van der Vaart (2000), but as mentioned above, they are still different. The most fundamental difference is that van der Vaart (2000) considers i.i.d. observations sampled from some unknown distribution $P$ (which is an element of a statistical model $\mathcal{M}$, which is a collection of distributions). van der Vaart (2000) considers the problem of estimating a particular functional of $P$ evaluated in the true distribution $P_0$. In what follows, we replace the distribution $P$ with a vector $\boldsymbol{y} \in \mathbb{R}^k$ and consider the problem of estimating a particular function of $\boldsymbol{y}$ evaluated in the true vector $\boldsymbol{y_0}$.

##### B.1.1 Models

We next define the model and related notions. The model reflects the prior knowledge on the possible values for the true vector. To arrive at meaningful efficiency results, we will restrict ourselves to certain types of smooth models.

**Definition B.1** (Model)**.** *A model $\mathcal{M}$ is a collection of possible true vectors. In what follows, we will consider the following type of models: $\mathcal{M} = \{\boldsymbol{g}(\boldsymbol{\theta}) : \boldsymbol{\theta} \in \Theta \subseteq \mathbb{R}^l\}$ where $\boldsymbol{g} : \mathbb{R}^l \to \mathbb{R}^k$. The model is correctly specified if $\boldsymbol{y_0} = \boldsymbol{g}(\boldsymbol{\theta_0})$ for some $\boldsymbol{\theta_0} \in \Theta$.*

To derive meaningful efficiency results, one needs to put certain restrictions on the model. The concept of a "smooth model", as defined next, corresponds in spirit to differentiability in quadratic mean[7] as defined by van der Vaart (2000).

**Definition B.2** (Smooth model)**.** *Let $\boldsymbol{\theta_0}$ be an interior point of $\Theta$. The model $\mathcal{M} = \{\boldsymbol{g}(\boldsymbol{\theta}) : \boldsymbol{\theta} \in \Theta \subseteq \mathbb{R}^l\}$ is smooth at $\boldsymbol{\theta_0}$ if*

$$\boldsymbol{g}(\boldsymbol{\theta_0} + th) - \boldsymbol{g}(\boldsymbol{\theta_0}) = t \cdot a_h + o(t)$$

*for any[8] $h \in \mathbb{R}^l$ and $t \to 0$. The vector $a_h$ is termed a score of the model $\mathcal{M}$ at $\boldsymbol{\theta_0}$.*

[7] We do not re-use this term (and instead use "smooth" model) because our definition essentially requires partial differentiability; using "quadratic mean" would be confusing.

[8] For a given $h$ and $t$, it is possible that $\boldsymbol{\theta_0} + th \notin \Theta$. We can define $\boldsymbol{g}$ evaluated in that point as returning $\boldsymbol{0}$ because $\boldsymbol{\theta_0} + th \in \Theta$ will hold eventually as $t \to 0$ because $\boldsymbol{\theta_0}$ is an interior point of $\Theta$ by assumption.

**Remark 7.** *The function $\boldsymbol{g}$ being partially differentiable at $\boldsymbol{\theta_0}$ is a necessary and sufficient condition for the corresponding model to be smooth at $\boldsymbol{\theta_0}$. Hence, if $\boldsymbol{g}$ is partially differentiable at $\boldsymbol{\theta_0}$, we have that*

$$\boldsymbol{g}(\boldsymbol{\theta_0} + th) - \boldsymbol{g}(\boldsymbol{\theta_0}) = t \cdot J_{\boldsymbol{g}}(\boldsymbol{\theta_0}) \cdot h + o(t), \tag{8}$$

*where $J_{\boldsymbol{g}}(\boldsymbol{\theta_0})$ is the matrix of partial derivatives (i.e., the Jacobian). Hence, the score for the direction $h$ in the parameter space is $a_h = J_{\boldsymbol{g}}(\boldsymbol{\theta_0}) \cdot h$.*

**Definition B.3** (Tangent space)**.** *The tangent space*[9] *of the smooth model $\mathcal{M} = \{\boldsymbol{g}(\boldsymbol{\theta}) : \boldsymbol{\theta} \in \Theta \subseteq \mathbb{R}^l\}$ at an interior point $\boldsymbol{\theta_0}$, denoted by $\mathcal{T}$, is the collection of scores: $\mathcal{T} := \{a_h : h \in \mathbb{R}^l\}$. This coincides with the column space of $J_{\boldsymbol{g}}(\boldsymbol{\theta_0})$ per the above remark (which follows from Remark 7.*

The tangent space essentially contains all directions in which one can move in the $\boldsymbol{y}$-space, starting from the true vector $\boldsymbol{y_0} = \boldsymbol{g}(\boldsymbol{\theta_0})$ while staying in the model $\mathcal{M}$. If the tangent space is $\mathbb{R}^k$, one can move in any direction. Hence, the model $\mathcal{M}$ then does not restrict the $\boldsymbol{y}$-space (at least locally around $\boldsymbol{y_0}$). On the other hand, if the tangent space is a strict subspace of $\mathbb{R}^k$, one can only move $\boldsymbol{y_0}$ in certain directions while staying in the model.

##### B.1.2 Target Parameters

The target parameter captures which part of the vector $\boldsymbol{y}$ one is interested in. To arrive at meaningful efficiency results, we will restrict ourselves to certain types of target parameters that are "smooth" in $\boldsymbol{y}$.

**Definition B.4** (Target parameter and estimand)**.** *The target parameter is a function $\psi : \mathcal{M} \to \mathbb{R}$ that maps vectors in the model to the parameter of interest. The estimand is the target parameter evaluated in the true vector: $\psi(\boldsymbol{y_0})$.*

**Definition B.5** (Pathwise differentiability)**.** *The target parameter $\psi$ is pathwise differentiable with respect to the smooth model $\mathcal{M}$ at an interior point $\boldsymbol{\theta_0}$ of $\Theta$ if, for any $h \in \mathbb{R}^l$, it holds that*

$$\frac{\partial \psi\left(\boldsymbol{g}(\boldsymbol{\theta_0} + th)\right)}{\partial t}\bigg|_{t=0} = a_h^\top \phi \tag{9}$$

*for some $\phi \in \mathbb{R}^k$. The vector $\phi$ is termed a gradient of the target parameter.*

Pathwise differentiable parameters are smooth functions of $\boldsymbol{y}$ in the sense that a small change in $\boldsymbol{\theta_0}$ in the direction $h$ induces a small change in the parameter that only depends on the score associated with the direction $h$.

By Remark 7, we can replace the score $a_h$ in the above definition with $J_{\boldsymbol{g}}(\boldsymbol{\theta_0}) \cdot h$. This fact is used in the following remark.

**Remark 8** (Uniqueness gradient)**.** *If the tangent space is $\mathbb{R}^k$, the gradient is unique. If the tangent space is a lower-dimensional subspace of $\mathbb{R}^k$, the gradient is not unique. Indeed, let $\mathcal{T}^\perp$ be the ($k-v$-dimensional) orthogonal complement of the $v$-dimensional tangent space $\mathcal{T} \subset \mathbb{R}^k$ for $v < k$, and let let $\phi$ be a gradient. Then, $\phi + a$ is also a gradient if and only if $a \in \mathcal{T}^\perp$ because $(J_{\boldsymbol{g}}(\boldsymbol{\theta_0})h)^\top(\phi + a) = (J_{\boldsymbol{g}}(\boldsymbol{\theta_0})h)^\top\phi$.*

[9] In semi-parametric efficiency theory, the collection of scores is termed the tangent set, while the closed linear span of the tangent set is termed the tangent space (van der Vaart, 2000, Chapter 25). Because we restricted ourselves to a particular type of models, these concepts translated to the current setting coincide.

**Remark 9.** *If the target parameter is $\psi\left(\boldsymbol{g}(\boldsymbol{\theta})\right) = \theta_j$ where $\theta_j$ is the j'th element of $\boldsymbol{\theta}$, then the target parameter is pathwise differentiable if $J_{\boldsymbol{g}}(\boldsymbol{\theta_0})$ is full rank. By construction we have that $\psi(\boldsymbol{g}(\boldsymbol{\theta_0}+th)) = \boldsymbol{\theta_0}+th$. Let $\mathbf{1}_j$ be the l-dimensional vector with a 1 at the j'th position and 0 at all other positions, such that $\theta_j = \boldsymbol{\theta}^\top \mathbf{1}_j$. We then also have that*

$$\frac{\partial \psi\left(\boldsymbol{g}(\boldsymbol{\theta_0}+th)\right)}{\partial t}\bigg|_{t=0} = h^\top \mathbf{1}_j = (J_{\boldsymbol{g}}(\boldsymbol{\theta_0})h)^\top J_{\boldsymbol{g}}(\boldsymbol{\theta_0})\left(J_{\boldsymbol{g}}(\boldsymbol{\theta_0})^\top J_{\boldsymbol{g}}(\boldsymbol{\theta_0})\right)^{-1}\mathbf{1}_j. \tag{10}$$

*A gradient in this case is thus $\phi = J_{\boldsymbol{g}}(\boldsymbol{\theta_0})\left(J_{\boldsymbol{g}}(\boldsymbol{\theta_0})^\top J_{\boldsymbol{g}}(\boldsymbol{\theta_0})\right)^{-1}\mathbf{1}_l$.*

##### B.1.3 Estimators

An estimator is a function of $\boldsymbol{y}_n$, possibly indexed by $n$, that is supposed to approximate $\psi(\boldsymbol{y}_0)$. In what follows, we denote a general estimator by $T_n$, which actually refers to the function $\boldsymbol{y} \mapsto T_n(\boldsymbol{y})$ evaluated in $\boldsymbol{y}_n$. We next define properties of general estimators for the target parameter. The definitions of the following two properties resemble the corresponding definitions in van der Vaart (2000).

**Definition B.6** (Asymptotic linearity). *An estimator $T_n$ for $\psi(\boldsymbol{y_0})$ is asymptotically linear if*

$$n^{1/2}\left(T_n - \psi(\boldsymbol{y_0})\right) = n^{1/2}\nabla_{T_n}^\top\left(\boldsymbol{y}_n - \boldsymbol{y_0}\right) + o_{P_{0,n}}(1), \tag{11}$$

*where $P_{0,n}$ is a sequence of measures satisfying (7). We term $\nabla_{T_n} \in \mathbb{R}^k$ the influence function of $T_n$.*

It follows that $n^{1/2}\left(T_n - \psi(\boldsymbol{y_0})\right) \underset{P_{0,n}}{\overset{d}{\to}} \mathcal{N}(0, \nabla_{T_n}^\top \Sigma_0 \nabla_{T_n})$ by the continuous mapping theorem and Slutky's lemma. Hence, under the current setup, any asymptotically linear estimator is root-$n$ consistent and asymptotically normal.

The following definition of regularity requires the estimator to obtain its limiting distribution in a "locally uniform" way and is essentially the same definition as in van der Vaart (2000, p. 115).

**Definition B.7** (Regular estimator). *Let $\mathcal{M}$ be a smooth and correctly specified model such that $\boldsymbol{y_0} = \boldsymbol{g}(\boldsymbol{\theta_0})$ for $\boldsymbol{\theta_0}$ an interior point of $\Theta$. Let $\boldsymbol{\theta}_n := \boldsymbol{\theta_0} + n^{-1/2}h$ for $h \in \mathbb{R}^l$ and let $P_{1,n}$ be a sequence of measures such that $n^{1/2}\left(\boldsymbol{y}_n - \boldsymbol{g}(\boldsymbol{\theta}_n)\right) \underset{P_{1,n}}{\overset{d}{\to}} \mathcal{N}\left(\mathbf{0}, \Sigma_0\right)$. An estimator $T_n$ for $\psi(\boldsymbol{y_0})$ is regular if*

$$n^{1/2}\left(T_n - \psi(\boldsymbol{g}(\boldsymbol{\theta}_n))\right) \underset{P_{1,n}}{\overset{d}{\to}} L(\boldsymbol{\theta_0}) \tag{12}$$

*where $L(\boldsymbol{\theta_0})$ is a measure that does not depend on the direction of approximation $h$.*

#### B.2 Efficiency

With the concepts defined above, we are now ready to state and prove some efficiency results for estimators of $\psi(\boldsymbol{y_0})$. The following two lemmas imply that the set of gradients, as characterized in Remark 8, coincides with the set of influence functions of regular and asymptotically linear estimators, further referred to as RAL estimators. Since influence functions characterize RAL estimators (up to asymptotic equivalence), the set of gradients also characterizes the set of RAL estimators. Hence, to study this class of estimators, we can study the set of gradients.

**Lemma B.1.** *Let $\psi : \mathbb{R}^k \to \mathbb{R}$ be a pathwise differentiable parameter with respect to the smooth and correctly specified model $\mathcal{M} := \{\boldsymbol{g}(\boldsymbol{\theta}) : \boldsymbol{\theta} \in \Theta\}$ where $\boldsymbol{y_0} = \boldsymbol{g}(\boldsymbol{\theta_0})$ for $\boldsymbol{\theta_0}$ in the interior of $\Theta$. For every RAL estimator $T_n$ (with respect to $\mathcal{M}$ at $\boldsymbol{\theta_0}$) holds that $\nabla_{T_n}^\top a = \phi^\top a \;\forall\, a \in \mathcal{T}$ where $\mathcal{T}$ is the tangent space of $\mathcal{M}$ at $\boldsymbol{\theta_0}$, $\nabla_{T_n}$ is the influence function of $T_n$, and $\phi$ is a gradient of $\psi$.*

*Proof.* Let $T_n$ be an arbitrary RAL estimator. Choose an arbitrary $h \in \mathbb{R}^l$ and define $\boldsymbol{\theta}_n := \boldsymbol{\theta_0} + n^{-1/2}h$. We let $P_{1,n}$ be a sequence of measures on the measurable space $\left(\mathbb{R}^k, \mathcal{B}\right)$, where $\mathcal{B}$ is the Borel $\sigma$-algebra, such that

$$n^{1/2}\left(\boldsymbol{y}_n - \boldsymbol{g}(\boldsymbol{\theta}_n)\right) \sim \mathcal{N}\left(\boldsymbol{0}, \Sigma_0\right) \quad \forall\, n \in \mathbb{N}.$$

We let $P_{0,n}$ be a sequence of measures on the same measurable space such that

$$n^{1/2}\left(\boldsymbol{y}_n - \boldsymbol{y_0}\right) \sim \mathcal{N}\left(\boldsymbol{0}, \Sigma_0\right) \quad \forall\, n \in \mathbb{N}.$$

By Lemma B.4, $P_{1,n}$ and $P_{0,n}$ are mutually contiguous, denoted as $P_{1,n} \lhd\rhd P_{0,n}$.

Because $T_n$ is asymptotically linear, we have that under $P_{0,n}$:

$$n^{1/2}\left(T_n - \psi(\boldsymbol{y_0})\right) = n^{1/2}\nabla_{T_n}^\top(\boldsymbol{y}_n - \boldsymbol{y_0}) + o_{P_{0,n}}(1).$$

Because $P_{1,n}$ and $P_{0,n}$ are mutually contiguous, any statistic that is $o_{P_{0,n}}(1)$ is also $o_{P_{1,n}}(1)$ by Le Cam's first lemma (van der Vaart, 2000, p. 88). Hence, we also have that

$$n^{1/2}\left(T_n - \psi(\boldsymbol{y_0})\right) = n^{1/2}\nabla_{T_n}^\top(\boldsymbol{y}_n - \boldsymbol{y_0}) + o_{P_{1,n}}(1).$$

We now rearrange terms to obtain that

$$\begin{aligned} n^{1/2}\left(T_n - \psi(\boldsymbol{g}(\boldsymbol{\theta}_n))\right) = {} & n^{1/2}\nabla_{T_n}^\top(\boldsymbol{y}_n - \boldsymbol{g}(\boldsymbol{\theta}_n)) + n^{1/2}\nabla_{T_n}^\top\left(\boldsymbol{g}(\boldsymbol{\theta}_n) - \boldsymbol{y_0}\right) \\ & - n^{1/2}\left(\psi(\boldsymbol{g}(\boldsymbol{\theta}_n)) - \psi(\boldsymbol{y_0})\right) + o_{P_{1,n}}(1). \end{aligned}$$

By the regularity of $T_n$ and the definition of $P_{1,n}$, we have that, respectively:

$$n^{1/2}\left(T_n - \psi(\boldsymbol{g}(\boldsymbol{\theta}_n))\right) \underset{P_{1,n}}{\overset{d}{\to}} \mathcal{N}\left(0, \nabla_{T_n}^\top\Sigma_0\nabla_{T_n}\right) \quad \text{and} \quad n^{1/2}\nabla_{T_n}^\top\left(\boldsymbol{y}_n - \boldsymbol{g}(\boldsymbol{\theta}_n)\right) \underset{P_{1,n}}{\overset{d}{\to}} \mathcal{N}\left(0, \nabla_{T_n}^\top\Sigma_0\nabla_{T_n}\right).$$

The above statements can only hold simultaneously if

$$n^{1/2}\nabla_{T_n}^\top\left(\boldsymbol{g}(\boldsymbol{\theta}_n) - \boldsymbol{y_0}\right) - n^{1/2}\left(\psi(\boldsymbol{g}(\boldsymbol{\theta}_n)) - \psi(\boldsymbol{y_0})\right) = o(1).$$

This further implies that

$$\begin{aligned} \nabla_{T_n}^\top \frac{\boldsymbol{g}\left(\boldsymbol{\theta_0} + n^{-1/2}h\right) - \boldsymbol{g}(\boldsymbol{\theta_0})}{n^{-1/2}} &= \frac{\psi\left(\boldsymbol{g}(\boldsymbol{\theta_0} + n^{-1/2}h)\right) - \psi(\boldsymbol{y_0})}{n^{-1/2}} + o(1) \\ \nabla_{T_n}^\top(J_{\boldsymbol{g}}(\boldsymbol{\theta_0})h) &= (J_{\boldsymbol{g}}(\boldsymbol{\theta_0})h)^\top\phi + o(1). \end{aligned}$$

The second equality follows from $\mathcal{M}$ being smooth (and Remark 7) and the pathwise differentiability of $\psi$ (where $\phi$ is a gradient of $\psi$). Because $\nabla_{T_n}^\top(J_{\boldsymbol{g}}(\boldsymbol{\theta_0})h)$ is a scalar, the above display is equivalent to $\nabla_{T_n}^\top(J_{\boldsymbol{g}}(\boldsymbol{\theta_0})h) = \phi^\top(J_{\boldsymbol{g}}(\boldsymbol{\theta_0})h) + o(1)$.

The set $\left\{J_{\boldsymbol{g}}(\boldsymbol{\theta_0})h : h \in \mathbb{R}^k\right\}$ is exactly the tangent space $\mathcal{T}$ of $\mathcal{M}$ at $\boldsymbol{\theta_0}$. Because we started the proof with an arbitrary $h \in \mathbb{R}^k$, we have that

$$\nabla_{T_n}^\top a = \phi^\top a \;\forall\, a \in \mathcal{T}.$$

□

**Lemma B.2.** *Following the same setup and notation as in Lemma B.1, the influence function $\nabla_{T_n}$ of any RAL estimator $T_n$ has the following representation: $\nabla_{T_n} = \phi + a$ for $\phi$ a gradient of $\psi$ and $a \in \mathcal{T}^{\perp}$.*

*Proof.* We can write $\nabla_{T_n}$ as $\phi + (\nabla_{T_n} - \phi)$ for $\phi$ a gradient of $\psi$. The proof is complete if $\nabla_{T_n} - \phi \in \mathcal{T}^{\perp}$. Lemma B.1 implies that $\nabla_{T_n}^{\top} a = \phi^{\top} a \; \forall \; a \in \mathcal{T}$, or equivalently, that $(\nabla_{T_n} - \phi)^{\top} a = 0 \; \forall \; a \in \mathcal{T}$. This directly implies that $\nabla_{T_n} - \phi \in \mathcal{T}^{\perp}$. □

**Corollary B.2.1.** *If the model is non-parametric (i.e., $\mathcal{T} = \mathbb{R}^k$), then there can only be one RAL estimator, up to asymptotic equivalence.*

**Lemma B.3** (Efficient gradient)**.** *Following the same setup and notation as in Lemma B.1, the efficient gradient $\phi_{\mathit{eff}}$ is defined as the gradient with the smallest variance in the sense that $\phi_{\mathit{eff}}^{\top}\Sigma_0\phi_{\mathit{eff}} - \phi^{\top}\Sigma_0\phi \leq 0$ for $\phi$ a gradient of $\psi$. The efficient gradient is given by*

$$\phi_{\mathit{eff}} := \Sigma^{-1} A \left(A^{\top}\Sigma^{-1}A\right)^{-1} A^{\top}\phi,$$

*where $\phi$ is an arbitrary gradient and $A$ is a matrix whose columns space is $\mathcal{T}$. Consequently, the following value is the lower bound for the variance of RAL estimators:*

$$\phi^{\top} A \left(A^{\top}\Sigma^{-1}A\right)^{-1} A^{\top}\phi.$$

*Proof.* The requirement for the efficient gradient leads to the following constrained minimization problem:

$$\boldsymbol{x}^{\top}\Sigma_0\boldsymbol{x} \quad \text{subject to} \quad A^{\top}\boldsymbol{x} = A^{\top}\phi,$$

where $\phi$ is a gradient and $A$ is a matrix whose columns space is $\mathcal{T}$. We follows the method of the Lagrange multiplier to solve this minimization problem. We, therefore, define the following Lagrangian:

$$\mathcal{L}(\boldsymbol{x}, \boldsymbol{\lambda}) := \boldsymbol{x}^{\top}\Sigma_0\boldsymbol{x} + \boldsymbol{\lambda}^{\top}\left(A^{\top}\boldsymbol{x} - A^{\top}\phi\right).$$

We compute the partial derivatives with respect to $\boldsymbol{x}$ and $\boldsymbol{\lambda}$ and set them to zero:

$$\begin{cases} \frac{d\mathcal{L}}{d\boldsymbol{x}} = 2\Sigma_0\boldsymbol{x} + A\boldsymbol{\lambda} = \mathbf{0} \\ \frac{d\mathcal{L}}{d\boldsymbol{\lambda}} = A^{\top}\boldsymbol{x} - A^{\top}\phi = \mathbf{0}. \end{cases}$$

These equations are solved by the following values:

$$\begin{cases} \boldsymbol{x} = \Sigma_0^{-1} A \left(A^{\top}\Sigma_0^{-1}A\right)^{-1} A^{\top}\phi \\ \boldsymbol{\lambda} = -2\left(A^{\top}\Sigma_0^{-1}A\right)^{-1} A^{\top}\phi \end{cases}$$

Hence, the efficient gradient is $\phi_{\text{eff}} = \Sigma^{-1} A \left(A^{\top}\Sigma^{-1}A\right)^{-1} A^{\top}\phi$. The minimized variance follows immediately by plugging in this expression for $\phi_{\text{eff}}$ into $\phi_{\text{eff}}^{\top}\Sigma\phi_{\text{eff}}$ and basic matrix algebra. □

In standard semi-parametric theory, the efficient gradient[10] can be found by projecting any gradient onto the tangent space $\mathcal{T}$ (Tsiatis, 2006). This does not hold in the current setup unless $\Sigma = I$. Indeed, if $\Sigma = I$, we have that

$$\phi_{\text{eff}} = A\left(A^{\top}A\right)^{-1} A^{\top}\phi,$$

which corresponds to the least-squares projection of $\phi$ onto the column space of $A$. By construction, the column space of $A$ is $\mathcal{T}$.

[10]This is often also termed the efficient influence function.

**Corollary B.3.1.** *Since (i) each RAL estimator's limiting distribution follows from its influence function, (ii) every influence function corresponds to a gradient, and (iii) the variance of an estimator with influence function* $\nabla_{T_n}$ *is* $\nabla_{T_n}^\top \Sigma_0 \nabla_{T_n}$*, Lemma B.3 implies that* $\phi_{\mathit{eff}}^\top \Sigma_0 \phi_{\mathit{eff}}$ *is a lower bound for the variance of the class of RAL estimators. An estimator* $T_n$ *that reaches this lower bound is termed efficient.*

**Remark 10.** *Returning to the target parameter defined in Remark 9 and applying the above corollary shows that the variance lower bound is*

$$\mathbf{1}_j^\top \left(J_{\boldsymbol{g}}(\boldsymbol{\theta_0})^\top J_{\boldsymbol{g}}(\boldsymbol{\theta_0})\right)^{-1} J_{\boldsymbol{g}}(\boldsymbol{\theta_0})^\top J_{\boldsymbol{g}}(\boldsymbol{\theta_0}) \left(J_{\boldsymbol{g}}(\boldsymbol{\theta_0})^\top \Sigma^{-1} J_{\boldsymbol{g}}(\boldsymbol{\theta_0})\right)^{-1} J_{\boldsymbol{g}}(\boldsymbol{\theta_0})^\top J_{\boldsymbol{g}}(\boldsymbol{\theta_0}) \left(J_{\boldsymbol{g}}(\boldsymbol{\theta_0})^\top J_{\boldsymbol{g}}(\boldsymbol{\theta_0})\right)^{-1} \mathbf{1}_j,$$

*which further simplifies by basic matrix algebra to the j'th diagonal element of* $\left(J_{\boldsymbol{g}}(\boldsymbol{\theta_0})^\top \Sigma^{-1} J_{\boldsymbol{g}}(\boldsymbol{\theta_0})\right)^{-1}$.

#### B.3 Miscellaneous

The following technical result is used in the efficiency proofs.

**Lemma B.4** (Generalization of problem 2, Chapter 6 in van der Vaart (2000))**.** *Let* $P_n$ *and* $Q_n$ *be a sequence of measures on the measurable space* $\left(\mathbb{R}^k, \mathcal{B}\right)$ *where* $P_n \sim \mathcal{N}(\mathbf{0}, \Sigma/n)$ *and* $Q_n \sim \mathcal{N}(\boldsymbol{\theta}_n, \Sigma/n)$*. Then* $P_n \triangleleft \triangleright Q_n$ *if and only if* $\boldsymbol{\theta}_n = O(n^{-1/2})$*.*

*Proof.* $\Longleftarrow$ . To show this implication, we will use van der Vaart (2000, Example 6.5) which continuous to hold if the weak converge is along a subsequence.

Consider the log-likelihood ratio of $Q_n$ and $P_n$, which is well defined since $Q_n$ and $P_n$ are absolutely continuous with respect to each other for any fixed $n \in \mathbb{N}$:

$$\begin{aligned}\log \frac{dQ_n}{dP_n} &= -\frac{n}{2}\left(X - \boldsymbol{\theta}_n\right)^\top \Sigma^{-1}\left(X - \boldsymbol{\theta}_n\right) + \frac{n}{2} X^\top \Sigma^{-1} X \\ &= n\boldsymbol{\theta}_n^\top \Sigma^{-1} X - \frac{n}{2}\boldsymbol{\theta}_n^\top \Sigma^{-1} \boldsymbol{\theta}_n.\end{aligned}$$

For a given $n$, we have that

$$\log \frac{dQ_n}{dP_n} \overset{P_n}{\sim} \mathcal{N}\left(-\frac{n}{2}\boldsymbol{\theta}_n^\top \Sigma^{-1}\boldsymbol{\theta}_n, n \cdot \boldsymbol{\theta}_n^\top \Sigma^{-1}\boldsymbol{\theta}_n\right).$$

Hence, letting $\mu_n$ and $\sigma_n^2$ be the mean and variance of the above univariate normal distribution, we have that $\mu_n = -\frac{1}{2}\sigma_n^2$.

If $\boldsymbol{\theta}_n = O(n^{-1/2})$, then we have that $\mu_n = O(1)$ and $\sigma_n^2 = O(1)$, and consequently, there exists a subsequence $n_j$ for $j \in \mathbb{N}$ such that $\mu_{n_j} \to \mu$ and $\sigma_{n_j}^2 \to \sigma^2$ where $\mu_{n_j} = -\frac{1}{2}\sigma_{n_j}^2$ still holds and, consequently, $\mu = -\frac{1}{2}\sigma^2$. Along this subsequence, we have that

$$\frac{dQ_{n_j}}{dP_{n_j}} \underset{P_n}{\overset{d}{\to}} e^{\mathcal{N}(\mu, \sigma^2)},$$

which implies that $P_n \triangleleft \triangleright Q_n$ by van der Vaart (2000, Example 6.5).

$\Longrightarrow$ . Assume that $P_n \triangleleft \triangleright Q_n$. We now show that this implies $\boldsymbol{\theta}_n = O(n^{1/2})$ by using the definition of contiguity.

Consider the sequence of sets $A_n := \left\{\boldsymbol{x} \in \mathbb{R}^k : x_1 \Sigma_{11}^{-1/2} \leq \frac{\Phi^{-1}(p_n)}{n^{1/2}}\right\}$ where $x_1$ is the first element of $\boldsymbol{x}$, $\Sigma_{11}$ is the first diagonal element of $\Sigma$, and $\Phi^{-1}$ is the quantile function of the standard normal distribution. Because $P_n \sim \mathcal{N}(\mathbf{0}, \Sigma/n)$, we have that $P_n(A_n) = \Phi\left\{\Phi^{-1}(p_n)\right\} = p_n$. We further let $p_n \to 0$ and, consequently, $P_n(A_n) \to 0$. Contiguity requires that $Q_n(A_n) \to 0$ as well.

By elementary probability theory, we have that for $\theta_{1,n}$ the first element of $\boldsymbol{\theta}_n$:

$$
\begin{aligned}
Q_n(A_n) &= Q_n\left(X_1 \Sigma_{11}^{-1/2} \leq \frac{\Phi^{-1}(p_n)}{n^{1/2}}\right) \\
&= Q_n\left(n^{1/2}(X_1 - \theta_{1,n})\Sigma_{11}^{-1/2} \leq \Phi^{-1}(p_n) - n^{1/2}\theta_{1,n}\Sigma_{11}^{-1/2}\right) \\
&= \Phi\left(\Phi^{-1}(p_n) - n^{1/2}\theta_{1,n}\Sigma_{11}^{-1/2}\right),
\end{aligned}
$$

where we used the fact that $n^{1/2}(X_1 - \theta_{1,n})\Sigma_{11}^{-1/2} \overset{Q_n}{\sim} \mathcal{N}(0,1)$.

We now see that $Q_n(A_n) \to 0$ can only hold if $\Phi^{-1}(p_n) - n^{1/2}\theta_{1,n}\Sigma_{11}^{-1/2} \to -\infty$. This can only hold if either $n^{1/2}\theta_{1,n} = O(1)$ or $n^{1/2}\theta_{1,n} \to +\infty$ (because we anyhow have that $\Phi^{-1}(p_n) \to -\infty$).

We now consider the sequence of sets $A'_n := \left\{\boldsymbol{x} \in \mathbb{R}^k : x_1 \Sigma_{11}^{-1/2} \geq \frac{\Phi^{-1}(1-p_n)}{n^{1/2}}\right\}$ and repeat the same arguments. Because $P_n \sim \mathcal{N}(\mathbf{0}, \Sigma/n)$, we have that $P_n(A'_n) = 1 - \Phi\left\{\Phi^{-1}(1-p_n)\right\} = p_n$. We further let $p_n \to 0$ and, consequently, $P_n(A'_n) \to 0$. Contiguity requires that $Q_n(A'_n) \to 0$ as well and by elementary probability theory, we have that:

$$
\begin{aligned}
Q_n(A'_n) &= Q_n\left(X_1 \Sigma_{11}^{-1/2} \geq \frac{\Phi^{-1}(1-p_n)}{n^{1/2}}\right) \\
&= Q_n\left(n^{1/2}(X_1 - \theta_{1,n})\Sigma_{11}^{-1/2} \geq \Phi^{-1}(1-p_n) - n^{1/2}\theta_{1,n}\Sigma_{11}^{-1/2}\right) \\
&= 1 - \Phi\left(\Phi^{-1}(1-p_n) - n^{1/2}\theta_{1,n}\Sigma_{11}^{-1/2}\right).
\end{aligned}
$$

We now see that $Q_n(A'_n) \to 0$ can only hold if $\Phi^{-1}(1-p_n) - n^{1/2}\theta_{1,n}\Sigma_{11}^{-1/2} \to +\infty$. This can only hold if either $n^{1/2}\theta_{1,n} = O(1)$ or $n^{1/2}\theta_{1,n} \to -\infty$ (because we anyhow have that $\Phi^{-1}(1-p_n) \to +\infty$). Together with the result in the previous paragraph, this implies that $\theta_{1,n} = O(n^{-1/2})$.

We can repeat the same arguments for the other elements of $\boldsymbol{\theta}_n$ which leads to the requirement that $\boldsymbol{\theta}_n = O(n^{-1/2})$. □

#### B.4 Application to the Estimation of Acceleration Factors

We apply the efficiency theory developed above to the estimation of time-specific and common acceleration factors.

##### B.4.1 Time-Specific Acceleration Factors

When dealing with time-specific acceleration factors, the (implicitly assumed) model is as follows:

$$
\mathcal{M} = \{\boldsymbol{g}(\boldsymbol{\theta}) : \boldsymbol{\theta} \in \Theta\}
$$

for $\boldsymbol{\theta} = (\boldsymbol{\alpha}^\top, \gamma_1, \ldots, \gamma_K)^\top$, $\boldsymbol{g}(\boldsymbol{\theta}) = (\boldsymbol{\alpha}^\top, f_0(\gamma_1 \cdot t_1; \boldsymbol{\alpha}), \ldots, f_0(\gamma_K \cdot t_K; \boldsymbol{\alpha}))^\top$, and $\Theta = \mathbb{R}^{2K+1}$.

**Remark 11.** *In the derivations of estimators for the time-specific acceleration factors, we did not explicitly assume a particular model, already implying that the model is non-parametric. For the sake of the argument, however, we make the underlying model explicit here. This will indeed turn out to be a non-parametric model.*

Let $J_{\boldsymbol{g}}(\boldsymbol{\theta})$ be the Jacobian of $\boldsymbol{\theta} \mapsto \boldsymbol{g}(\boldsymbol{\theta})$. From Remark 7 follows that the tangent space of $\mathcal{M}$ at $\boldsymbol{\theta_0}$ is the column space of $J_{\boldsymbol{g}}(\boldsymbol{\theta_0})$. Hence, if this column space is $\mathbb{R}^{2K+1}$, where $2K+1$ is the dimension of the data, the model is non-parametric. The Jacobian is as follows:

$$J_{\boldsymbol{g}}(\boldsymbol{\theta_0}) = \begin{pmatrix} I_{K+1} & \mathbf{0}_{(\boldsymbol{K}+\mathbf{1})\times \boldsymbol{K}} \\ A & D_\gamma \end{pmatrix},$$

where

$$A := \begin{pmatrix} \frac{\partial f_0(\gamma_1 \cdot t_1;\boldsymbol{\alpha})}{\partial \boldsymbol{\alpha}^\top}\Big|_{\boldsymbol{\alpha}=\boldsymbol{\alpha_0}} \\ \vdots \\ \frac{\partial f_0(\gamma_K \cdot t_K;\boldsymbol{\alpha})}{\partial \boldsymbol{\alpha}^\top}\Big|_{\boldsymbol{\alpha}=\boldsymbol{\alpha_0}} \end{pmatrix} \quad \text{and} \quad D_\gamma := \begin{pmatrix} \frac{\partial f_0(\gamma_1 \cdot t_1;\boldsymbol{\alpha})}{\partial \gamma_1}\Big|_{\gamma_1=\gamma_{1,0}} & \cdots & 0 \\ \vdots & \ddots & \vdots \\ 0 & \cdots & \frac{\partial f_0(\gamma_K \cdot t_K;\boldsymbol{\alpha})}{\partial \gamma_K}\Big|_{\gamma_K=\gamma_{K,0}} \end{pmatrix}. \quad (13)$$

This Jacobian exists (i.e., $\boldsymbol{\theta} \mapsto \boldsymbol{g}(\boldsymbol{\theta})$ is partially differentiable at $\boldsymbol{\theta_0}$) by Assumptions 2 and A.1. The former assumption ensures that $\gamma_{0,j} \cdot t_j \in [0, t_K]$ and the latter ensures that $\gamma_j \mapsto f_0(\gamma_j \cdot t_j; \boldsymbol{\alpha_0})$ is differentiable for $\gamma_j$ such that $\gamma_j \cdot t_j \in [0, t_K + \delta]$.

Under additionally Assumption 1 we have that $\frac{\partial f_0(t;\boldsymbol{\alpha_0})}{\partial t} \neq 0$ for $t \in [0, t_K]$. This implies that the above matrix is of full rank (i.e., of rank $2K+1$); consequently, the tangent space $\mathcal{T}$ is $\mathbb{R}^{2K+1}$ and the model is thus non-parametric. By Corollary B.2.1, there can only be one RAL estimator for pathwise differentiable parameters in this model. From Remark 9 follows that the control group parameters $\boldsymbol{\alpha}$ and time-specific acceleration factors $\boldsymbol{\gamma}$ as target parameters are pathwise differentiable.

We now derive the variance lower bound for $\boldsymbol{\alpha}$ and $\boldsymbol{\gamma}$ as target parameters. This lower bound follows from Remark 10: the variance lower bounds for the elements of $\boldsymbol{\theta}$ are the corresponding diagonal elements of $\left(J_{\boldsymbol{g}}(\boldsymbol{\theta_0})^\top \Sigma_0^{-1} J_{\boldsymbol{g}}(\boldsymbol{\theta_0})\right)^{-1}$. Since $J_{\boldsymbol{g}}(\boldsymbol{\theta_0})$ is invertible with the following inverse

$$J_{\boldsymbol{g}}(\boldsymbol{\theta_0})^{-1} = \begin{pmatrix} I_{K+1} & \mathbf{0}_{(K+1)\times(K)} \\ -D_\gamma^{-1} A & D_\gamma^{-1} \end{pmatrix},$$

we have that $\left(J_{\boldsymbol{g}}(\boldsymbol{\theta_0})^\top \Sigma_0^{-1} J_{\boldsymbol{g}}(\boldsymbol{\theta_0})\right)^{-1} = J_{\boldsymbol{g}}(\boldsymbol{\theta_0})^{-1} \Sigma_0 (J_{\boldsymbol{g}}(\boldsymbol{\theta_0})^\top)^{-1}$, which further simplifies to

$$\begin{pmatrix} I_{K+1} & \mathbf{0}_{(K+1)\times K} \\ -D_\gamma^{-1} A & D_\gamma^{-1} \end{pmatrix} \Sigma_0 \begin{pmatrix} I_{K+1} & -A^\top D_\gamma^{-1} \\ \mathbf{0}_{K\times(K+1)} & D_\gamma^{-1} \end{pmatrix}.$$

We now have that the variance lower bound for $\boldsymbol{\gamma}$ corresponds to the diagonal elements of

$$D_\gamma^{-1} \begin{pmatrix} -A & I_{K+1} \end{pmatrix} \Sigma_0 \begin{pmatrix} -A^\top \\ I_{K+1} \end{pmatrix} D_\gamma^{-1}.$$

This corresponds to the asymptotic variance of the contrast-based estimator of the time-specific acceleration factors, derived in Lemma A.3. It follows similarly that the variance lower bound for $\boldsymbol{\alpha}$ is

$$\begin{pmatrix} I_{K+1} & \mathbf{0}_{(K+1)\times K} \end{pmatrix} \Sigma_0 \begin{pmatrix} I_{K+1} \\ \mathbf{0}_{K\times(K+1)} \end{pmatrix}$$

which corresponds to the $(K+1) \times (K+1)$ submatrix of $\Sigma_0$ in the top-left corner; these are the matrix entries corresponding to the asymptotic variance of $\hat{\boldsymbol{\alpha}}_n$.

##### B.4.2 Common Acceleration Factor

When dealing with a common acceleration factor, the model is as follows:

$$\mathcal{M} = \{\boldsymbol{g}(\boldsymbol{\theta}) : \boldsymbol{\theta} \in \Theta\},$$

for $\boldsymbol{\theta} = (\boldsymbol{\alpha}^\top, \gamma)^\top$, $\boldsymbol{g}(\boldsymbol{\theta}) = (\boldsymbol{\alpha}^\top, f_0(\gamma \cdot t_1; \boldsymbol{\alpha}), \ldots, f_0(\gamma \cdot t_K; \boldsymbol{\alpha}))^\top$, and $\Theta = \mathbb{R}^{K+2}$.

Let $J_{\boldsymbol{g}}(\boldsymbol{\theta})$ be the Jacobian of $\boldsymbol{\theta} \mapsto \boldsymbol{g}(\boldsymbol{\theta})$. From Remark 7 follows that the tangent space of $\mathcal{M}$ at $\boldsymbol{\theta_0}$ is the column space of $J_{\boldsymbol{g}}(\boldsymbol{\theta_0})$. The Jacobian is as follows:

$$J_{\boldsymbol{g}}(\boldsymbol{\theta_0}) = \begin{pmatrix} I_{K+1} & \boldsymbol{0_{(K+1)\times 1}} \\ A & D_\gamma \end{pmatrix} \quad \text{where} \quad D_\gamma := \begin{pmatrix} \left.\frac{\partial f_0(\gamma \cdot t_1; \boldsymbol{\alpha})}{\partial \gamma}\right|_{\gamma=\gamma_0} \\ \vdots \\ \left.\frac{\partial f_0(\gamma \cdot t_K; \boldsymbol{\alpha})}{\partial \gamma}\right|_{\gamma=\gamma_0} \end{pmatrix},$$

and $A$ is as defined for the time-specific acceleration factors in (13). Along the same arguments as for the time-specific acceleration factors, this Jacobian exists by Assumptions 2 and A.1, and under additionally Assumption 1 this Jacobian is of full rank (i.e., of rank $K+2$). Hence, the tangent space $\mathcal{T}$ is the columns space of $J_{\boldsymbol{g}}(\boldsymbol{\theta_0})$, which is a $K+2$-dimensional subspace of $\mathbb{R}^{2K+1}$. There can thus be multiple RAL estimators, that are not asymptotically equivalent, for pathwise differentiable parameters. As for the time-specific acceleration factors, from Remark 9 follows that the control group parameters $\boldsymbol{\alpha}$ and common acceleration factor $\gamma$ as target parameters are pathwise differentiable.

We now derive the variance lower bound for $\boldsymbol{\alpha}$ and $\gamma$ as target parameters. This lower bound follows from Remark 10: the variance lower bounds for each element of $\boldsymbol{\theta}$ are the corresponding diagonal elements of $\left(J_{\boldsymbol{g}}(\boldsymbol{\theta_0})^\top \Sigma_0^{-1} J_{\boldsymbol{g}}(\boldsymbol{\theta_0})\right)^{-1}$. This is exactly the asymptotic variance matrix of the GLS estimator for $(\boldsymbol{\alpha}^\top, \gamma)$, as derived in Lemma A.9. Hence, the GLS estimator is efficient.

### C Additional Information About the Data Application

#### C.1 Missing Data

There is a substantial amount of missingness in the outcome variable that also depends on the treatment group, as shown in Table 1. There is no missingness in the baseline covariates. The analysis presented in the main text is valid under missing at random (MAR) and a correctly specified model (i.e., the linear mixed model should be correctly specified). Under these assumptions, the acceleration factor estimators are also valid. However, small deviations from MAR could have a large impact on the results because there are a lot of missing data. Since MAR is also an untestable assumption, the results should be interpreted with care. To obtain inferences that are robust to certain violations of MAR, one could draw formal inferences in a sensitivity analysis using the so-called intervals of ignorance and uncertainty for the acceleration factors (Vansteelandt et al., 2006).

Table 1: Number and proportion of missing values for the ADAS-Cog scores by measurement occasion and treatment arm. The $p$-values correspond to Fisher's exact tests for equal proportions of missingness between the treatment groups at each measurement occasion.

| Measurement Occasion | Active Treatment, $n = 148$ | Placebo, $n = 79$ | $p$-value |
|---|---|---|---|
| Baseline | 0 (0%) | 0 (0%) | - |
| Week 12 | 7 (4.7%) | 1 (1.3%) | 0.30 |
| Week 24 | 24 (16%) | 4 (5.1%) | 0.018 |
| Week 40 | 39 (26%) | 8 (10%) | 0.004 |
| Week 48 | 52 (35%) | 10 (13%) | <0.001 |

#### C.2 Proportional Slowing Assumption

In the main text, we assessed the proportional slowing assumption informally by comparing the time-specific acceleration factor estimates and formally with the test for proportional slowing based on the minimized GLS criterion. In Figure 1A, we plot the estimated marginal means (EMMs) and the fitted trajectories. The fitted trajectories closely match the EMMs. This visual check thus provides no evidence against proportional slowing.

**Remark 12.** *For the (adaptive) weights-based estimator of the common acceleration factor, the EMMs in the control group are not re-estimated and the estimated reference trajectory is the interpolation function between the control-group EMMs (as shown in Figure 1A). For the GLS estimator of the common acceleration factor, however, the control group means are re-estimated and, consequently, the estimated reference trajectory is the interpolation function between the re-estimated values (i.e., $\tilde{\boldsymbol{\alpha}}_n$ as defined in (7)). This is shown in Figure 1B, which also illustrates that the estimated reference trajectory does not perfectly interpolate between the EMMs in the control group (although the estimated reference trajectory still matches the control group's EMMs closely).*

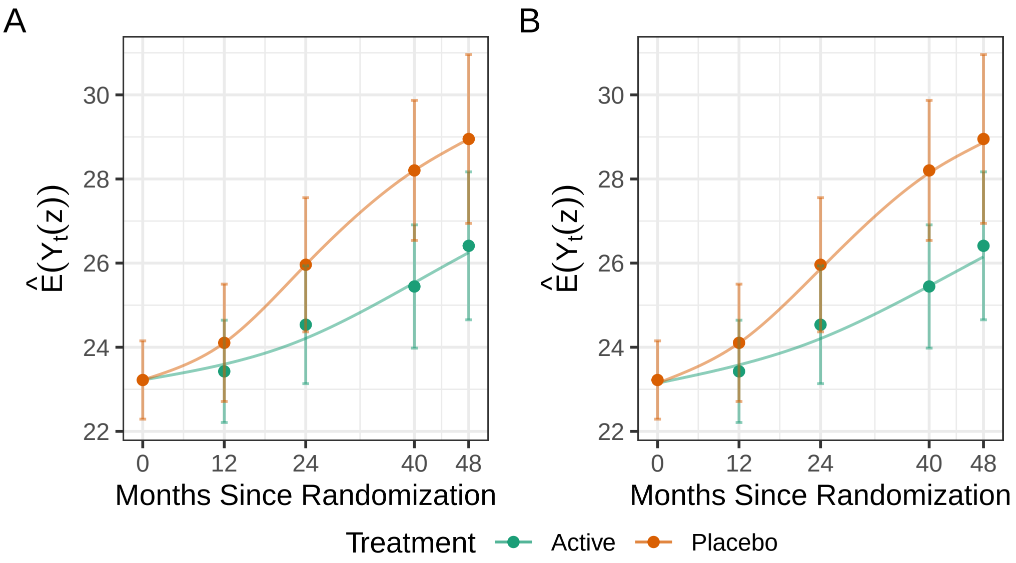

Figure 1: Estimated marginal means (EMMs) with 95% confidence intervals. The full lines are the estimated trajectories. For the placebo group, this is based on natural cubic spline interpolation between (A) the EMMs, $\hat{\boldsymbol{\alpha}}_n$, or (B) the re-estimated control group parameters, $\tilde{\boldsymbol{\alpha}}_n$, as defined in (7). These estimated reference trajectories correspond to $f_0(t;\hat{\boldsymbol{\alpha}}_n)$ and $f_0(t;\tilde{\boldsymbol{\alpha}}_n)$, respectively. For the active treatment group, the estimated trajectory is (A) $f_0(t \cdot \hat{\gamma}_n;\hat{\boldsymbol{\alpha}}_n)$ where $\hat{\gamma}_n = 0.54$ is the adaptive weights-based estimate of the common acceleration factor or (B) $f_0(t \cdot \tilde{\gamma}_n;\tilde{\boldsymbol{\alpha}}_n)$ where $\tilde{\gamma}_n = 0.54$ is the GLS estimate of the common acceleration factor.

### D Simulation Study

In this appendix, we first describe the mixed model for repeated measure (MMRM) that was used in the simulations in more details. Next, we provide additional simulation results for estimators based on natural cubic spline interpolation (some results regarding these estimators were already discussed in the main text) and linear interpolation.

#### D.1 Mixed Model for Repeated Measures

The mixed model for repeated measures (MMRM) is a standard method to analyze longitudinal data in clinical trials (Mallinckrodt et al., 2008) and is the most common primary analysis method in Alzheimer's disease (Dickson et al., 2024; Dickson et al., 2023; Van Dyck et al., 2023). The MMRM is a linear mixed model that makes few assumptions about the mean and covariance structure by including time as a categorical covariate and using an unstructured covariance matrix. In this section, we describe the specific MMRM that was used in the simulations.

The linear predictor of our MMRM contains the interaction effect between time, treated as a categorical covariate, and treatment, except for $t = 0$ where we assume that the mean outcome is equal in both treatment groups. Formally, the mean outcome is modeled as

$$E(Y_{t_j} \mid Z = z) = \begin{cases} \alpha_j \text{ if } z = 0 \\ \beta_j \text{ if } z = 1 \end{cases} \quad \text{for } j \in \{1, \ldots, K\}. \tag{14}$$

and $E(Y_{t_0} \mid Z = 0) = E(Y_{t_0} \mid Z = 1) = \alpha_0$. This essentially means that we have a mean parameter for each time-treatment combination, except at baseline. The covariance matrix is further assumed to be unstructured but common to all subjects[11]. This is a correctly specified model for the data-generating model we consider in the simulations. These MMRMs are fitted with restricted maximum likelihood (REML) using the `mmrm()` function from the *mmrm* R package (Sabanes Bove et al., 2024).

**Remark 13.** *We could also adjust for baseline covariates by including them in the linear predictor in (14). This was done in the MMRM in the data application, but not in the simulations. The time-treatment specific means then depend on baseline covariates, but we can still define and estimate population-level means through the estimated marginal means (EMMs, also known as least-squares means) (Cai, 2014; Searle et al., 1980).*

Conventionally, the treatment effect is expressed in terms of the mean differences at the post-randomization measurement occasions: $(\alpha_1 - \beta_1, \ldots, \alpha_K - \beta_K)'$. To test for a treatment effect in the MMRM, we formulate the following null and alternative hypotheses:

$$H_0 : \alpha_j = \beta_j \; \forall \, j \in \{1, \ldots, K\}$$
$$H_1 : \alpha_j \neq \beta_j \text{ for at least one } j \in \{1, \ldots, K\}.$$

In an MMRM, this null hypothesis can be tested with several methods, among which we will use the $F$-test with the Kenward-Roger approximate degrees-of-freedom (Kenward & Roger, 1997).

In a linear mixed model, the parameter estimates are asymptotically normally distributed if the model is correctly specified, that is,

$$n^{1/2} \left\{ \begin{pmatrix} \hat{\boldsymbol{\alpha}}_n \\ \hat{\boldsymbol{\beta}}_n \end{pmatrix} - \begin{pmatrix} \boldsymbol{\alpha_0} \\ \boldsymbol{\beta_0} \end{pmatrix} \right\} \xrightarrow{d} \mathcal{N}\left(\mathbf{0}, \Sigma_0\right).$$

[11] This is not fully general as the covariance matrix may depend on the assigned treatment.

In the simulations, we estimate the covariance matrix $\Sigma_0$ with the *Asymptotic Covariance* as implemented in the *mmrm* R package. This is the default covariance matrix estimator and is defined as follows:

$$\hat{\Sigma}_n := \left(X^\top \Omega_n^{-1} X\right)^{-1} X^\top \Omega_n X \left(X^\top \Omega_n^{-1} X\right)^{-1},$$

where $X$ is the design matrix and $\Omega_n$ the estimated covariance matrix of the residuals (based on the marginal restricted likelihood).

#### D.2 Additional Results

##### D.2.1 Natural Cubic Spline Interpolation

In Figure 2, we plot the mean squared error (MSE) of the adaptive weights-based and GLS estimators for the common acceleration factor. These plots indicate that the estimator converges to the true common acceleration factor. However, the model is slightly misspecified in all scenarios except the scenario with the 36 months measurement pattern; this is further explained in Appendix D.3. Consequently, in all but the latter scenarios, the estimator will not converge to the truth but to a value near the true common acceleration factor.

In Figures 3 and 4, we plot the empirical standard deviations and the corresponding median estimated standard errors that are based on, respectively, the asymptotic formula (i.e., Lemma A.1) and the parametric bootstrap. The standard errors based on the asymptotic formula underestimate the empirical standard deviation, except for $n = 1000$. Hence, this standard error estimator may not be trustworthy in small to medium samples. In contrast, the median bootstrap standard error closely matches the empirical standard deviation. Only for a very small sample size ($n = 50$) is there some overestimation of the empirical standard deviation. Hence, the parametric bootstrap provides a standard error estimator that is generally valid but some caution remains warranted in very small samples.

In Figure 5, we plot the empirical type 1 error rate and power for testing $H_0 : \gamma_0 = 1$ (i.e., no treatment effect) using the test based on the adaptively weighted contrast function (as described at the end of Appendix A.2.3) and the minimized GLS criterion (as described in Remark 6). The corresponding empirical operating characteristics for the $F$-test in the MMRMs are superimposed in gray. Both types of tests are valid tests for the null of no treatment effect, but the former are 1 degree-of-freedom tests while the latter is a $K$ degrees-of-freedom test. Figure 5 indicates that the type 1 error rate of the GLS test is inflated for the sample sizes between 50 and 500; the power is not well-calibrated here. For $n = 1000$, type 1 error of the GLS test is close to nominal and its power is larger than or equal to the power of the corresponding $F$-test. In contrast, the adaptive weights-based test has a type 1 error rate close to nominal for all sample sizes, but has a lower power than the corresponding $F$-test.

In Figure 6, we plot the empirical type 1 error rate and power for the test of $H_0 : \gamma_0 = 1$ based on the parametric bootstrap ($B = 500$) and the percentile confidence intervals, in combination with the adaptive weights-based or GLS estimator of the common acceleration factor. The parametric-bootstrap tests are conservative for most sample sizes. The one based on the adaptive weights-based estimator generally has lower power than the $F$-test, the one based on the GLS estimator has similar or larger power.

##### D.2.2 Linear Interpolation

For completeness, we reproduce all earlier figures, including those from the main text, for the corresponding estimators and tests based on linear interpolation (instead of natural cubic spline

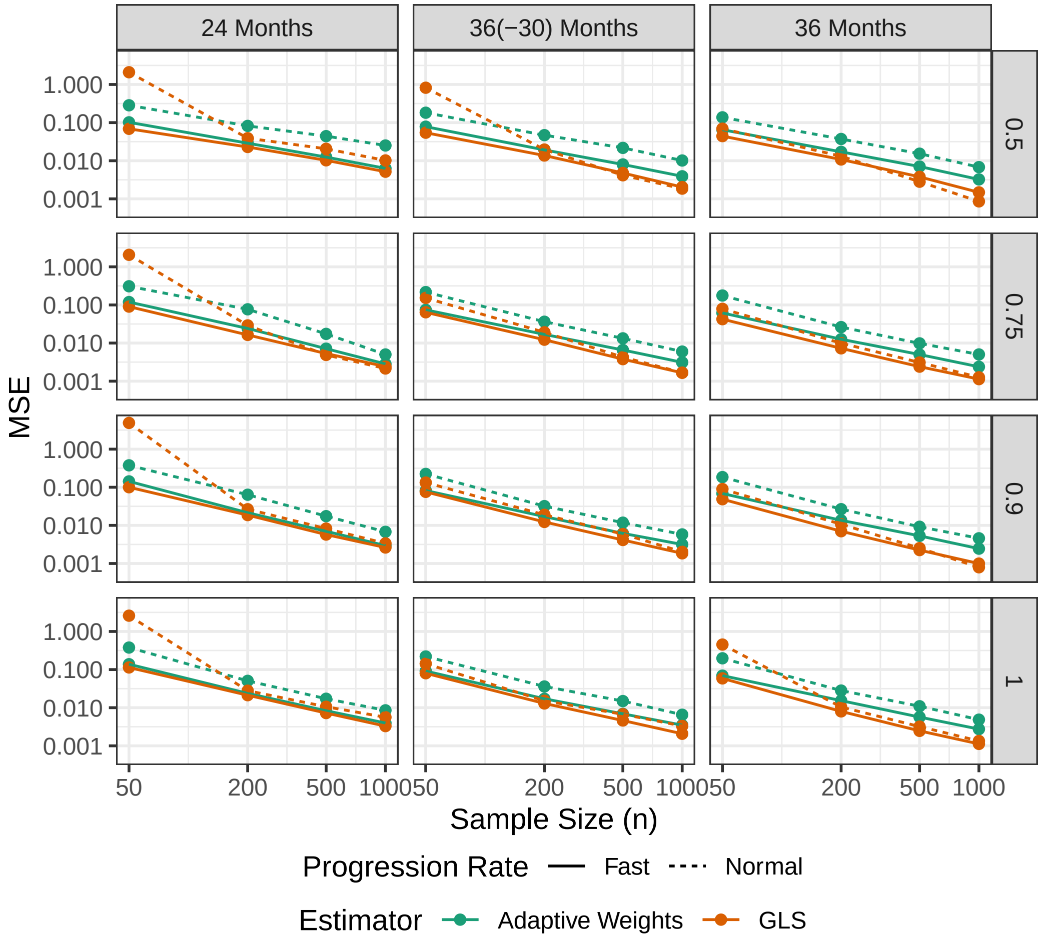

Figure 2: The mean squared error (MSE) of the adaptive weights-based and GLS estimator for the common acceleration factor as function of the sample size. The rows correspond to the true acceleration factors, the columns correspond to the measurement patterns. Note that both axes are log10-transformed.

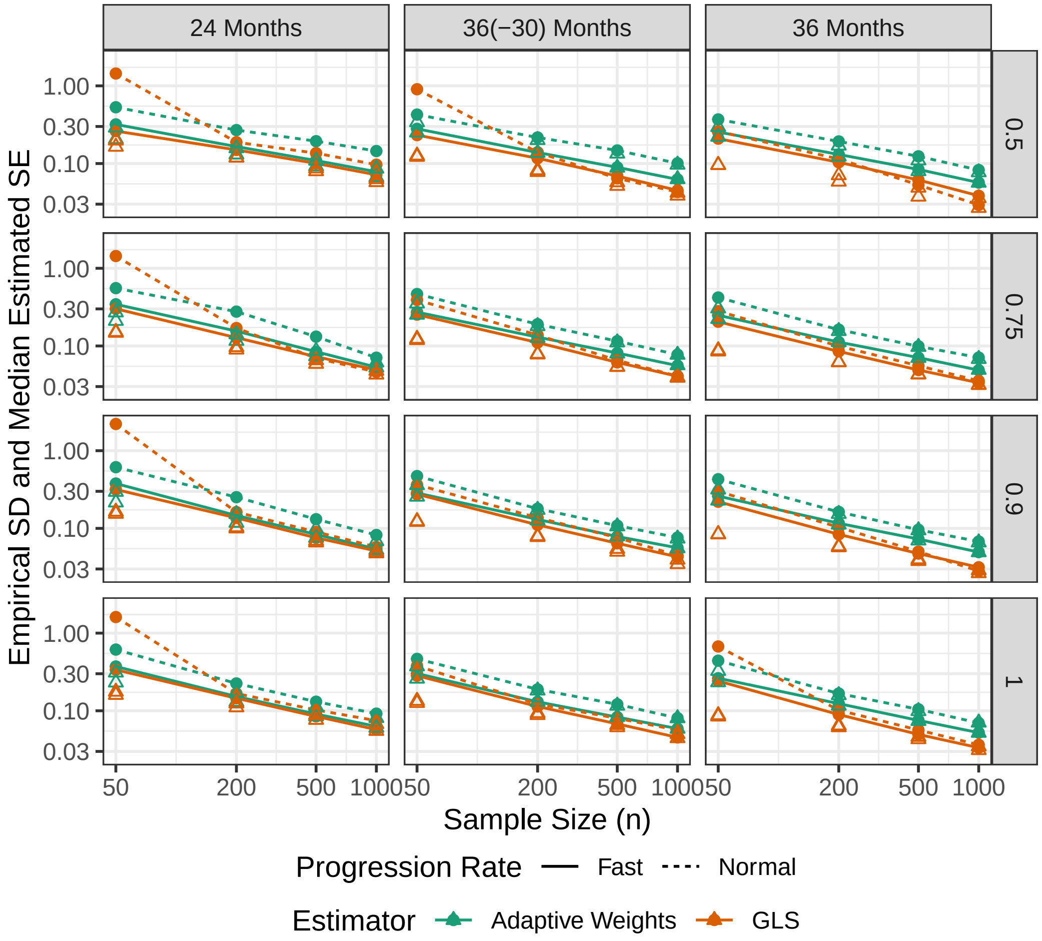

Figure 3: The empirical standard deviations (SDs) and the median estimated standard errors (SEs) of the estimator for the common acceleration factor as functions of the sample size. The SE is estimated based on the asymptotic formula. The dots and connecting lines represent the empirical standard deviations. The triangles represent the median estimated SEs. The rows correspond to the true acceleration factors, the columns correspond to the measurement patterns. Note that both axes are log10-transformed.

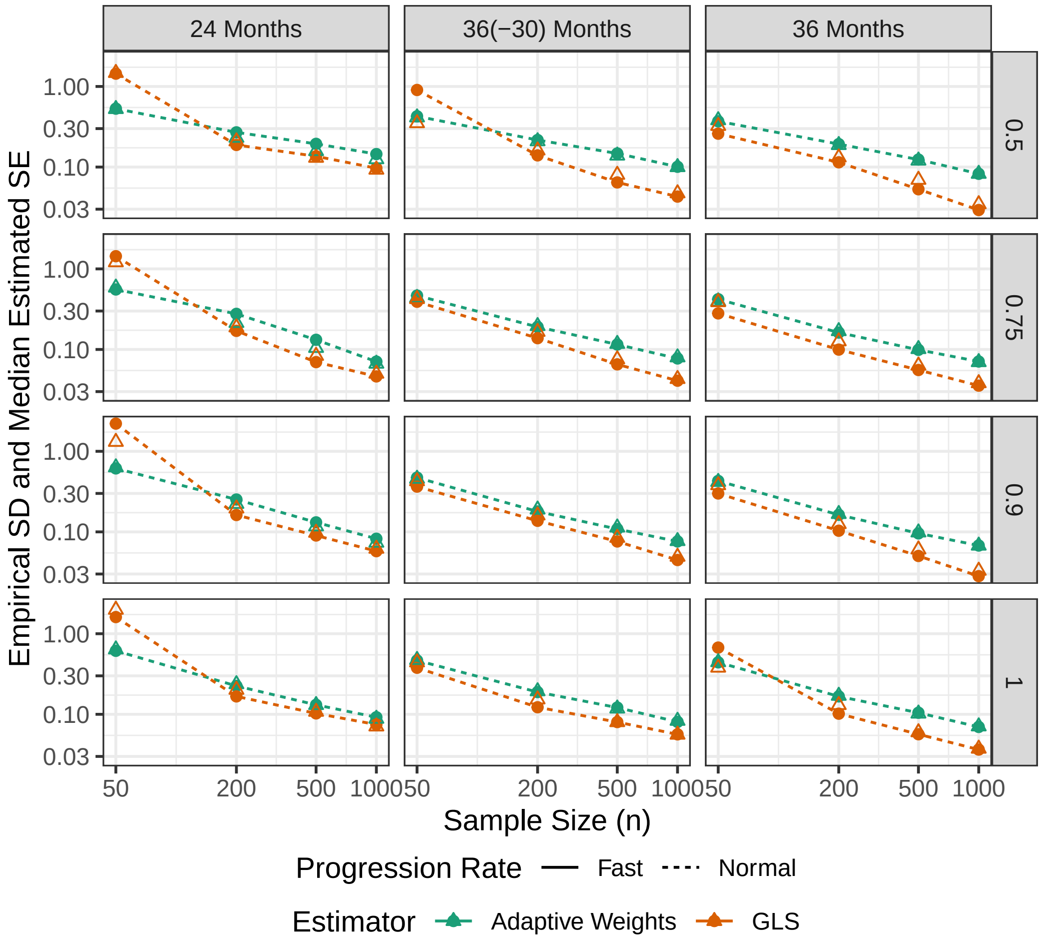

Figure 4: The empirical standard deviations (SDs) and the median estimated standard errors (SEs) of the estimator for the common acceleration factor as functions of the sample size. The SE is estimated based on the parametric bootstrap. The dots and connecting lines represent the empirical standard deviations. The triangles represent the median estimated SEs. The rows correspond to the true acceleration factors, the columns correspond to the measurement patterns. Note that both axes are log10-transformed.

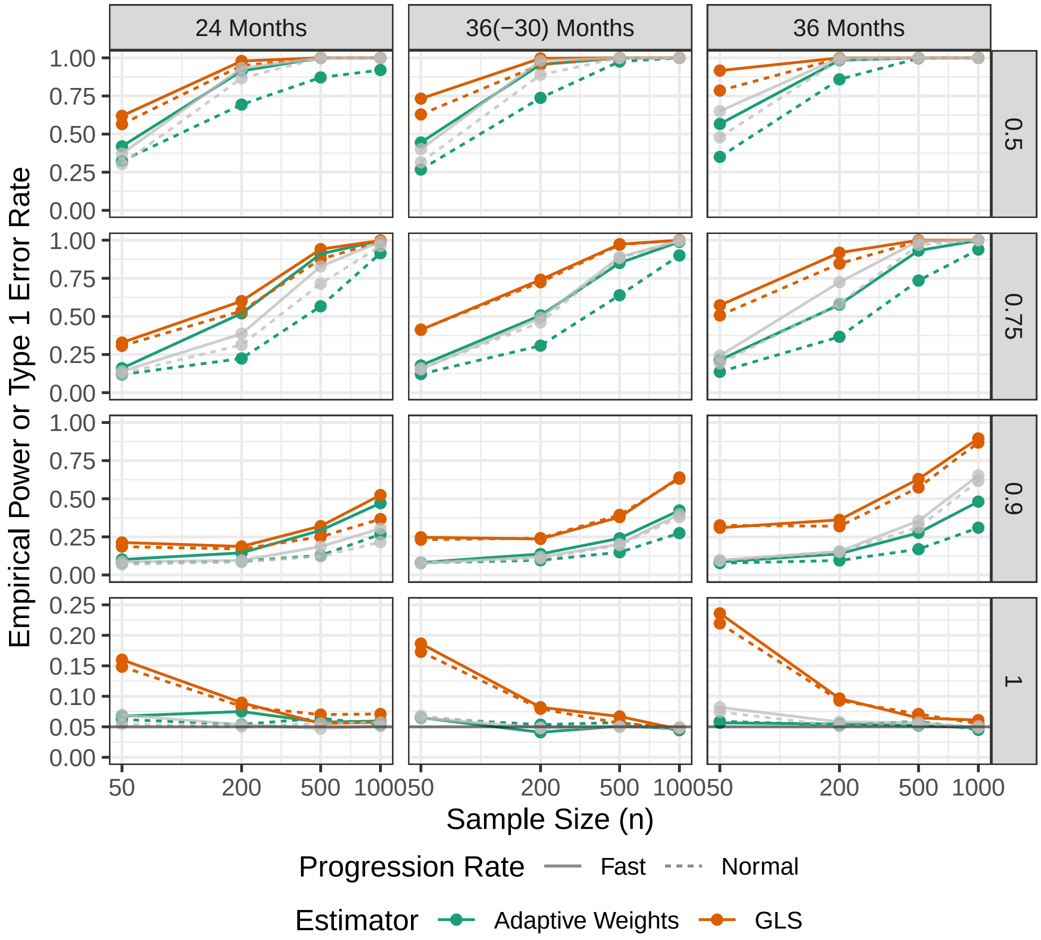

Figure 5: Empirical type 1 error rate and power as functions of the sample size for the testing $H_0 : \gamma_0 = 0$ based on the adaptive weights-based estimator and GLS criterion. The results for the $F$-test for the null of no treatment effect in the MMRM are overlaid in gray. The rows correspond to the true acceleration factors, the columns correspond to the measurement patterns.

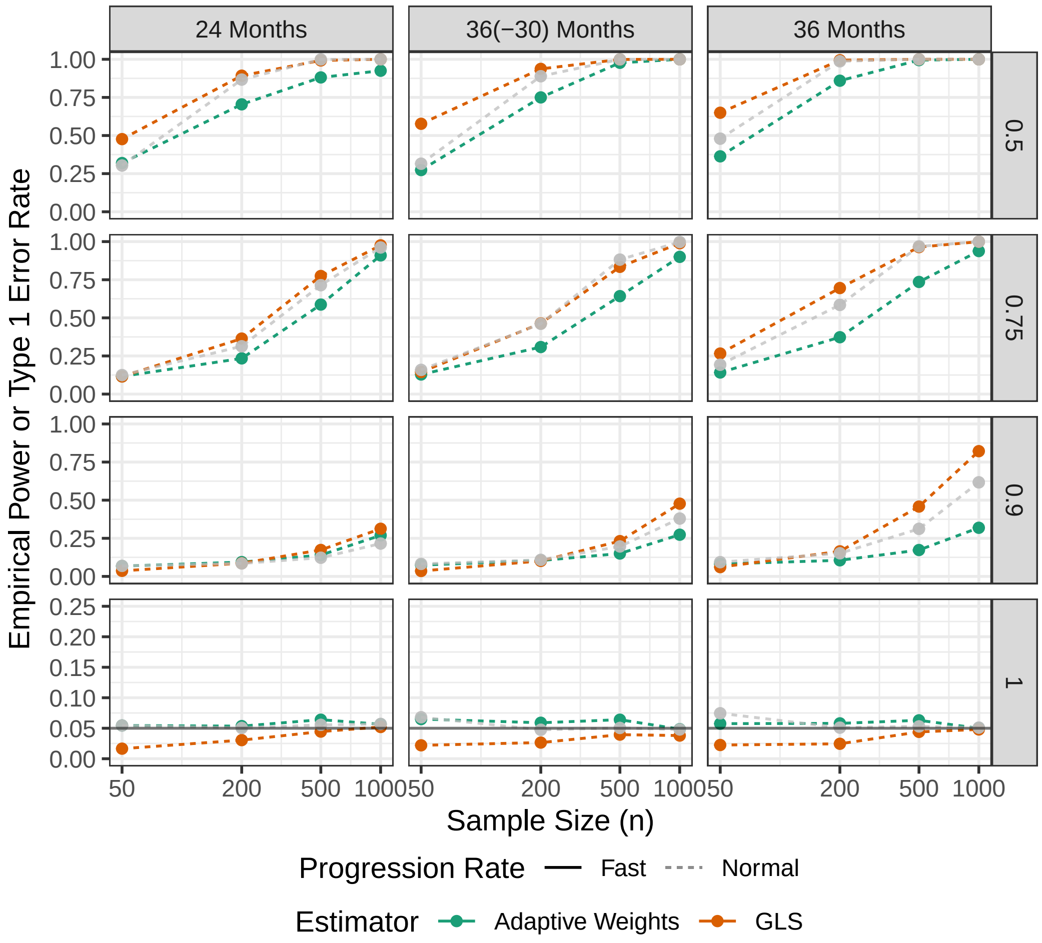

Figure 6: Empirical type 1 error rate and power as functions of the sample size for the testing $H_0 : \gamma = 0$ based on the parametric bootstrap in combination with the adaptive weights-based and GLS estimators of the common acceleration factor. The results for the $F$-test for the null of no treatment effect in the MMRM are overlaid in gray. The rows correspond to the true acceleration factors, the columns correspond to the measurement patterns.

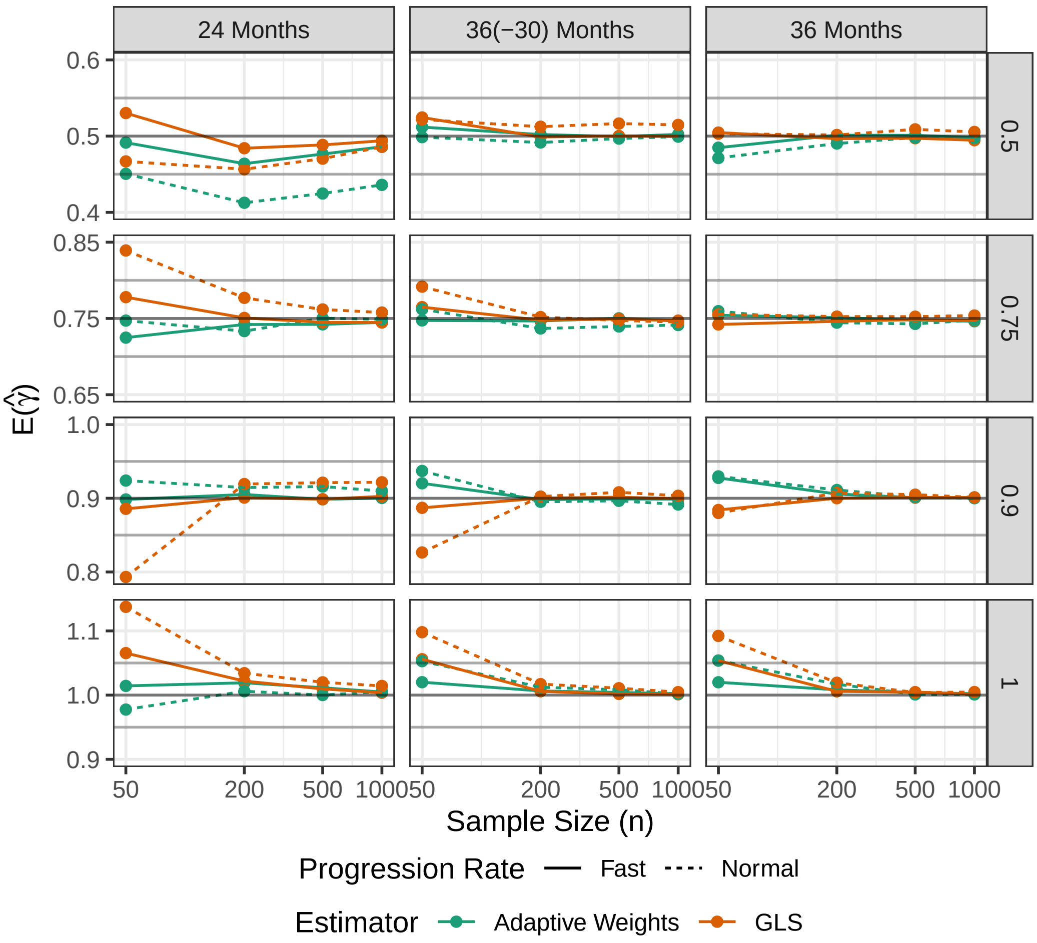

Figure 7: The means of the estimated common acceleration factors as functions of the sample size. The rows correspond to the true common acceleration factors, the columns correspond to the measurement patterns. In each subplot, a black horizontal line represents the true acceleration factor and gray horizontal lines represent the 0.05-margin around the true value.

interpolation) in Figures 7–11. We did not repeat the parametric bootstrap with linear interpolation because of the computational cost of the bootstrap. Note that the data-generating model remained unchanged (i.e., it is still based on natural cubic spline interpolation); consequently, Assumption 6 is violated in all scenarios except for $\gamma = 1$.

#### D.3 Model Misspecification for Correct Interpolation Method

In the simulations presented in the main text, the correct interpolation method (i.e., natural cubic spline interpolation) was used for estimating the acceleration factors. Nonetheless, Assumption 6 is actually violated in the *24 Months* and *36(-30) Months* scenarios. This type of model misspecification is illustrated next.

Consider the true mean ADAS-Cog scores in the normal progression scenario and connect them using natural cubic spline interpolation. This yields in Figure 12 the full red line for $\gamma = 1$ and the full blue line for $\gamma = 0.90$. These trajectories evaluated in $\{0, 6, 12, 18, 24, 30, 36\}$—these are the black dots in Figure 12—are the true means from which the data are generated in the simulations.

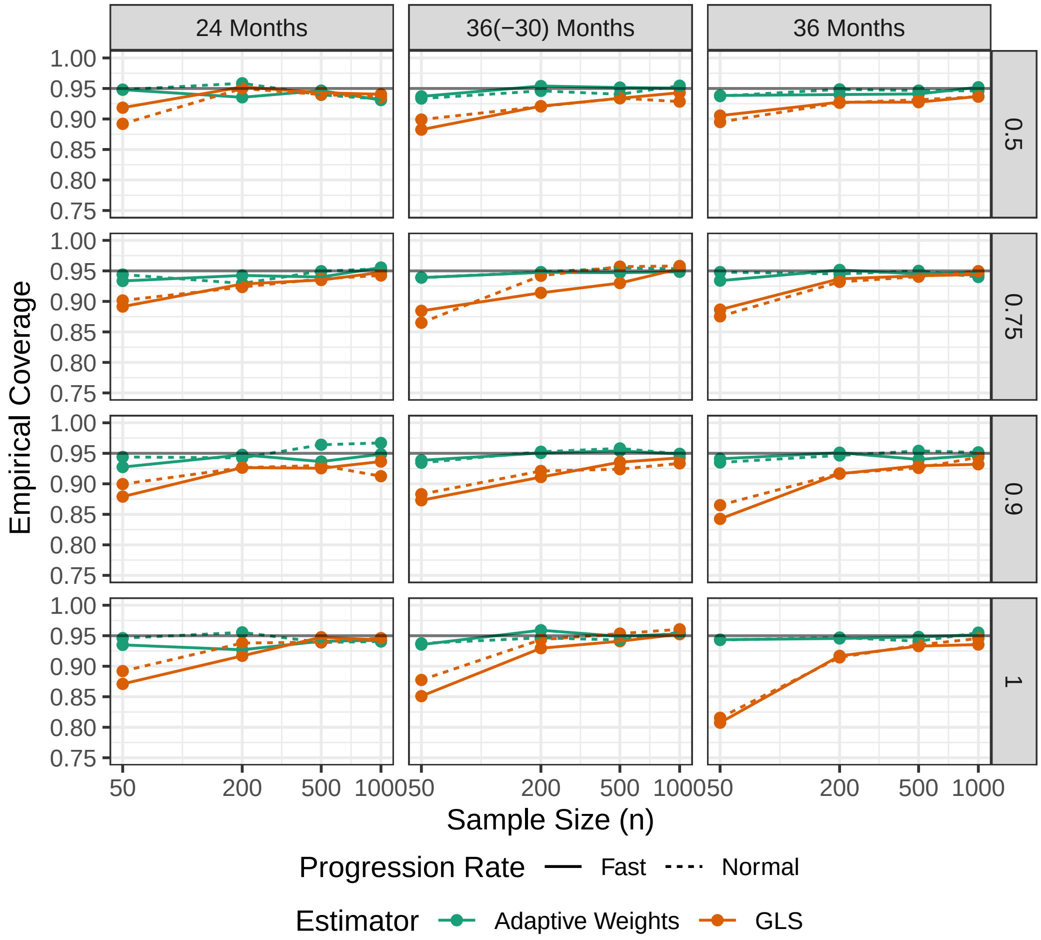

Figure 8: The empirical coverage rates of the confidence intervals based on inverting hypothesis tests as functions of the sample size. The rows correspond to the true acceleration factors, the columns correspond to the measurement patterns. The black horizontal lines indicate 95% coverage.

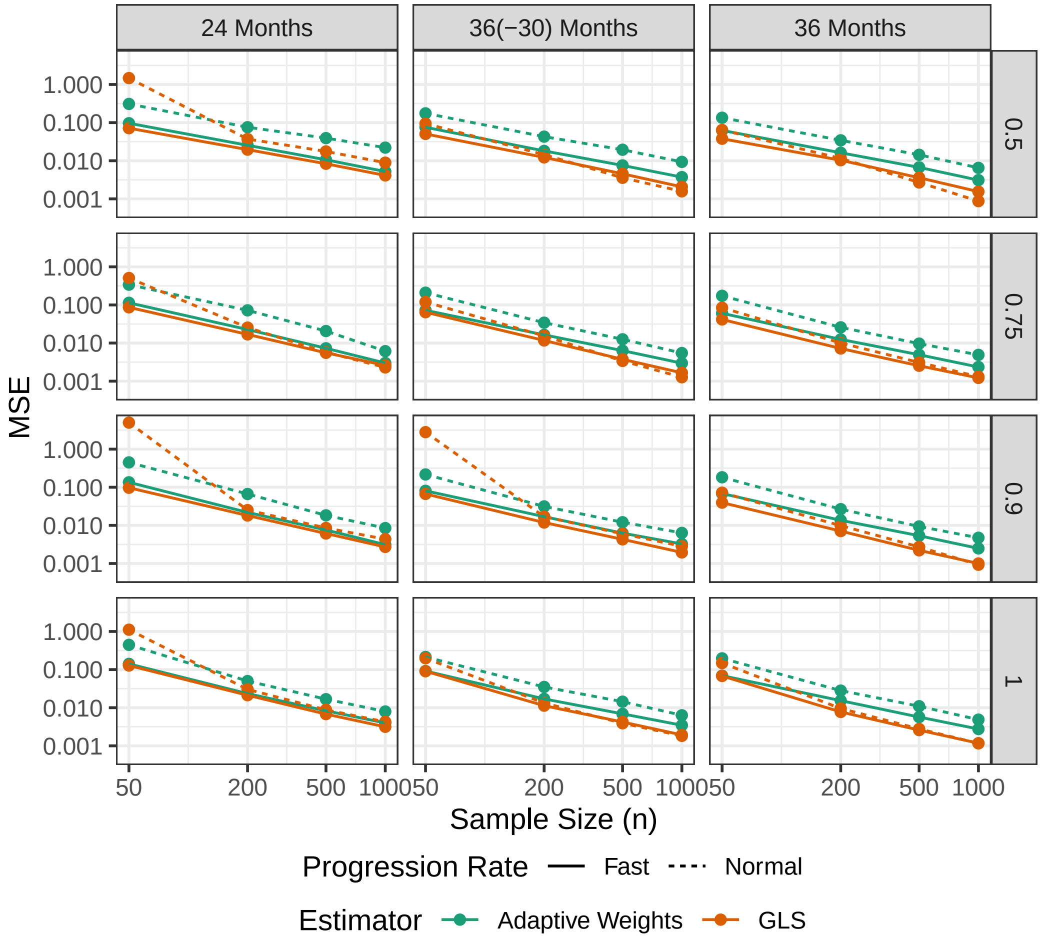

Figure 9: The mean squared error (MSE) of the adaptive weights-based and GLS estimator for the common acceleration factor as function of the sample size. The rows correspond to the true acceleration factors, the columns correspond to the measurement patterns. Note that both axes are log10-transformed.

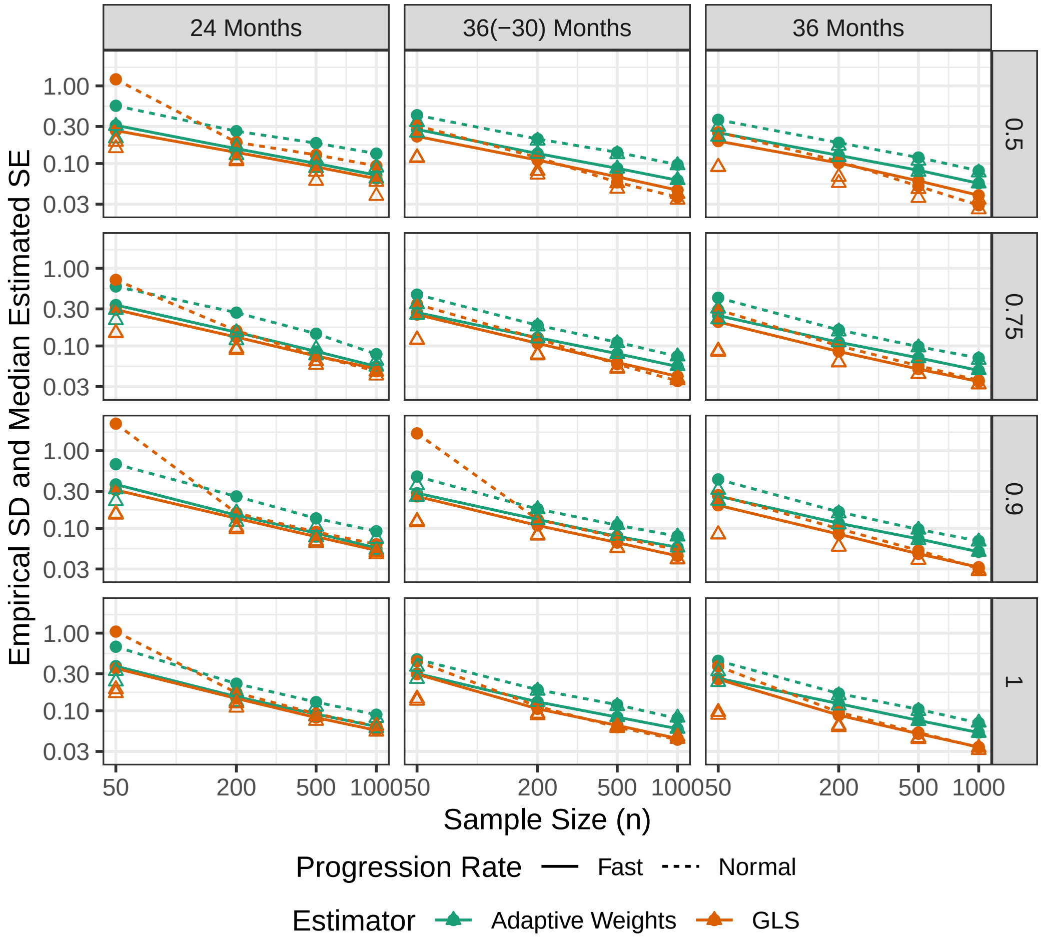

Figure 10: The empirical standard deviations (SDs) and the median estimated standard errors (SEs) of the estimator for the common acceleration factor as functions of the sample size. The SE is estimated based on the asymptotic formula. The dots and connecting lines represent the empirical standard deviations. The triangles represent the median estimated SEs. The rows correspond to the true acceleration factors, the columns correspond to the measurement patterns. Note that both axes are log10-transformed.

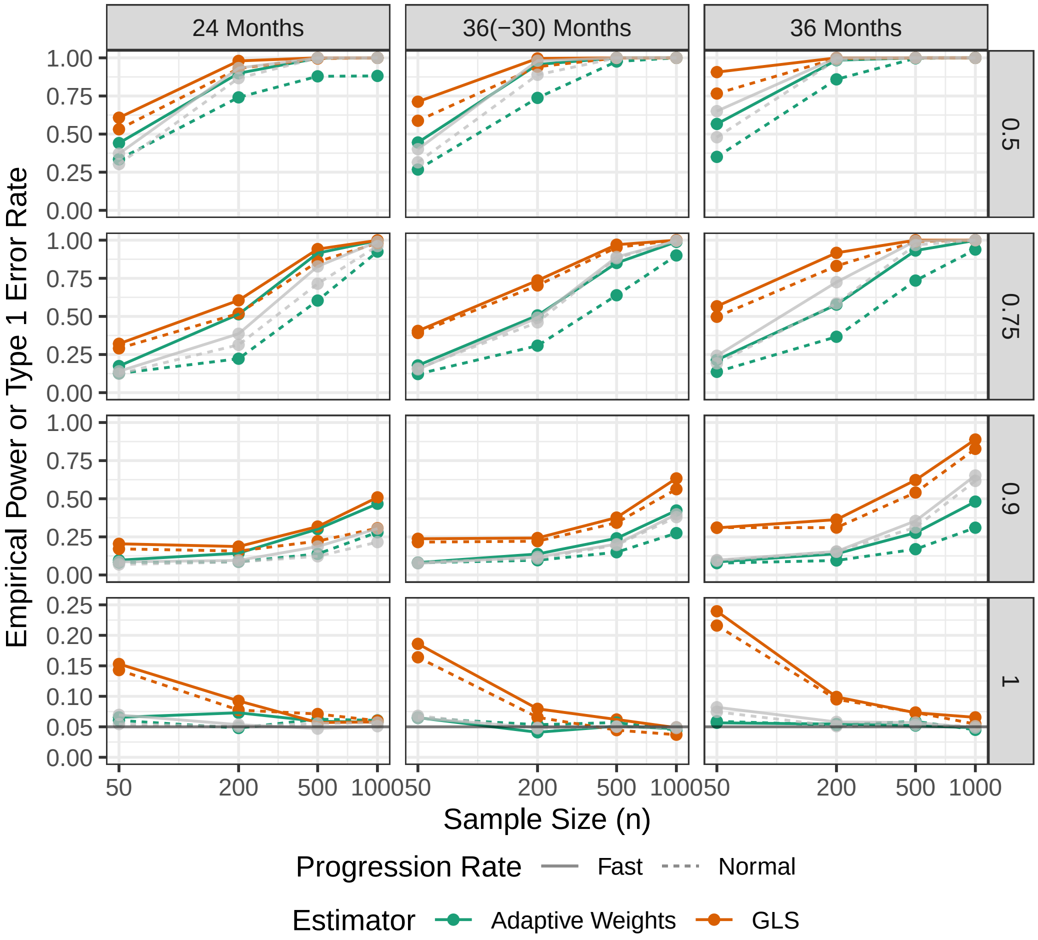

Figure 11: Empirical type 1 error rate and power as functions of the sample size for the testing $H_0 : \gamma_0 = 0$ based on the adaptive weights-based estimator and GLS criterion. The results for the $F$-test for the null of no treatment effect in the MMRM are overlaid in gray. The rows correspond to the true acceleration factors, the columns correspond to the measurement patterns.

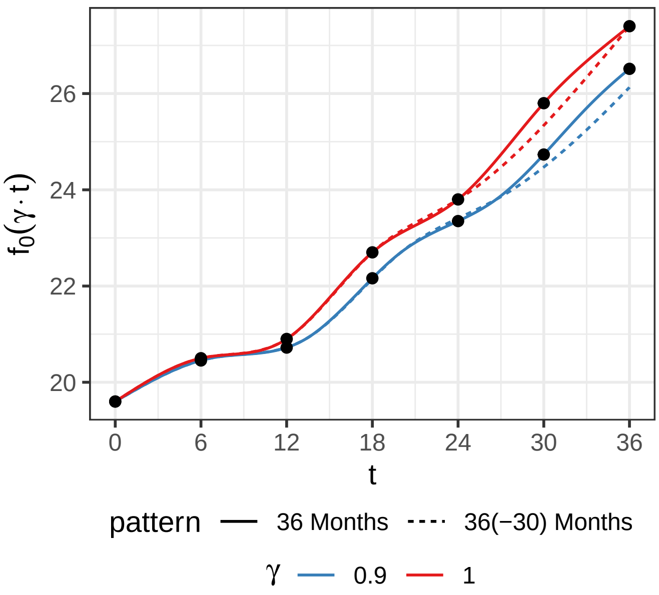

Figure 12: Natural cubic spline interpolating functions for the normal progression true mean vector. The full and dashed lines correspond, respectively, to the interpolating functions for the true mean vector in the 36 Months and 36(-30) Months measurement patterns.

If we repeat the same steps but drop the measurement at 30 months (which corresponds to the *36(-30)* measurement pattern), we obtain the dashed lines. The black dots no longer lie on the interpolated trajectories for $\gamma = 1$ as well as $\gamma = 0.90$. The model is thus misspecified because Assumption 6 is violated, even though the correct interpolation method is apparently used.

**Remark 14.** *This illustration clearly shows that Assumption 6 and the proportional slowing assumption will not hold exactly in practice. Hence, one should always interpret the estimated acceleration factors with some skepticism and view the estimate as a useful summary of the treatment effect that is, nonetheless, arbitrary to some degree. As the number of measurement occasions increases, the sensitivity to the interpolation method should decrease, however.*
